# Prophylactic Treatment of COVID-19 in Care Homes Trial (PROTECT-CH)

**DOI:** 10.1101/2022.08.29.22279359

**Authors:** Philip M Bath, Jonathan Ball, Matthew Boyd, Heather Gage, Matthew Glover, Maureen Godfrey, Bruce Guthrie, Jonathan Hewitt, Robert Howard, Thomas Jaki, Edmund Juszczak, Daniel Lasserson, Paul Leighton, Val Leyland, Wei Shen Lim, Pip Logan, Garry Meakin, Alan Montgomery, Reuben Ogollah, Peter Passmore, Philip Quinlan, Caroline Rick, Simon Royal, Susan D Shenkin, Clare Upton, Adam L Gordon, the PROTECT-CH Trialists

**Affiliations:** Stroke Trials Unit, Mental Health & Clinical Neuroscience, University of Nottingham, Nottingham, NG7 2UH, UK; Stroke, Nottingham University Hospitals NHS Trust, Queen’s Medical Centre, Derby Road, Nottingham NG7 2UH, UK; Infections, Immunity and Microbes, School of Life Sciences, University of Nottingham, Nottingham, NG7 2UH, UK; Division of Pharmacy Practice and Policy, School of Pharmacy, University of Nottingham, NG7 2RD, UK; Surrey Health Economics Centre, Department of Clinical and Experimental Medicine, University of Surrey, Guildford, GU2 7YW, UK; c/o Nottingham Clinical Trials Unit, University of Nottingham, Nottingham, NG7 2UH, UK; Advanced Care Research Centre, Usher Institute, University of Edinburgh, Edinburgh, EH8 9AG, UK; Department of Geriatric Medicine, Llandough Hospital, Penarth, CF64 2XX, UK; Division of Psychiatry, University College London, London W1T 7NF UK; MRC Biostatistics Unit, University of Cambridge, Cambridge, CB2 0SR UK; Nottingham Clinical Trials Unit, University of Nottingham, Nottingham, NG7 2UH, UK; Acute Ambulatory Care, Warwick Medical School, University of Warwick, Coventry, CV4 7AL, UK; Ambulatory Assessment Unit, Department of Geratology, Oxford University Hospitals NHS Foundation Trust, Oxford, OX3 9DU, UK; Lifespan and Population Sciences, School of Medicine, University of Nottingham, Nottingham, NG7 2UH, UK; Bramcote, Nottingham NG9 UK; Respiratory Medicine, Nottingham University Hospitals NHS Trust, Nottingham, NG5 1PB; Unit of Injury, Inflammation and Recovery, School of Medicine, University of Nottingham, NG7 2UH, UK; Nottingham City Care Partnership, Nottingham, NG6 8WR, UK; Centre for Public Health, Institute for Clinical sciences, Queen’s University Belfast, Grosvenor Road, Belfast BT12 6BJ; Digital Health & Digital Research Service, University of Nottingham, Nottingham, NG7 2UH, UK; University of Nottingham Health Service, Cripps Health Centre, University Park, Nottingham NG7 2QW UK; Ageing and Health, Usher Institute and Advanced Care Research Centre, University of Edinburgh, Edinburgh, EH16 4SB, UK; NIHR Applied Research Collaboration-East Midlands (ARC-EM), Institute of Mental Health, Triumph Road, Nottingham. NG7 2TU.; Division of Medical Sciences and Graduate Entry Medicine, University of Nottingham, Derby Medical School, Royal Derby Hospital, Derby, DE22 3NE, UK

**Keywords:** Care home, COVID-19, prevention, nursing home, prophylaxis, randomised controlled trial, residential home

## Abstract

**Background:** Coronavirus disease 2019 (COVID-19) is associated with significant mortality and morbidity in care homes. Novel or repurposed antiviral drugs may reduce infection and disease severity through reducing viral replication and inflammation.

**Objective:** To compare the safety and efficacy of antiviral agents (ciclesonide, niclosamide) for preventing SARS-CoV-2 infection and COVID-19 severity in care home residents.

**Design:** Cluster-randomised open-label blinded endpoint platform clinical trial testing antiviral agents in a post-exposure prophylaxis paradigm.

**Setting:** Care homes across all four United Kingdom member countries.

**Participants:** Care home residents 65 years of age or older.

**Interventions:** Care homes were to be allocated at random by computer to 42 days of antiviral agent plus standard care versus standard of care and followed for 60 days after randomisation.

**Main outcome measures:** The primary four-level ordered categorical outcome with participants classified according to the most serious of all-cause mortality, all-cause hospitalisation, SARS-CoV-2 infection and no infection. Analysis using ordinal logistic regression was by intention to treat. Other outcomes included the components of the primary outcome and transmission.

**Results:** Delays in contracting between NIHR and the manufacturers of potential antiviral agents significantly delayed any potential start date. Having set up the trial (protocol, approvals, insurance, website, database, routine data algorithms, training materials), the trial was stopped in September 2021 prior to contracting of care homes and general practitioners in view of the success of vaccination in care homes with significantly reduced infections, hospitalisations and deaths. As a result, the sample size target (based on COVID-19 rates and deaths occurring in February-June 2020) became unfeasible.

**Limitations:** Care home residents were not approached about the trial and so were not consented and did not receive treatment. Hence, the feasibility of screening, consent, treatment and data acquisition, and potential benefit of post exposure prophylaxis were never tested. Further, contracting between the University of Nottingham and the PIs, GPs and care homes was not completed, so the feasibility of contracting with all the different groups at the scale needed was not tested.

**Conclusions:** The role of post exposure prophylaxis of COVID-19 in care home residents was not tested because of changes in COVID-19 incidence, prevalence and virulence as a consequence of the vaccination programme that rendered the study unfeasible. Significant progress was made in describing and developing the infrastructure necessary for a large scale Clinical Trial of Investigational Medicinal Products in care homes in all four UK nations.

**Future work:** The role of post-exposure prophylaxis of COVID-19 in care home residents remains to be defined. Significant logistical barriers to conducting research in care homes during a pandemic need to be removed before such studies are possible in the required short timescale.

## Chapter 1: Introduction

### 1.1 Background

In 2019 a novel coronavirus-induced disease (COVID-19) emerged in Wuhan, China. A month later the Chinese Center for Disease Control and Prevention identified a new beta-coronavirus (SARS coronavirus 2, or SARS-CoV-2) as the aetiological agent.^1^ Subsequently, the disease has spread around the world causing more than 261 million cases and 5.2 million deaths (COVID-19 Dashboard, Johns Hopkins University, accessed 29 November 2021).^2^ In the UK, 10.2 million cases and 145,218 deaths have been recorded.^2^

SARS-CoV-2 infection has been devastating in care homes causing profound morbidity, mortality and disruption of care home routines to the detriment of the wellbeing of residents, families and staff. By the end of 2020, England had recorded 19,179 deaths due to COVID-19 in care home residents,^3^ this explaining ∼30% of the excess mortality associated with COVID-19 and reducing life expectancy in Scottish care home residents by approximately half a year.^4^ Hygiene measures, prevention of visiting by friends and family of residents, routine testing for COVID-19, and use of personal protection equipment by staff have reduced infection. Nevertheless, outbreaks of infection have continued, and prophylaxis measures were introduced, especially pre-exposure prophylaxis with vaccination.^5, 6^ By December 2021, more than 126 million doses of vaccine had been delivered in the UK, this including first, second, third and booster injections. Vaccination of residents (and staff) started in early 2021 using the Pfizer and AstraZeneca vaccines. Although there were concerns that vaccines might be less effective in older people with multiple comorbidities and immunosenescence, this has not been seen and vaccination of more than 90% of care home residents and 80% of staff has significantly reduced disease, especially that which is severe and leads to hospitalisation and death. Although vaccines were developed against the Wuhan/wild type virus, they have also been sufficiently effective against alpha, beta, gamma and delta variants (European Centre for Disease Prevention and Control, accessed 29/11/2021). The work outlined in this report was conceived before vaccine roll-out, commenced in parallel with the beginning of mass vaccination at the time when the efficacy of vaccination in care home populations remained uncertain, and concluded once the efficacy of vaccination had been established, rendering the proposed work unfeasible. Specifically, PROTECT-CH was designed in October 2020 using data from the significant wave of excess deaths in care homes due to the first/Wuhan wave of infection in March-May 2020 (Figure 1, bottom left panel); apart from a smaller wave related to the alpha variant in January-February 2021, there have been no other periods of excess deaths in care homes. This pattern of excess deaths differs from that seen for deaths at home and in hospital (Figure 1, top panels). The uncertainty around the future course of COVID-19 infection has increased again with the emergence of the Omicron variant, rates of Omicron infections in care homes have been low although international experience of Omicron is limited. A modest reduction in vaccine efficacy would lead to substantial morbidity and mortality and there is still a need for interventions that prevent infections and transmission in care homes.

**Figure 1.**
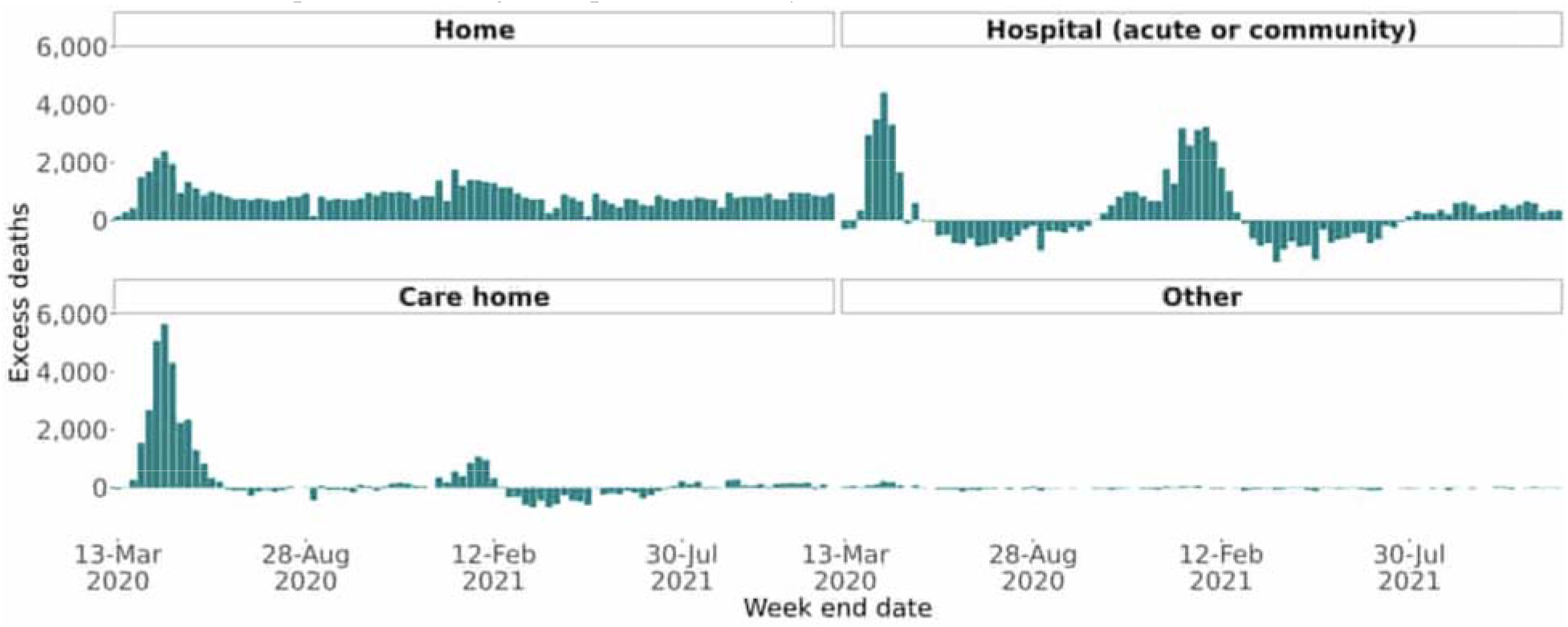
Excess death rates in England & Wales (7 March 2020-10 December 2021) at home (top left), in hospital (top right), in care homes (bottom left) and elsewhere (bottom right). Excess deaths are registered deaths minus the 2015-2019 average by weeks. Source: Office for National Statistics - Deaths registered weekly in England & Wales).

Numerous interventions have demonstrated *in vitro* activity against SARS-CoV-2 and some have been tested clinically. Of these, ciclesonide, a non-halogenated inhaled corticosteroid used in the prophylaxis of asthma, has been shown to block SARS-CoV-2 RNA replication by targeting the viral replication-transcription complex^8^ and inhibit SARS-CoV-2 cytopathic activity^7^. Pharmacodynamic studies have shown that inhaled ciclesonide has potent anti-inflammatory activity in patients with asthma, and does not appear to have clinically relevant systemic effects, even at high doses. In a case series, ciclesonide treatment was associated with higher blood lymphocyte counts, potentially important since lymphopenia is associated with severe COVID-19. Several uncontrolled case series of ciclesonide use in COVID-19 have been reported but the lack of control groups, small size and concurrent testing of other potential antiviral agents limit their interpretation.

Niclosamide anhydrous is a salicylanilide introduced as an oral anthelmintic in the early 1960s for treating tapeworm infestations. Niclosamide is a multimodal drug that inhibits or regulates multiple signalling pathways and biological processes via pleotropic activities. Recent studies have indicated that niclosamide may have broad clinical applications beyond the treatment of parasites and *in vitro* data suggest that niclosamide inhibits SARS-CoV-2 replication and cellular penetration. Niclosamide has also been shown to have non-steroidal anti-inflammatory activity both experimentally and clinically. As such, nasal administration as a spray may be most effective as a post-exposure prophylactic for early-stage infection when viral load is a main issue. Although niclosamide is a substrate and inhibitor of CYP1A2 *in vitro*; intranasal administration is unlikely to lead to plasma levels where CYP inhibition is seen.

The literature relating to COVID-19 prevention and treatment is fast moving and the state of knowledge at the time the trial was designed in late 2020 was far behind what is now known in late 2021, as shown in Table 1. Recently, the efficacy of oral antiviral agents (mulnupiravir,^13, 14^ PF-07321332) for treating COVID-19 has been reported and both have emergency use authorisation from the MHRA.

**Table 1.**
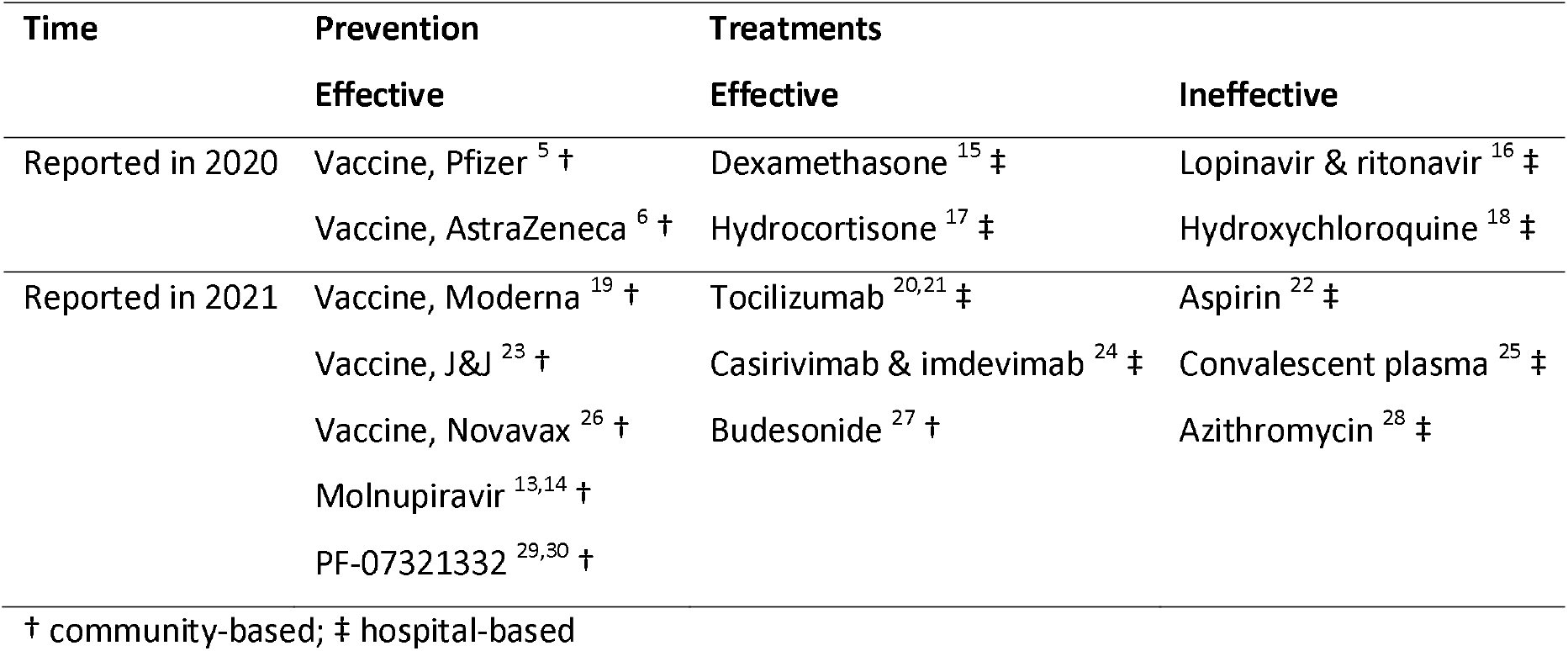
State of knowledge about COVID-19 treatments in 2020 and 2021. Only large trials are quoted. No trials focussed on care home participants.

Here, we report the design, start-up and close-down of the trial platform. The aim was to reduce the severity of COVID-19 in infected residents and transmission of SARS-CoV-2 within care homes.

### 1.2 Rationale for the trial

PROTECT-CH was designed in late 2020 and was predicated on:

- A high risk of SARS-CoV-2 transmission and COVID-19 disease in care homes.
- Treatment of care home residents who have been potentially exposed to SARS-CoV-2 due to an index case with an antiviral agent (post-exposure prophylaxis) might reduce viral spread and disease severity.
- There were no proven antiviral agents to treat or prevent SARS-CoV-2.

Hence, post exposure prophylaxis with an antiviral agent might reduce transmission within care homes and disease severity in residents.

## Chapter 2: Methods

### 2.1. Aims and objectives

#### 2.1.1 Aim

To set in place a research and governance platform for the efficient delivery of a suite of randomised comparisons to prevent COVID-19 infection and reduce severity/transmission and death in residents in care homes.

#### 2.1.2 Primary objective

To provide reliable estimates of the effect of trial treatments for each pairwise comparison with the standard care arm on SARS-CoV-2 infection, morbidity and mortality 60 days after randomisation.

#### 2.1.3 Secondary objectives

To assess the effects of trial treatments on mortality (all-cause and cause-specific), admission to hospital (all-cause and cause-specific), healthcare referrals for COVID-19, infection (asymptomatic, symptomatic), time to symptomatic infection and safety through serious adverse reactions.

To assess the effects of trial treatments on transmission of SARS-CoV-2 infection.

#### 2.1.4 Tertiary objectives

To assess the cost-effectiveness of trial treatments and explain the contextual factors which influence trial processes including adherence to intervention and outcome measurement regimens, and which might impact on subsequent implementations of pre- or post-exposure prophylaxis for COVID-19 in care homes.

### 2.2. Design

Cluster-randomised open-label blinded endpoint platform trial testing antiviral agents in a post-exposure prophylaxis paradigm.

The first fully approved protocol (version 1) is available at:

https://www.protect-trial.net/files/documents/protect-ch-protocol-final-v1-0-13-may-2021-1.pdf

The second fully approved protocol (version 2) is available at:

https://www.protect-trial.net/files/resources/protocol-final-v2-0-01-jul-2021_for-website-corrected-1.pdf

Further information is given at: https://fundingawards.nihr.ac.uk/award/NIHR133443

### 2.3. Study setting

The trial was due to be carried out in care homes in all four UK home nations (England, Northern Ireland, Scotland, Wales).

### 2.4. Participants

Adult residents of age 65 years or more were eligible for inclusion if they were a resident at a UK care home. To participate, residents would have to give written consent, or written proxy consent was to be obtained from a personal legal representative or relative if the resident lacked capacity; the consent form and patient information sheet are available on the trial website. The full eligibility criteria are:

#### 2.4.1. Care Home inclusion/exclusion criteria

##### Care home criteria at trial entry

***Inclusion criteria***

- Location: UK care homes for older people, with and without nursing.
- Size: ≥20 beds in the care home in total.

***Exclusion criteria***

- Care Quality Commission quality rating as Inadequate, or equivalent in devolved administrations.

##### Care Home criteria at treatment phase

***Exclusion criteria***

- Positive polymerase chain reaction or lateral flow test (or equivalent) for SARS-CoV-2 in any resident and/or staff within previous 4 weeks

#### 2.4.2. Resident inclusion/exclusion criteria

##### Resident criteria at trial entry

***Inclusion criteria***

- Resident in a Care Home.
- Age ≥65 years
- Able to give informed consent for participation or a personal legal representative has been identified who can give consent if resident lacks capacity.

*Exclusion criteria:*

- Identified by care home staff to have entered end-stage palliative care.
- Resident in care home for short-term respite care.
- Resident’s general practitioner is unable to support their involvement in the trial.

##### Resident criteria at treatment phase

*Exclusion criteria:*

- Currently taking all of the trial interventions.
- Contraindication to all trial interventions - see protocol’s IMP appendix.
- In treatment phase of another COVID-19 prevention or treatment trial

### 2.5. Data collected at baseline

Baseline data relate to those relevant to care homes and separately to residents.

#### Care homes

- Care home type (residential vs nursing vs nursing and residential)
- Prior COVID-19 in care home at any time (yes vs no)
- Size of Care Home - Total number of residents in care home (small (<30 residents), medium (>30, <50 residents), large (>50 residents))
- Care home has capacity to give oxygen and/or dexamethasone (yes vs no)

#### Residents

- Demographics: age, sex and ethnicity
- Medical history from care home records

### 2.6. Interventions

The NIHR Prophylaxis Oversight Group recommended in February 2021 that PROTECT-CH should test two drugs, inhaled ciclesonide and intranasal niclosamide.

#### Ciclesonide

Inhaled ciclesonide is a licensed drug for asthma prophylaxis ^31–33^ that has demonstrated anti-SARS-CoV-2 activity in vitro ^7, 8^ and observational clinical data relating to activity in COVID-19.^10–12^ Although still unpublished, the COVIS Pharma Group announced the top-line results of a double-blind placebo-controlled phase III trial of inhaled ciclesonide (320 µg bd); the trial studied 400 non-hospitalised patients with symptomatic SARS-CoV-2 infection. Although ciclesonide did not alter the primary outcome - time to alleviation of COVID-19 symptoms, treatment was associated with reduced visits to the emergency department or hospitalisation (9% vs 30%, p=0.030) (https://www.ema.europa.eu/en/news/insufficient-data-use-inhaled-corticosteroids-treat-covid-19; accessed 1 October 2021).

Although licensed in the UK, the version of inhaled ciclesonide made available to PROTECT-CH was an unlicensed version manufactured by Ayrtons, a UK specialist pharmaceutical company. Ciclesonide will be tested in an ongoing UK trial (PROphylaxis for paTiEnts at risk of COVID-19 infecTion; PROTECT-V), (https://www.camcovidtrials.net/trials/view,protectv_50.htm; accessed 1 October 2021).

Ciclesonide was to be prescribed once daily for 42 days, administered as follows:

- Two actuations (320 µg) inhaled via mouth sequentially. Participants who are unable to tolerate a face mask would use the spacer mouthpiece taking two actuations.
- One actuation (160 µg) inhaled via nose. Participants who are unable to tolerate a face mask would not receive the intranasal actuation.

#### Niclosamide

Oral niclosamide is approved for treating tapeworm infestations and as a general piscicide in aquaculture. Niclosamide has demonstrated anti-SARS-CoV-2 activity in vitro ^7^ and in vivo ^34^ and safety and tolerability data in a phase I trial in normal volunteers.^35^

Although not licensed in the UK, the British National Formulary describes oral niclosamide as the most widely used drug for tapeworm infection (https://bnf.nice.org.uk/treatment-summary/helminth-infections.html; accessed 1 October 2021). Inhaled niclosamide is an unlicensed formulation being developed by the Danish company, Union Therapeutics. A trial of inhaled niclosamide is ongoing for the prevention of COVID-19 in patients with renal disease (haemodialysis, renal transplant, inflammatory renal diseases). Niclosamide is being assessed in the PROTECT-V trial and has recruited 350 of a planned 1,500 patients to date from 23 sites (https://www.camcovidtrials.net/trials/view,protectv_50.htm; accessed 1 October 2021). Niclosamide (1% in 20 ml) was to be administered for 42 days intranasally 140 µL spray into each nostril twice daily (equivalent to a total daily dose of 4.7 mg of niclosamide free acid).

#### Control group

Both ciclesonide and niclosamide were to be given in addition to standard care and compared with standard care alone, i.e. there was to be no placebo.

### 2.7. Randomisation

As a cluster-randomised trial, care homes were to be randomised dynamically using a probabilistic minimisation algorithm to balance across important baseline care home characteristics: type (residential vs mixed residential and nursing vs nursing), prior SARS-CoV-2 infection at any time, size (<30 residents, 31-50, >50), capacity to give oxygen and/or dexamethasone. The probability of allocating to the group that minimised the imbalance was 90%. Eligible nursing homes were to be assigned in a 1:1:1 ratio to receive ciclesonide and standard care, niclosamide and standard care, or standard care alone. Residents who had a definite need for or contraindication to either drug were not included in analyses of that comparison with standard care. Residents, care home staff and general practitioners would be aware of the assigned treatments i.e. allocation concealment was to be ensured by enrolling care homes and residents prior to allocation. Although the trial was open-label and so residents and treating staff were to be unblinded, the primary outcome was to be blinded being derived from national routinely collected health data.

Care homes were to be randomised once they had an indication of a developing infection, e.g. recent positive PCR or lateral flow test (or equivalent) in any resident or member of staff (index case). Care homes were to be randomised to ciclesonide vs niclosamide vs control.

### 2.8. Assessments before and immediately after randomisation

Assessments were to be performed both before and immediately after randomisation:

#### Post consent/pre-randomisation

- Eligibility screen
- Demographics and medical history
- Quality of Life: EQ-5D-5L; EQ-VAS
- Number of residents in care home

#### Day 0/immediately post-randomisation

- Number of residents in care home.
- Source of infection of SARS-CoV-2 (resident, staff), date and type of test (lateral flow, PCR or equivalent).

### 2.9. Outcomes

#### 2.9.1. Primary efficacy outcome

The primary efficacy outcome was to be a four-level ordered categorical (ordinal) scale with participants classified according to the highest level, that is, the most serious event they experienced during the 60-day period following randomisation:

1. No SARS-CoV-2 infection.
2. SARS-CoV-2 infection (diagnosed using PCR, lateral flow test or equivalent) without admission to hospital.
3. Admission to hospital, all-cause.
4. Death, all-cause.

Outcomes were to be assessed at 60 (and 120 days in a secondary analysis) following randomisation and information on events would be obtained from UK routine data, with national sources shown in Table 2.

**Table 2.** Sources of routine national data by UK nation.

| Home nation | England | Northern Ireland | Scotland | Wales |
| --- | --- | --- | --- | --- |
| Death | NHS Digital | Health & Social Care | Public Health | Secure Anonymised |
|  |  | Trusts NI (HSCNI) | Scotland (PHS) | Information Linkage (SAIL) |
| Hospitalisation | NHS Digital | Health & Social Care | Public Health | Secure Anonymised |
|  |  | Trusts NI (HSCNI) | Scotland (PHS) | Information Linkage (SAIL) |
| SARS-CoV-2 | NHS Digital | Health & Social Care | Public Health | Secure Anonymised |
|  |  | Trusts NI (HSCNI) | Scotland (PHS) | Information Linkage (SAIL) |
| Primary care | NHS Digital | Health & Social Care | Albasoft | Secure Anonymised |
|  |  | Trusts NI (HSCNI) |  | Information Linkage (SAIL) |
| Other | - | Honest Brokerage | - | - |
|  |  | Service NI (HSBNI) |  |  |

##### Choice of the primary efficacy outcome

The primary efficacy outcome was designed to “capture the ability of the drug candidate to prevent/reduce morbidity and mortality from COVID-19 in individuals, and to reduce transmission in care home settings” thereby addressing the NIHR Commissioning brief (https://www.nihr.ac.uk/documents/20111-commissioning-brief-for-prophylaxis-platform-study-in-care-homes/25902; accessed 26 October 2020). Hence, the outcome needed to include asymptomatic and symptomatic transmission, morbidity as healthcare interventions and hospitalisation, and mortality. This approach follows the WHO recommendation to use ordinal outcomes in COVID-19 trials,^36^ a recommendation that has been followed by many COVID-19 trials.^37^ We adapted the ordinal scale to fit the care home context.

Using an ordinal outcome had additional advantages. First, it allowed the effect of treatment to be assessed on the severity of recurrent events as well as their rate. In general, interventions that reduce the risk of events also reduce the severity of those events that do occur;^38, 39^ conversely, interventions that increase events also increase the severity of those events.^40^ Second, using an ordinal outcome improves statistical power compared to using a dichotomous outcome for a given sample size.

#### 2.9.2. Secondary efficacy outcomes

##### Secondary outcomes at day 60 following randomisation (unless otherwise stated)

- Time to healthcare referral for COVID-19, e.g., discussion outside of care home with GP (excluding routine visit), 111, 999 /ambulance paramedic or Emergency Department assessment (without admission), remote hospital consultation
- Time to use of dexamethasone in the care home for COVID-19
- Time to use of oxygen in the care home for COVID-19
- Time to SARS-CoV-2 infection - positive PCR or lateral flow test (or equivalent)

`o` with symptoms of COVID-19
`o` without symptoms of COVID-19
`o` total, i.e. either with or without symptoms of COVID-19
- Time to first admission to hospital
- Cause-specific hospital admission
- Time to death
- Days alive and not in hospital
- Cause-specific mortality, including COVID-19, stroke, pulmonary embolism, myocardial infarction
- Electronic frailty index at 60 days

##### Secondary outcome at day 120 following randomisation

Ordinal outcome for the most serious event experienced during the 120 days post-randomisation with the following levels:

1. No SARS-CoV-2 infection
2. SARS-CoV-2 infection but resident remains in care home
3. Admission to hospital, all-cause
4. Death, all-cause.

##### Clinical – care home level

- Number of SARS-CoV-2 infections in residents in the care home (aggregate data including residents not participating in PROTECT)

##### Economic evaluation

- EQ-5D-5L utilities and EQ-VAS at 60+/-2 days
- Quality Adjusted Life Years (QALY)
- Healthcare resource use and costs
- Incremental cost-per QALY and Net Monetary Benefit

#### 2.9.3. Safety outcomes

- Serious Adverse Reactions (SAR, excluding primary and secondary outcomes) and Suspected Unexpected SARs (SUSARs)
- Adverse events relevant to the intervention (see relevant IMP-specific Appendix)

Care homes were to report SAEs and AEs and the planned process for this is shown in Figure 2.

**Figure 2.**
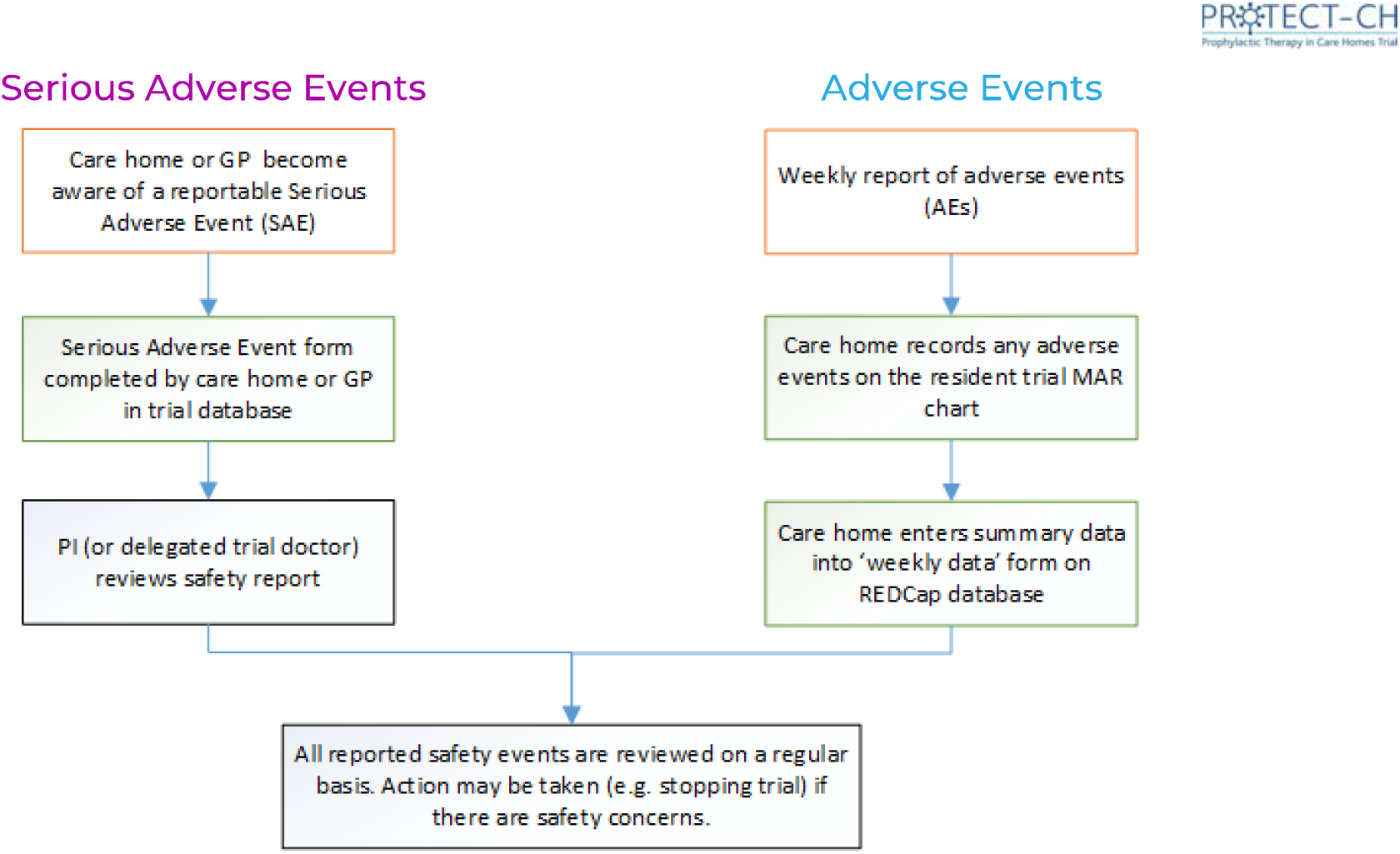
Process for reporting serious adverse events and adverse events.

### 2.10. Study oversight

The trial was conceived and designed by the grant applicants who wrote the protocol. The study was given NIHR Urgent Public Health level 1 badging (dated 1 March 2021); approved by the Medicines and Healthcare products Regulatory Agency (MHRA, UK competent authority, CTA 03057/0073/001-0001, 15 May 2021), UK research ethics committee (REC) and Health Research Authority (HRA, 21/SC/0166, dated 17 May 2021); and registered (EudraCT 2021-000185-15).

The trial was overseen by independent Platform Steering (PSC) and Data Monitoring Committees (DMC), Platform Management Committee (comprising the grant applicants) and the Platform Management Group. The day-to-day conduct of the trial was run by an Executive Committee based at the PROTECT-CH coordinating centre at the Nottingham Clinical Trials Unit. Membership of the committees/groups is listed at the end of the report.

#### This report/monograph

Report writing was performed independent of the funders, sponsor and pharmaceutical companies. The corresponding author wrote the first draft of this report; this and subsequent drafts were edited by the grant applicants, who all approved the decision to submit the manuscript for publication. The corresponding author had final responsibility for the decision to submit for publication and is the guarantor for the study.

#### Platform Steering Committee (PSC)

The Platform Steering Committee (PSC) comprised 9 independent and 1 non-independent members and was to provide oversight for the entire project. It would have received information as to the progress of the project against pre-determined milestones from the Platform Management Group and information on safety and efficacy from the Data Monitoring Committee (DMC). The PSC were to follow a pre-defined charter and were charged with providing independent oversight of the project. The PSC charter is available on the trial website.

The PSC supported the plan to close the trial and approved the close down plan.

#### Data Monitoring Committee (DMC)

The independent DMC comprised five independent members and was to review unblinded data in confidence regularly. The DMC was responsible for safeguarding the interests of trial participants, assessing the safety and efficacy of the intervention during the trial, assessing data integrity, and for monitoring the overall conduct of the trial. The DMC were to follow a pre-defined charter and were charged with informing the PSC if, at any time, the data showed evidence beyond reasonable doubt of a difference between the randomised groups in the primary outcome. They were also to consider data in the light of external information such as results from completed trials. The DMC charter is available on the trial website (<u>protect-trial.net)</u>.

The DMC supported the plan to close the trial and approved the close down plan.

### 2.11. Statistical analyses

The analysis and reporting of the trial were to be in accordance with CONSORT guidelines for adaptive and cluster designs ^41, 42^ with the primary comparative analyses being conducted according to randomised allocation (intention-to-treat). All comparative analyses were to be based on contemporaneously randomised care homes. Primary comparative analyses would have employed a multi-level ordinal logistic regression model with adjustment for the minimisation factors and individual-level covariates (age, sex, vaccination status) and a random effect to adjust for clustering within care homes. The treatment comparison would have been presented as an adjusted common odds ratio (with 95% confidence intervals) for a shift in the direction of a better outcome on the ordinal scale.^38, 39, 43–45^ Prespecified analyses of the primary outcome were to be performed in subgroups defined by the adjustment factors: care home type, prior SARS-CoV-2 infection in the care home, number of residents in care home, capacity to give oxygen, age, sex, vaccination status. Secondary outcomes were to be analysed using appropriate regression models dependent on data type (binary, categorical, continuous, time to event), adjusted similarly and accounting for clustering within care homes. All P values would have been two-sided and reported without adjustment for multiple testing. Analyses were to be performed using Stata and R.

The full statistical analysis plan, as approved by the Platform Steering Committee (PSC), is appended in Appendix 1.

#### 2.11.1. Sample size

A total of 530 residents per group were required to detect an odds ratio of 0.67 for a 4-level ordinal primary outcome (no infection 60%, infection and remain in care home 15%, all-cause hospitalisation 10%, all-cause mortality 15%), assuming a two-sided significance level of 5% and 90% statistical power, with no adjustment for clustering.^46, 47^ Care homes of varying size were to be included with an average of 40 beds per care home;^48^ we assumed that not all residents would take part in the study and so expected that approximately 32 (range 20-60, coefficient of variation for care home size 0.49 ^49^) residents would be recruited from each care home. Assuming an intra-cluster correlation of 0.11 gave a design effect or inflation factor of 5.25.^50^ Therefore, to compare a single active treatment versus standard care, we would need around 174 care homes and in excess of 5,500 residents. Allowing for the uncertainty surrounding the parameters listed above, we proposed a sample size of 200 care homes involving 6,400 residents.

Therefore, a comparison of two active (unrelated) treatments versus standard care (in a 1:1:1 allocation ratio) would require 300 care homes in total, corresponding to around 9,600 residents. Since only ∼40% of care homes might go positive during the trial, we would need to recruit 2.5 times these numbers, i.e., 750 homes and 24,000 residents, and then randomise the first 300 care homes that reported an infection. We would then re-estimate the sample size during the trial once 60-day outcome data were available for at least 75% of care homes randomised to standard care.

#### 2.11.2. Health economics

The primary economic evaluation planned was a within trial cost-utility analysis based on outcomes at day 60, with an NHS cost perspective. Healthcare resource use collection was designed to be parsimonious and feasible and would be collected by eCRF in addition to routine data sources. Resource use data collection was to include primary care contacts, use of ambulance services and secondary care attendance or stays. Health related quality of life measured using the EQ-5D-5L and EQ-VAS (proxy report) at 60 days was to be measured and EQ-5D-5L presented descriptively in addition to estimating between group differences. Self-reported HRQoL was also to be collected where possible. Self-report and proxy report response patterns were to be explored and reporting subgroups (self-report vs proxy) examined in sensitivity analysis.

Statistical analyses were to be conducted in line with other outcomes, using linear mixed effects models, additionally controlling for differences in baseline EQ5D utility, accounting for potential non-normality in economic data and correlation between cost and QALYs, where appropriate. Missing data was to be assessed and handled appropriately depending on the nature of the missingness.

Incremental costs (including any potential savings) associated with prophylaxis for care-home residents were to be estimated. EQ-5D-5L was to be used to compute Quality Adjusted Life Years (QALYs) and estimate incremental QALYs. Costs and QALYs were to be combined to estimate the incremental cost-effectiveness ratio (ICER), and present incremental net monetary benefit (INMB) at various willingness to pay thresholds. Uncertainty would be characterised using bootstrap sampling and cost-effectiveness acceptability curves. A secondary analysis would be performed based on outcomes at 120 days (survival and resource use from routine sources).

The full health economics analysis plan, as approved by the Platform Steering Committee, is appended in Appendix 2.

#### 2.11.3. Protecting against bias, including blinding

Multiple measures were to be taken to minimise bias: recruitment according to pre-defined inclusion/exclusion criteria, with exclusion of patients enrolled in other trials; central data registration with real-time data validation and concealment of allocation; research staff trained in trial protocol and processes; analysis by intention-to-treat; analyses adjusted for baseline prognostic variables, including minimisation factors.

### 2.12. Sites, investigators, and monitoring

#### Training of Investigators

All PROTECT-CH care home staff were to be trained in the trial protocol and processes, and assessment scales. The training included an introduction to Good Clinical Practice with increased detail covering aspects relevant to staff, e.g. consent and IMP management. (More information on this follows.)

#### Schedule for Monitoring of Sites and Data Integrity

Care home monitoring was to be performed remotely by the NCTU Coordinating Centre with the aim of ensuring quality control of the delivery of the protocol, collection of data, and adherence with national regulations and ethics. Remote monitoring to confirm the presence of the participant and their consent, eligibility criteria, selected data critical to the trial (demographics, prescription of interventions) and report serious adverse events were planned. In-person monitoring visits were to be performed as deemed necessary by the Coordinating Centre.

Central statistical monitoring of the data was to be performed according to Buyse *et al*^51^ during the trial and prior to locking of the data. Checks would include logic and range checks, digit preference, comparison of univariate data between sites and comparison of multiple variable models between countries. The monitoring procedures would have been compliant with the requirements of the sponsor, the national ethics committees and MHRA and fulfilled Good Clinical Practice requirements.

### 2.13. Sub-studies

#### Process evaluation

A nested process evaluation was designed to run concurrently with the trial. The evaluation was informed by a realist approach ^52–55^ with the following objectives:

- To provide contextualised insight into the delivery of the intervention(s);
- To consider acceptability of the intervention to staff, residents, and their families;
- To reflect upon and inform trial processes.

It was proposed that 10-20 care homes would be purposively selected to test programme theories which describe how trial design and intervention delivery were intended to work. Homes would be selected to reflect contexts where different mechanisms might be triggered by the introduction of prophylactic measures and trial procedures. Factors in this might be size of care home, home ownership, the presence of nursing staff, proportions of residents living with dementia, and/or prior experience of COVID-19 (deaths/hospitalisations).

In each setting care home records and trial data were to be reviewed. Interviews were to be undertaken with care home and healthcare professionals. Where possible residents and their families would be interviewed. Interviews would consider both the appropriateness of prophylactic intervention in care homes as well as detail about local experiences of intervention. To inform the on-going delivery of the platform trial interviews would additionally seek insight about the local experience of trial procedures.

The focus of analysis in realist evaluation is the iterative development of the initial programme theories (Figure 3). This is done through the conceptual lens of Context-Mechanism-Outcome (C-M-O) configurations. Here thematic analysis of interview data (M) mapped against local contextual features (C) and Covid cases (O).

**Figure 3.**
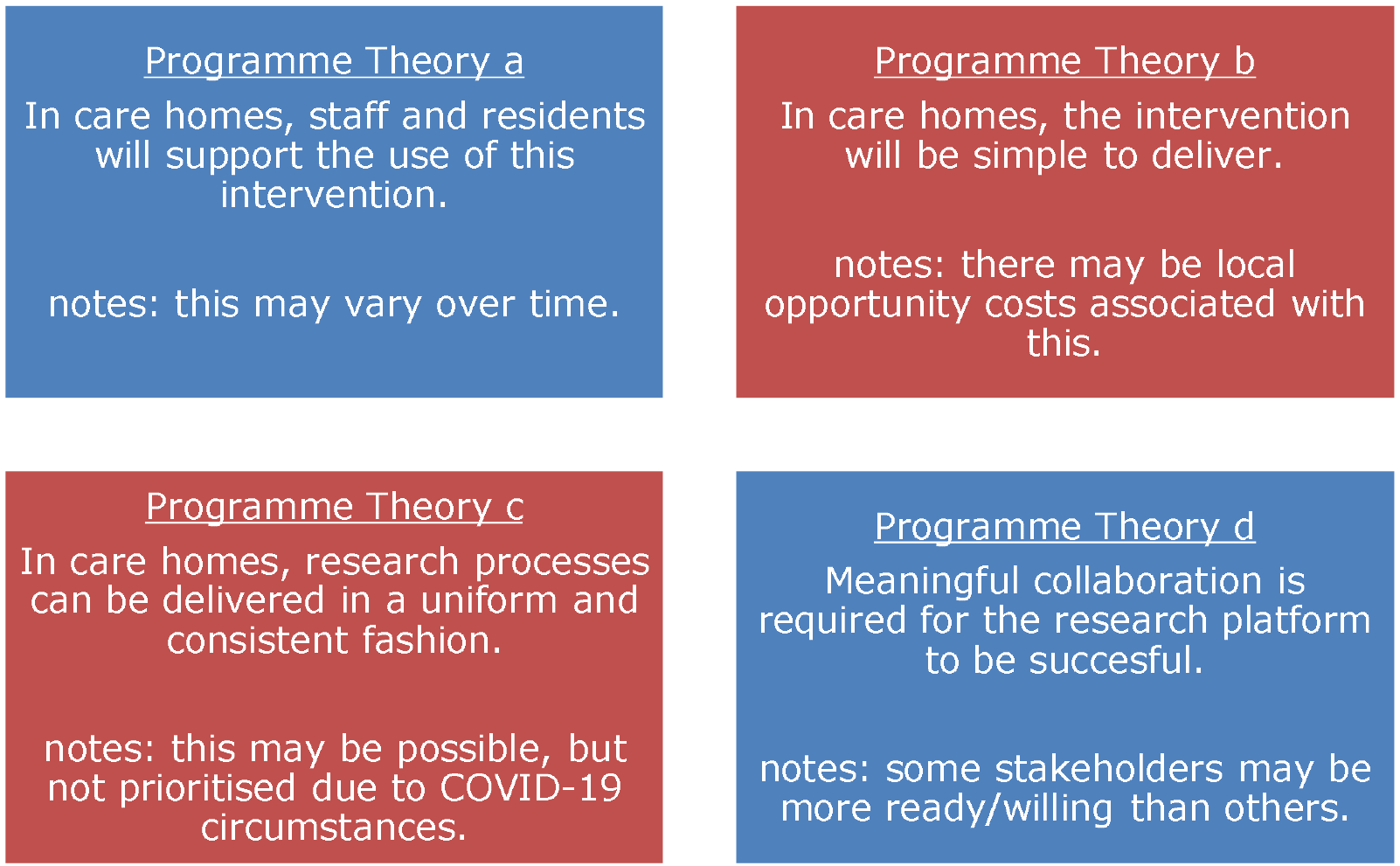
Initial (simple) Programme Theories.

Recurrent patterns in these C-M-O configurations provide insight about where, and why, measures are readily adopted and have positive impact.

### 2.14. Sharing

Anonymised subsets of individual patient data were to be shared with other research groups and projects (e.g., Cochrane Collaboration; Virtual International care homes Trials Archive [held in Glasgow, Robertson Institute]) once the main and secondary trial findings had been published and following agreement by the Platform Steering Committee. A contract would be set up between the University of Nottingham (as represented by the Chief Investigator) and groups that were to receive data.

### 2.15. Pre-start-up activities

#### Principal investigators (PIs)

With testing of unlicensed IMPs, it was not possible for care home managers to be the principal investigator at each site; this contrasts with our previous FiCH,^56^ FinCH ^57^ and BEET-Winter trials.^58^ Instead, we set up a network of 35 medics across the UK comprising some of the applicants as well as academic and NHS consultants and GPs. PIs would have a portfolio of 10-30 care homes within their geographical region to supervise. Their roles and responsibilities are shown in Figure 4. PIs were to be trained via the website-mounted slide deck (see section on training). To support PI development, fortnightly online meetings were set up and hosted by the study team throughout the lifespan of the project.

**Figure 4.**
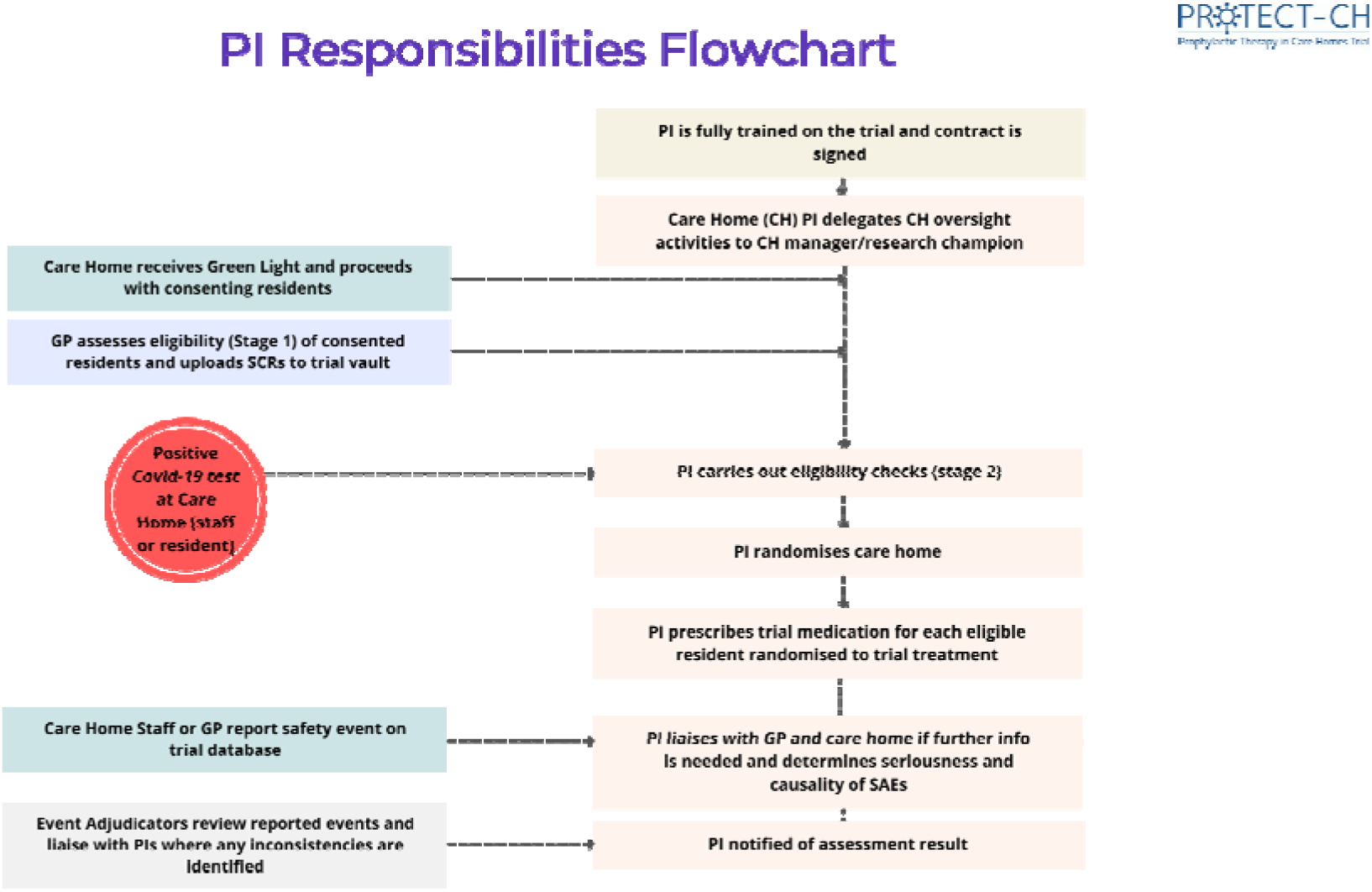
Principal investigator roles and responsibility.

#### Research nurses (RNs)

External qualified nurses would have been required to assist with the trial delivery:

1. To train care home staff and residents on how to administer unlicensed IMP
2. Residential care homes have few if any qualified nurses, and nursing homes may have insufficient numbers.

Research nurses were to be recruited from two sources, those aligned with the principal investigators (whose time was to be paid for from the grant), and those working with the Clinical Research Network (CRN), including EnRICH (England), EnRICH Scotland(Scotland) and EnRICH Cymru (Wales); the costs for the latter would come from service support costs, or equivalents. Their main roles would be to lead on consent, help administer IMP on the first day of administration and general support for enter data into REDCap, especially related to SAE reporting (Figure 5). Overall, we estimated that the trial would have needed 30 research nurses evenly spread across the UK. Discussions with CRN suggested that finding these numbers would be challenging especially without knowing which care homes had signed up. As with PIs, it was unclear in some organisations as to whether they were willing to contract for RNs.

**Figure 5.**
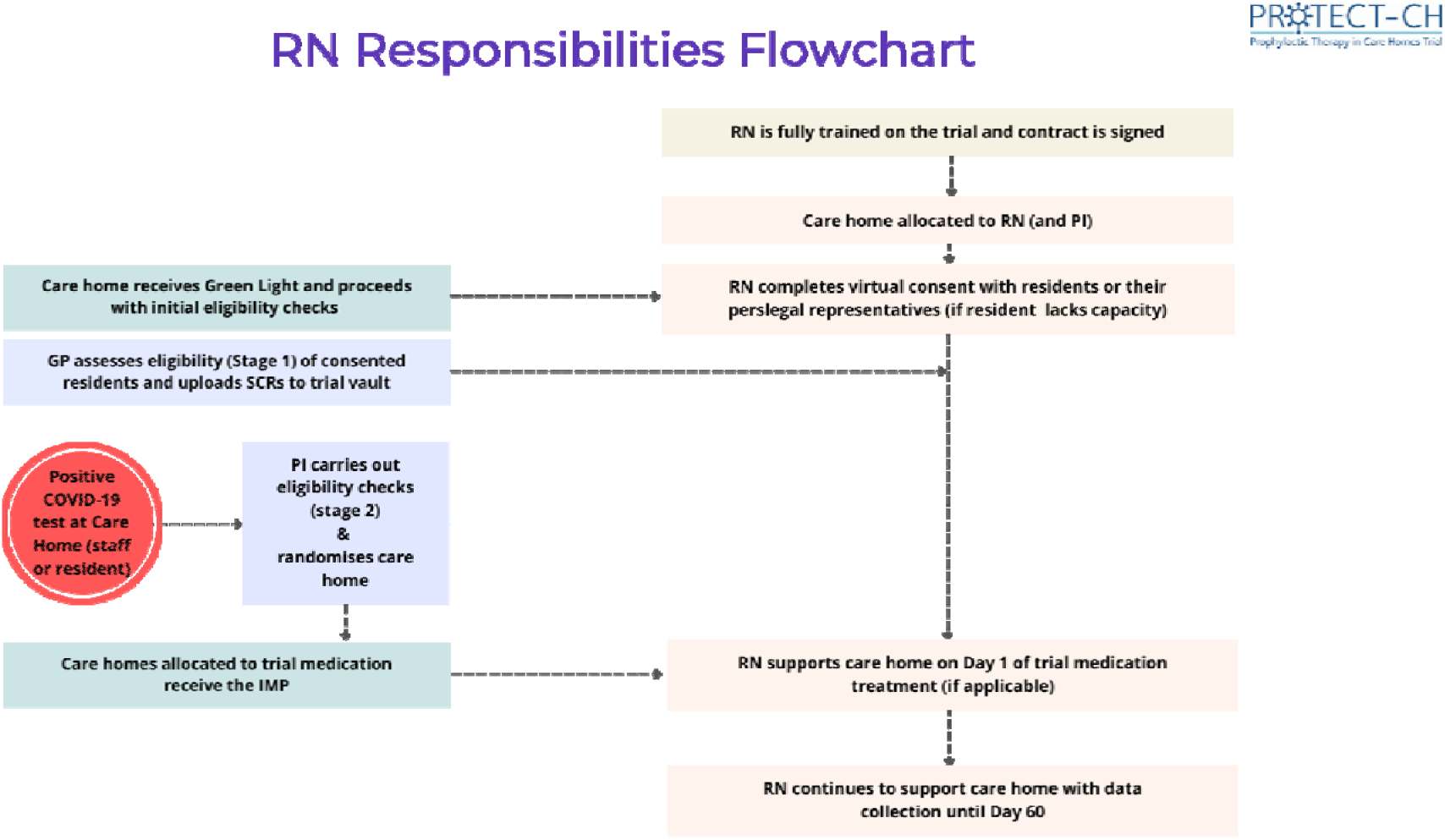
Research nurse roles and responsibility.

#### General practitioners (GPs)

Although not planned at start, MHRA requirements for the active involvement by resident’s GPs meant we would have had to identify and then contract individually with them. With approximately 4 (range 1-10) GPs working in each care home, we would need to contract with approximately 3000 GPs, a massive undertaking. Our PMC, which included two GP members (BG, SR), decided that GPs should be involved at two stages:

1. To provide information on the residents’ eligibility and relevant history. The most straight-forward means of this would be via GPs sharing the summary care record for each resident with this submitted to a secure database.
2. To assist with reporting serious adverse events if they were involved with managing the event. GPs were to be trained via the website mounted slide deck (see section on training). GPs are aligned with individual residents so without knowing which residents were going to sign up, it was not possible to start approaching GPs formally; in reality, only four signed up based on the limited information that we could share with them although contracting was not initiated (Figure 6).

**Figure 6.**
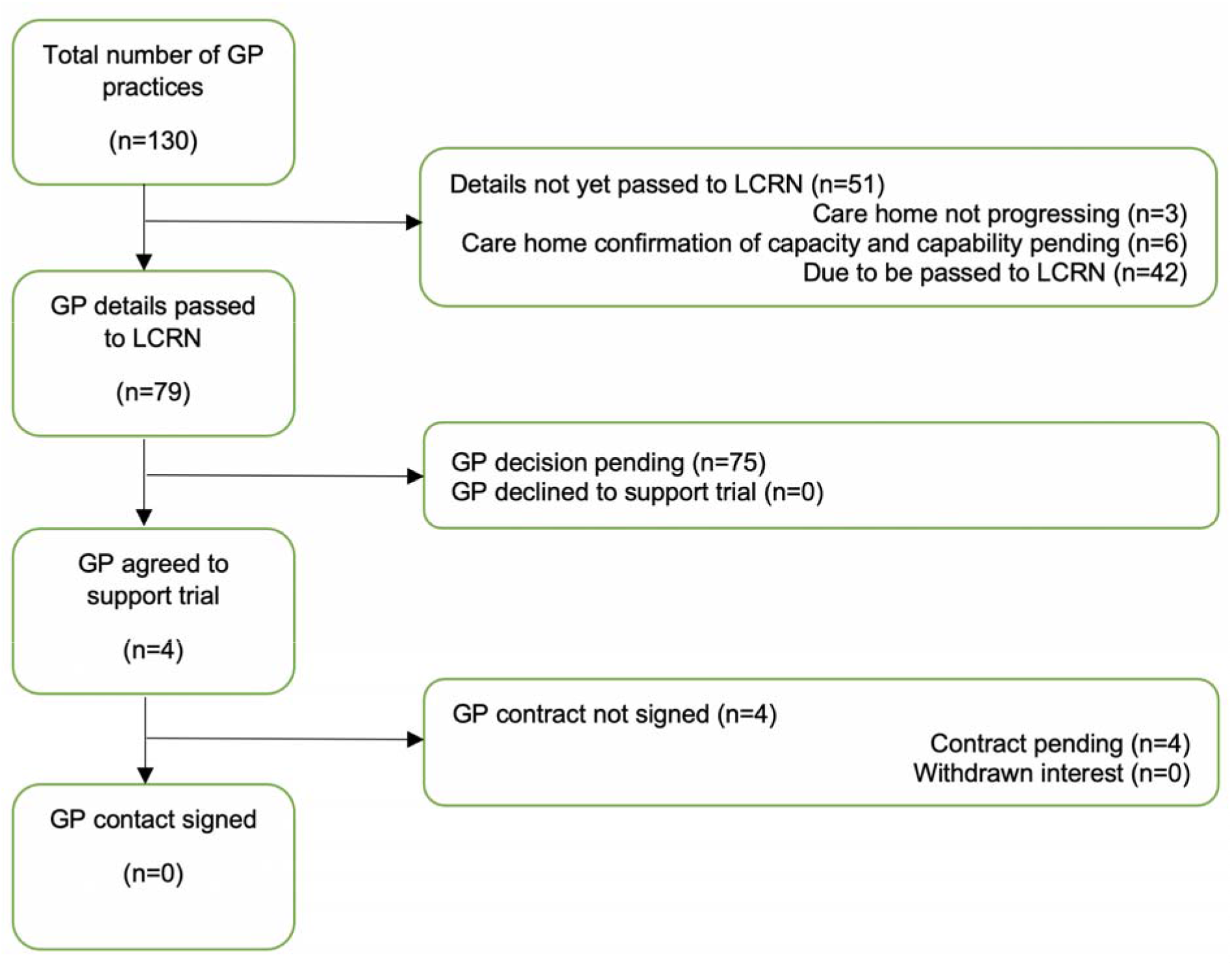
Status of GP recruitment and contracting.

#### Care homes

Since there is no easily accessible central register of care homes, we identified these using multiple ascertainment mechanisms:

1. Potential care homes were identified from emailing and social media lists and those that had participated in our earlier COVID-SEARCH, FiCH, FinCH and BEET-Winter trials. These were emailed with information about the trial and invited to express interest via a REDCap database. Of 518 care homes who submitted an EoI, 308 completed the form (England 268, Northern Ireland 3, Scotland 16, Wales 12) although not all of these were eligible (figure 7).
2. The chief executives or other board members of large care home chains were approached directly and invited to join briefing video-conference calls.
3. The chief executives of national care home membership organisations (National Care Forum for not-for-profit homes, Care England for corporate providers, Care Scotland, and the Care Providers Alliance as the umbrella organisation of care home providers in England) were approached and asked to promote the trial among their membership.
4. The chief executive of the Care Quality Commission (CQC), the independent regulator of health & social care in England, was approached and asked to promote the trial among the care homes they inspect. Information about the trial was included in the CQC digital newsletter for care homes.
5. Via the Clinical Research Network (CRN) and, through this, the national Enabling Research in Care Homes (EnRICH) networks for care homes known to them.
6. Media (Appendix 5) and social media coverage of the trial led to approaches by some medium-sized care home chains; again, these were invited to join a briefing video-conference call.

**Figure 7.**
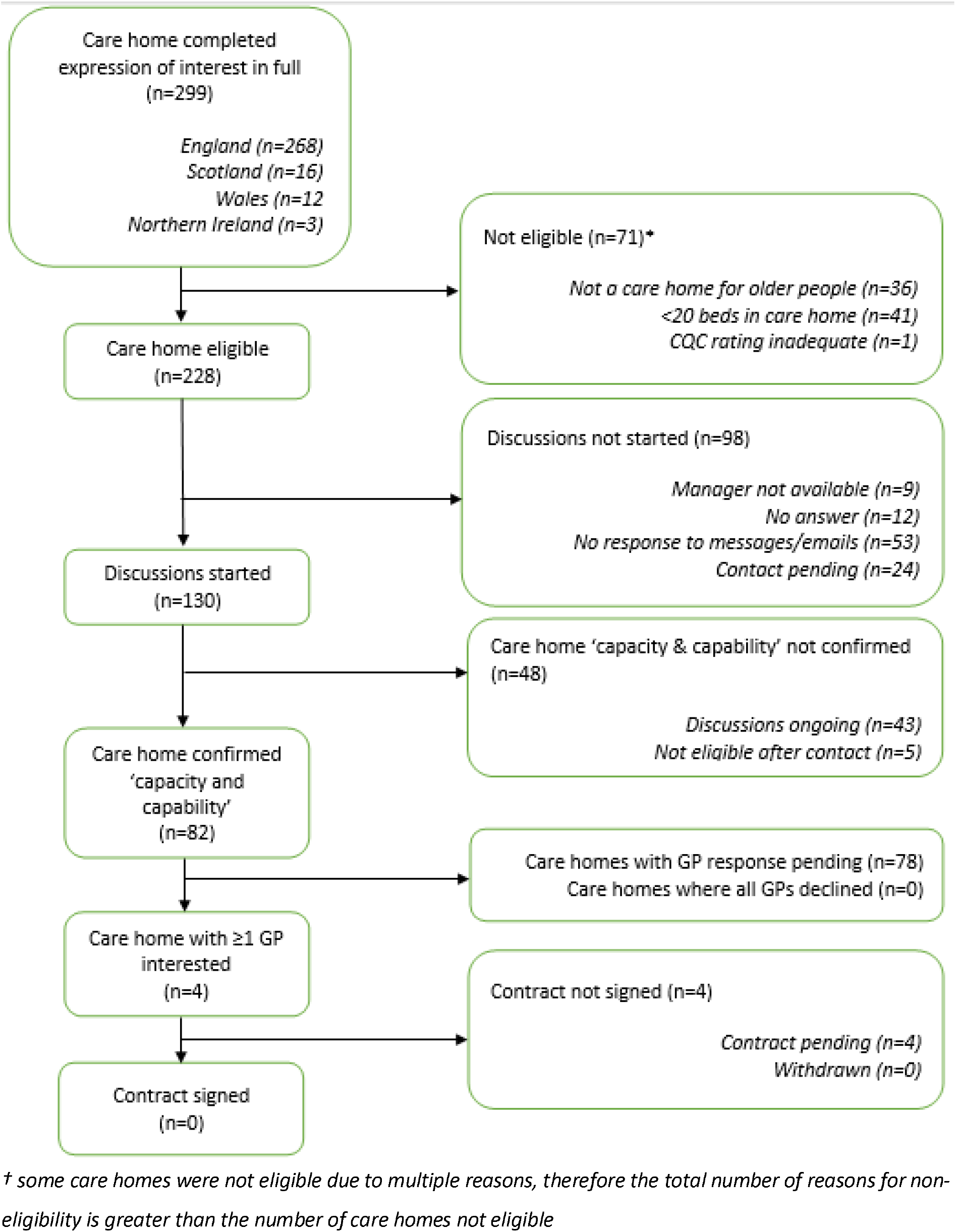
Status of care home eligibility and contracting.

Of solo, small and medium sized care homes chains, 228 fulfilled the platform inclusion criteria and were contacted via email and of these, 130 had follow-up discussions and 82 confirmed their capacity to deliver the trial. Of these 82 care homes, confirmation of GP support was obtained via the LCRN for 4 care homes, and these were deemed ready to initiate contracting by the time the decision was made to close the platform. Two of the large care homes chains were interested potentially in joining. In all cases, the inability to tell the care homes what the interventions were limited further progression.

To maintain care home engagement and build relations as the basis for conducting the trial, we planned and co-ordinated fortnightly 1-hour-duration educational workshops (Appendix 6), conducted online through Microsoft Teams. The first half of each event comprised study updates and an opportunity to consult care home providers on design issues with the protocol or study materials. The second part included a series of talks by invited expert speakers, pitched at care home staff and based on their suggestions for content. A programme for these events is included as appendix 3, and recordings of the sessions remain available via our study website (permission required for access).

#### Insurance

Conventionally, insurance can be categorised as that covering the:

- Sponsor, host institution and their staff, and the protocol. The University has trials insurance to cover this for most trials, but this was not designed to cover a large platform trial such as PROTECT-CH. Hence, UoN would need to purchase additional cover through the grant for the platform.
- Recruiting sites, in this case care homes and their staff for non-negligent events. Unlike NHS hospitals and general practices, the majority of care home insurance policies do not cover research. As a result, UoN would need to purchase additional cover through the grant for care homes.
- Participating healthcare staff (medics, nurses) for negligent procedures. Discussions with medical defence/protection societies/unions suggested this cover would extend to PROTECT-CH at no extra cost.

At the time we commenced the study, insurance costs for care home providers had increased substantially and many providers had found stringent conditions attached to new or existing policies.^60^ Insurers of the sector were particularly risk aware given the events of the first wave of the pandemic.

Discussions with several insurance companies providing routine cover for care homes suggested that none were willing to extend this to cover research. Primary drivers for this position included the:

- Perceived risk of doing research in research-naïve sites.
- Perceived risk of a trial testing unlicensed drugs.
- Uncertain risk associated with COVID-19 in care homes taking account of the high death rate in wave 1 of the pandemic and opening up of care homes to relatives and visitors with the increased risk of importing infection into the care home.

In the absence of commercial insurance, discussions were held with DHSC, and a fallback position was for Her Majesties Government to underwrite the trial as part of the Coronavirus Act 2020 (https://www.legislation.gov.uk/ukpga/2020/7/contents/enacted, accessed 3 October 2021). Subsequently, DHSC and the UoN Procurement Office separately identified that Lloyd’s, Brokered by Aon UK Ltd and underwritten by the Newline Syndicate (Table 3), could provide a single insurance covering UoN, care homes and healthcare staff, this covering:

> “all Bodily Injury resulting or alleged to have resulted from the same Trial shall be considered as resulting from one Occurrence and having occurred during that Period of Insurance in which the first Claim is made against the Insured irrespective of the number of claimants or the period over which such Bodily Injury is likely to result in a Claim or Claims being made against the Insured at some future date.”

**Table 3.** Suppliers of services for the trial.

| Activity | Description | Supplier |
| --- | --- | --- |
| Drug | Inhaled ciclesonide | Ayrtons (https://ayrtons.com) |
|  | Intranasal niclosamide | Union Therapeutics<br>(https://www.uniontherapeutics.com) |
| Training |  | In house |
| Website | Host trial materials | fry-it (https://fry-it.com) |
| Database |  | REDCap (https://www.project-redcap.org) |
| Insurance | Cover protocol, care homes and healthcare staff (medics, nurses) | Lloyd's. Broker: Aon UK Ltd<br>Underwriter Newline Syndicate<br>(https://newlinegroup.com/newline-syndicate-1218/) |
| Central pharmacy | Manufacture, QP, store, dispense, distribute drugs | BCM Specials (http://www.bcm-specials.co.uk) |
| Trusted Research Environment | Retain data from national data registries for the project | University of Dundee |
| Site management system | Create bespoke system to retain the site documentation and training | University of Aberdeen Centre for Healthcare Randomised Trials (CHaRT) |

Contracting for this insurance was commenced in July 2021 but not completed due to the platform closure.

#### Investigational medical products (IMP)

The NIHR Prophylaxis Oversight Group requested that the trial study two unlicensed drugs, inhaled ciclesonide (Ayrtons, UK) and intranasal niclosamide (Union Therapeutics, Denmark) (Table 3).

Both companies were approached and offered to provide the drug for free to the trial. Unfortunately, neither could agree to contracting terms compliant with the master contract between NIHR and UoN. Specific issues were the need for NIHR to obtain background IP from the company (in case the company could or would not develop the agent if effective) and refusal by NIHR to agree to the company using the final data for licensing approval and marketing support until after the results were available. Much time was lost in trying to resolve this difference. Eventually, Ayrtons agreed to sell their drug at cost price without any expectation of access to data. Contracting for this drug was never completed due to the platform closure. Fine detail on the agreement with Union was not obtained but was likely to involve sale rather than gift of drug.

A separate complication was the MHRA’s insistence that each resident’s general practitioner (GP) was involved taking account that the platform involved unlicensed IMP and the population were frail and would often lack capacity. With several GPs having residents in each care home, and many GPs having patients in more than one home, this would involve identifying, contracting and training several thousands of GPs. Further, some residents would not be able to enter the trial if their GP could or would not take part.

The issues with IMP contracting, in particular, set the study date back by four months.

#### Central Pharmacy

With the potential to test up to three drugs, it was vital that the manufacture, qualified person (QP), storage, dispensing and distribution were managed through a central pharmacy. For the planned interventions, this would need to be able to:

- Keep stock refrigerated.
- Dispense 7 days per week. In a PEP design, a care home developing a positive case on a Friday would need drugs arriving on Saturday rather than on the Monday (or Tuesday if a bank holiday week-end).

Thirteen pharmacies, both NHS and commercial, were approached and discussions held by videoconferencing and email. However, only one (BCM Specials, Table 3) was able to guarantee 7 day working, albeit at significant additional cost. Alternate weekly meetings were held with this company from May 2021 and contracting commenced but never completed due to the platform closure.

#### Databases and file storage

##### REDCap database

An early decision in trial start-up was to develop the primary database using the REDCap database environment (Table 3) rather than developing a bespoke system. The system was housed on NCTU servers and comprised development, test and live versions.

The component systems comprised: Care Home expression of interest survey (REDCap), Aberdeen CHaRT site management (bespoke), Care Home management including minimisation randomisation and delegation log (REDCap), PROTECT-CH trial database including all patient level data and functionality: Eligibility checks, eConsent, data collection, reports and data export to Dundee Trusted Research Environment (TRE) for merging with routine data (REDCap). Roles for logins were created within user groups in care homes, PIs and GPs, and surveys for PROMs and eConsent for Personal Legal Representative (PLR) for residents without the capacity to consent themselves.

Those parts of the database system that are not confidential have been uploaded to GitHub: https://github.com/Nottingham-CTU/PROTECT-CH-TRIAL

##### eTMF

As remote working was in force, an eTMF was set up using SharePoint document libraries to manage document history and version control, and Power Automate for document approval flows to replace ‘wet signing’ paper copies or separate emailed approvals. Managed term sets were also used for metadata tagging on documents to improve searchability within the eTMF.

##### Vault document repository

Files containing identifiable information including the summary care record and medicine administration record were stored in a bespoke encrypted database housed on a separate server to the REDCap database. The system was derived from one supporting the MRC ENOS, HTA TARDIS, HTA TICH-2 and BHF RIGHT-2 trials and has now subsequently been further enhanced to support current NIHR trials run from the Stroke Trials Unit at the University of Nottingham including RfPB DASH, EME RECAST-3, HTA MAPS-2, HTA TICH-3 and HTA PEAST.

##### Website

Initially, a website was set up on the UoN server but a preference for a professionally-hosted website led to a search for suitable host. FRY-IT (Table 3), who host the RECOVERY trial website (https://www.recoverytrial.net, accessed 3 October 2021), were approached and contracted. The initial specification was to host information about the trial, training materials and frequently asked questions but this website (https://www.protect-trial.net, accessed 16 February 2022) will now be used following close down as a public repository for all trial materials.

### 2.16. Trial procedures

Considerable time, effort and discussions were spent in designing the flow of residents into the trial and their treatment and follow-up, as shown in the following Figures 8, 9, 10.

**Figure 8.**
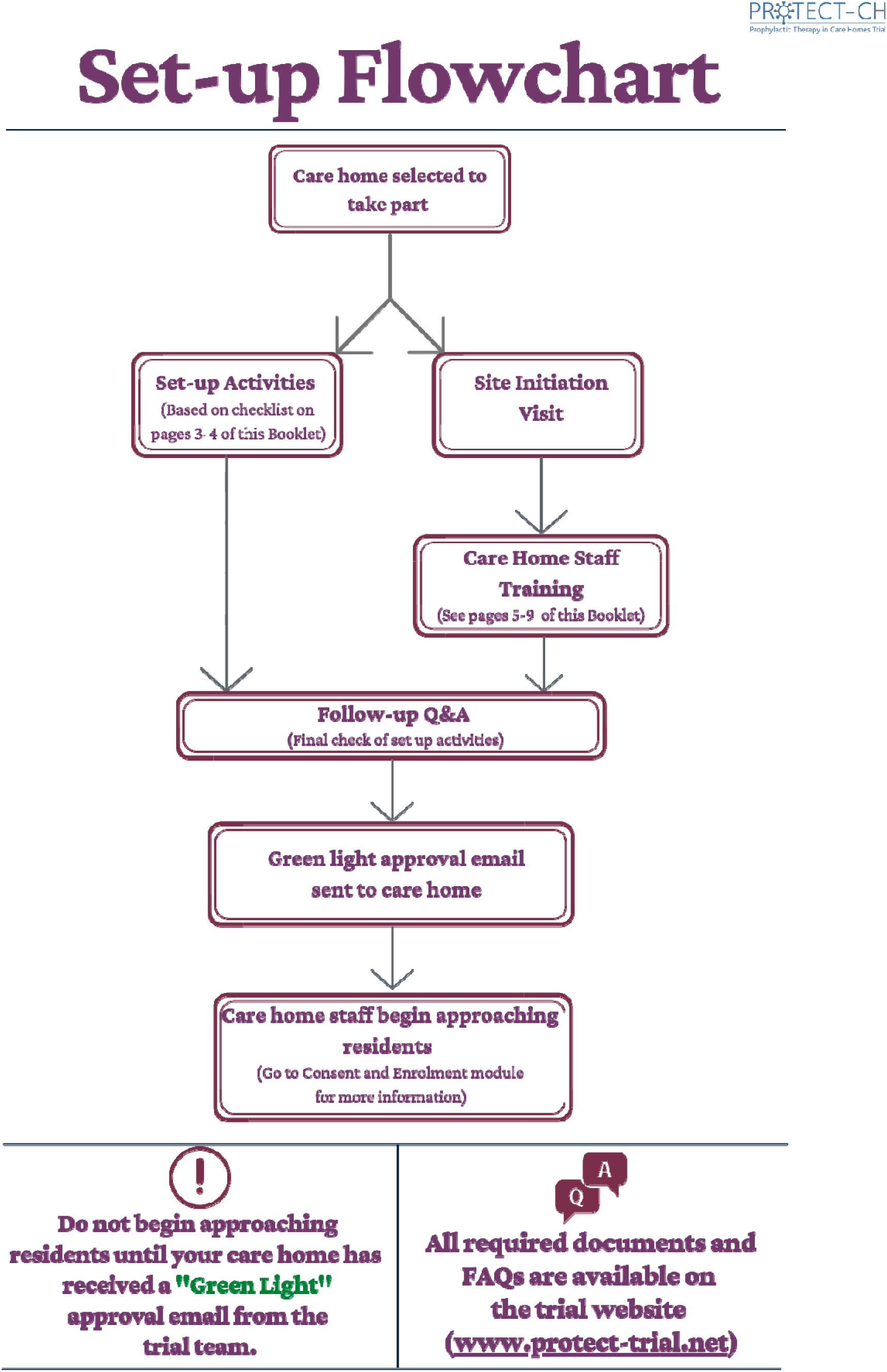
Flowchart for setting-up a care home.

**Figure 9.**
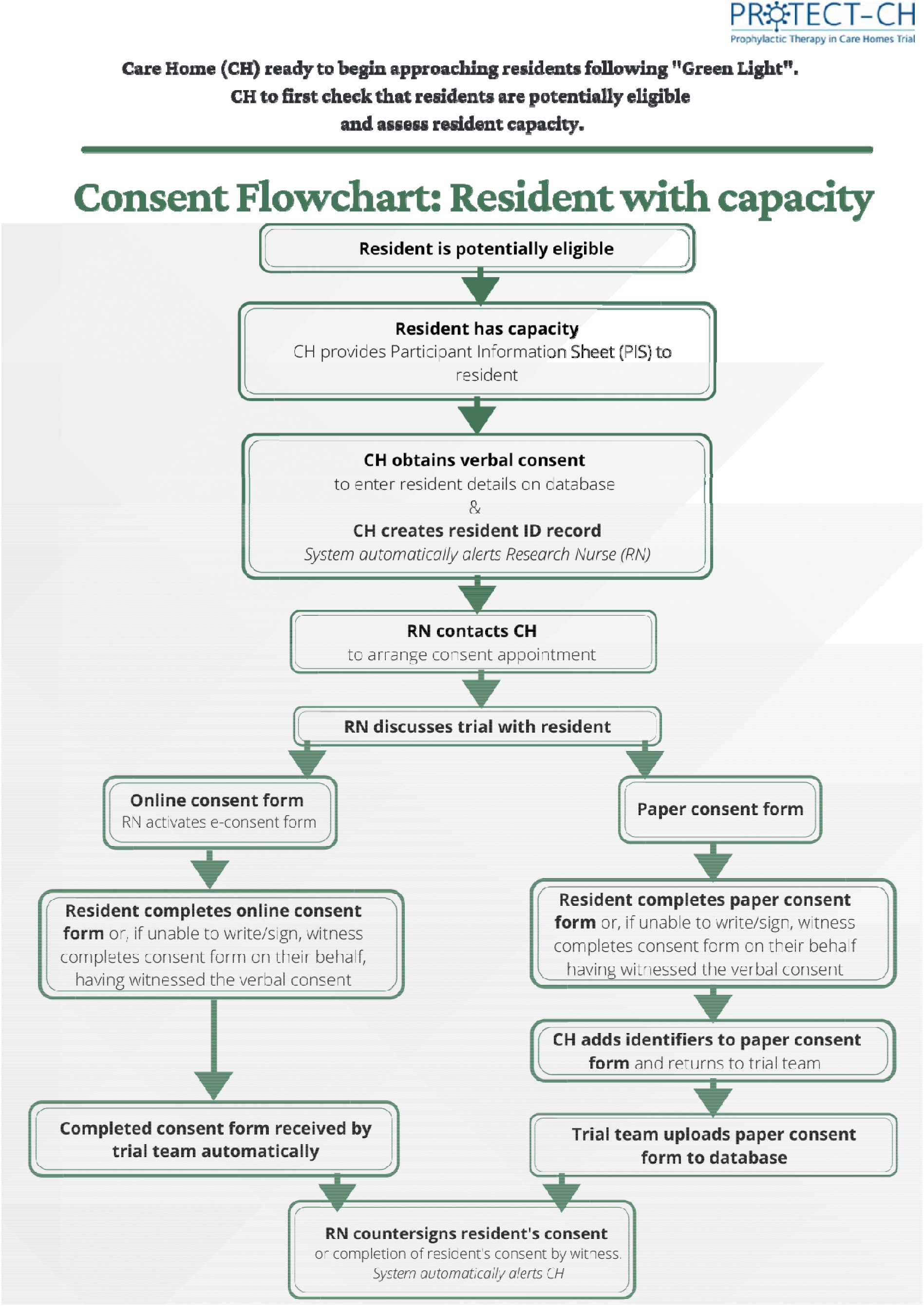
Flowchart: Consent - Residents with capacity.

**Figure 10.**
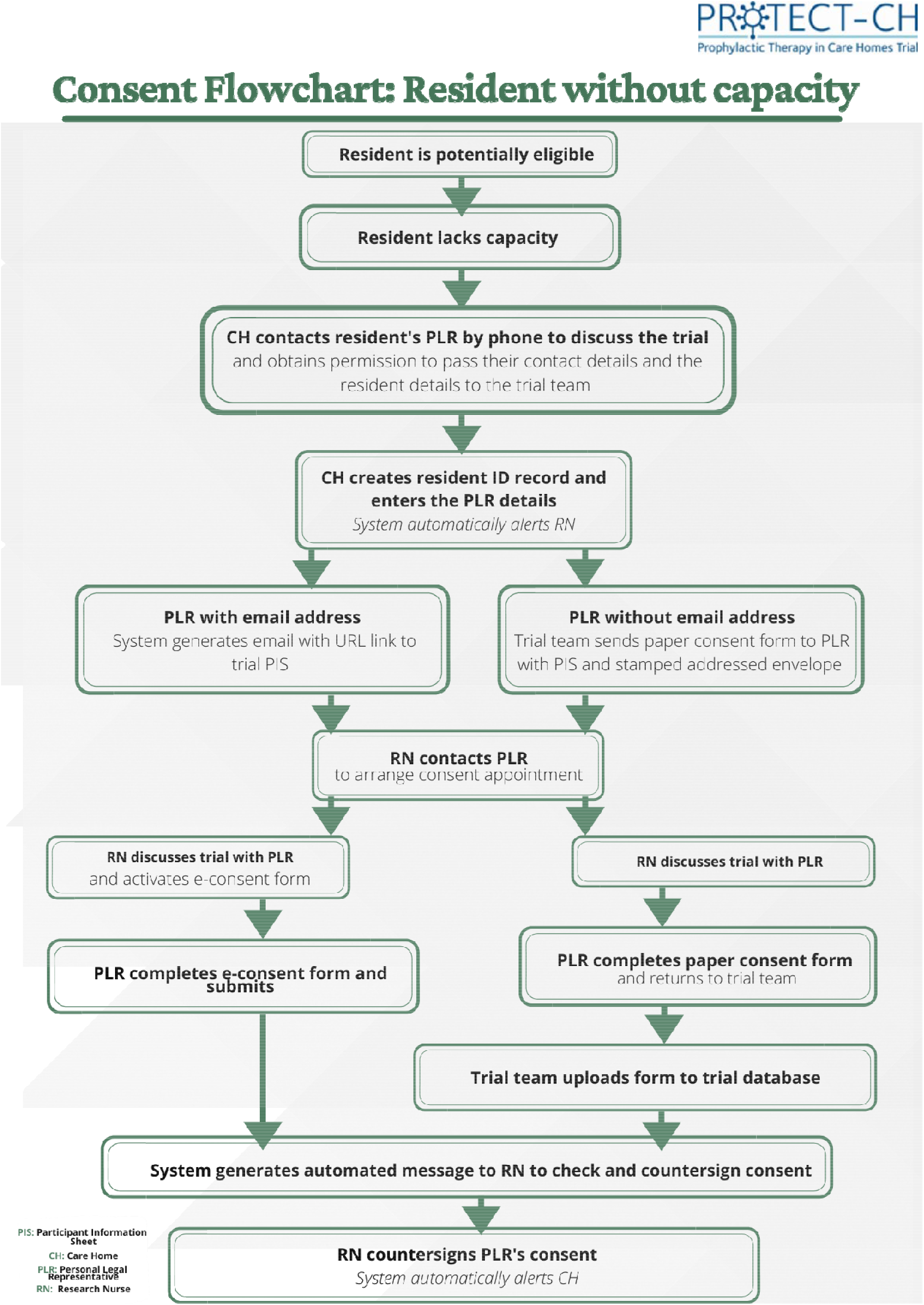
Consent flowchart: Residents without capacity.

#### Care home recruitment

Care homes that expressed interest in participating in the trial/platform were to have been assessed for suitability as follows (Figure 7):

1. Confirmation that CH fulfils eligibility based on inclusion/exclusion criteria.
2. Confirmation from the relevant comprehensive local research network (CLRN) that they had research nurse capacity.
3. Confirmation that GPs associated with the CH would be willing to support their patient’s/resident’s participation.
4. Confirmation that the CH was willing to contract with the NCTU coordinating centre.
5. Training of care home staff.

Challenges related to identifying care homes and general practitioners are discussed in chapter 3.

#### Resident eligibility and consent

Care home staff were to provide a superficial triage of residents for their outline eligibility for joining the trial. Following identification of eligible residents, research nurses led the consent process:

1. Assessment of capacity using a version of the three question approach if uncertain as to whether the resident had capacity.^63, 64^
2. Residents with capacity: consent led by the research nurse (RN, Figure 7).
3. Residents lacking capacity: the research nurse obtains proxy consent from the personal legal representative (PLR, Figure 8).

Consent could be performed either face to face or virtually with residents or PLR.

#### Main eligibility check

Following consent/proxy consent, a full eligibility check would be performed by the GP:

1. Coordinating centre notify GP that resident has consented/assent has been completed.
2. GP completes online eligibility check on REDCAP.
3. Resident is included.
4. GP (or delegate) uploads Summary Care Record to REDCAP.

#### Care home outbreak with positive SARS-CoV-2 test

Following recruitment and consent, residents were to wait for their care home to have a positive SARS-CoV-2 test in either a member of staff or resident based on either a lateral flow or a polymerase chain reaction test (Figures 11, 12, 13, 14), before being randomised into the trial.

**Figure 11.**
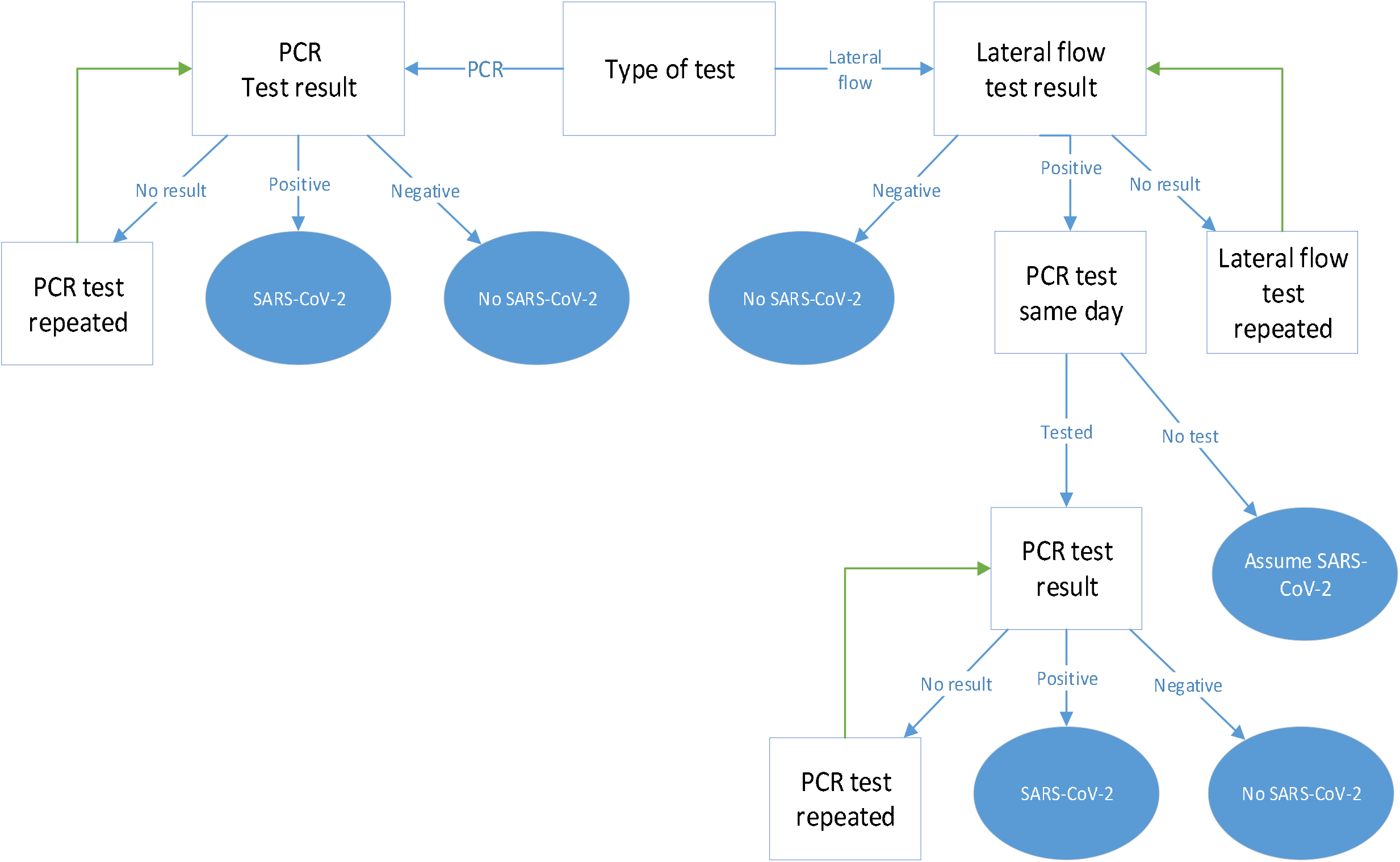
SARS-CoV-2 infection scenarios after a test in PROTECT-CH.

**Figure 12.**
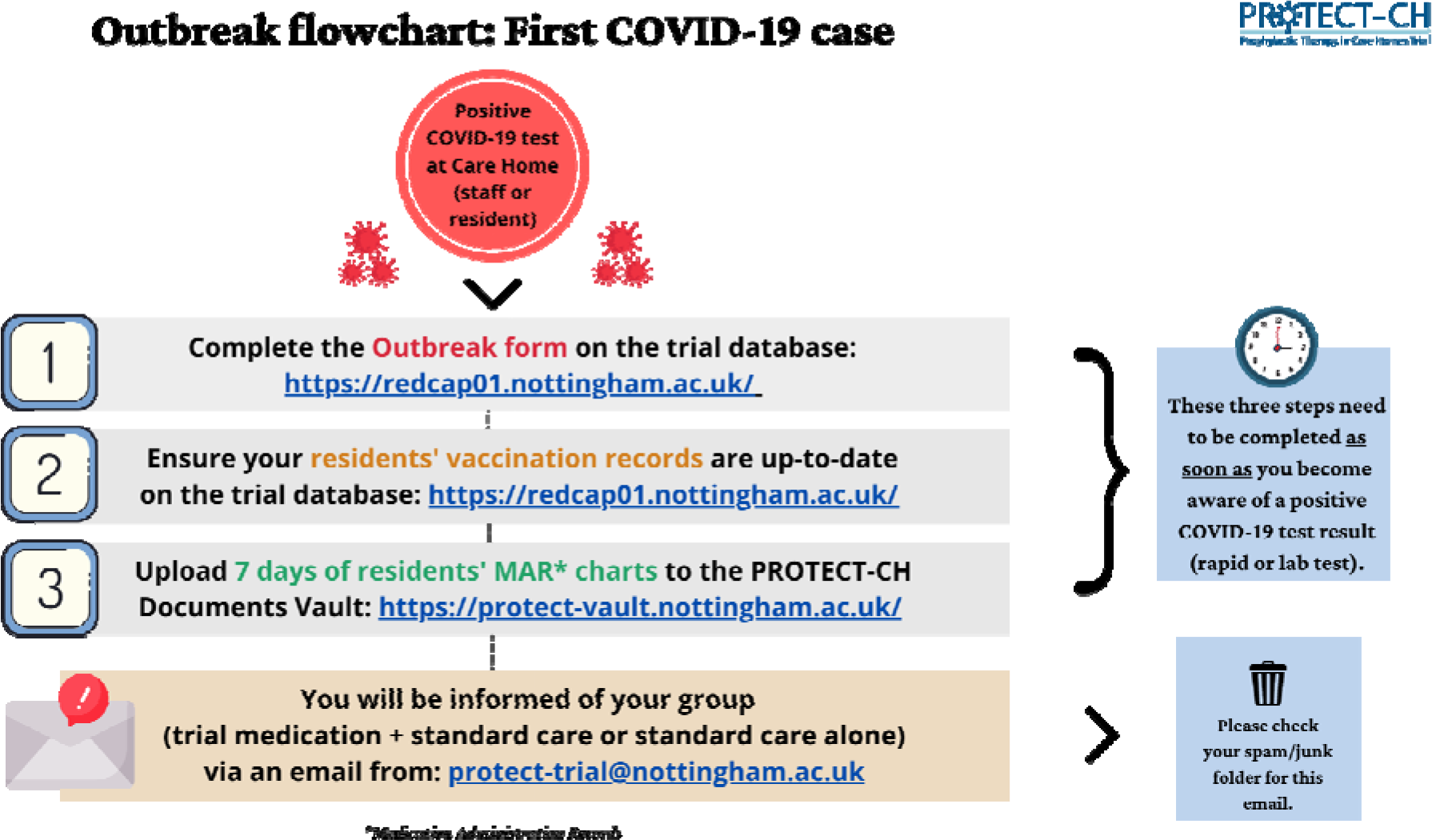
Flowchart: Outbreak - first COVID-19 case.

**Figure 13.**
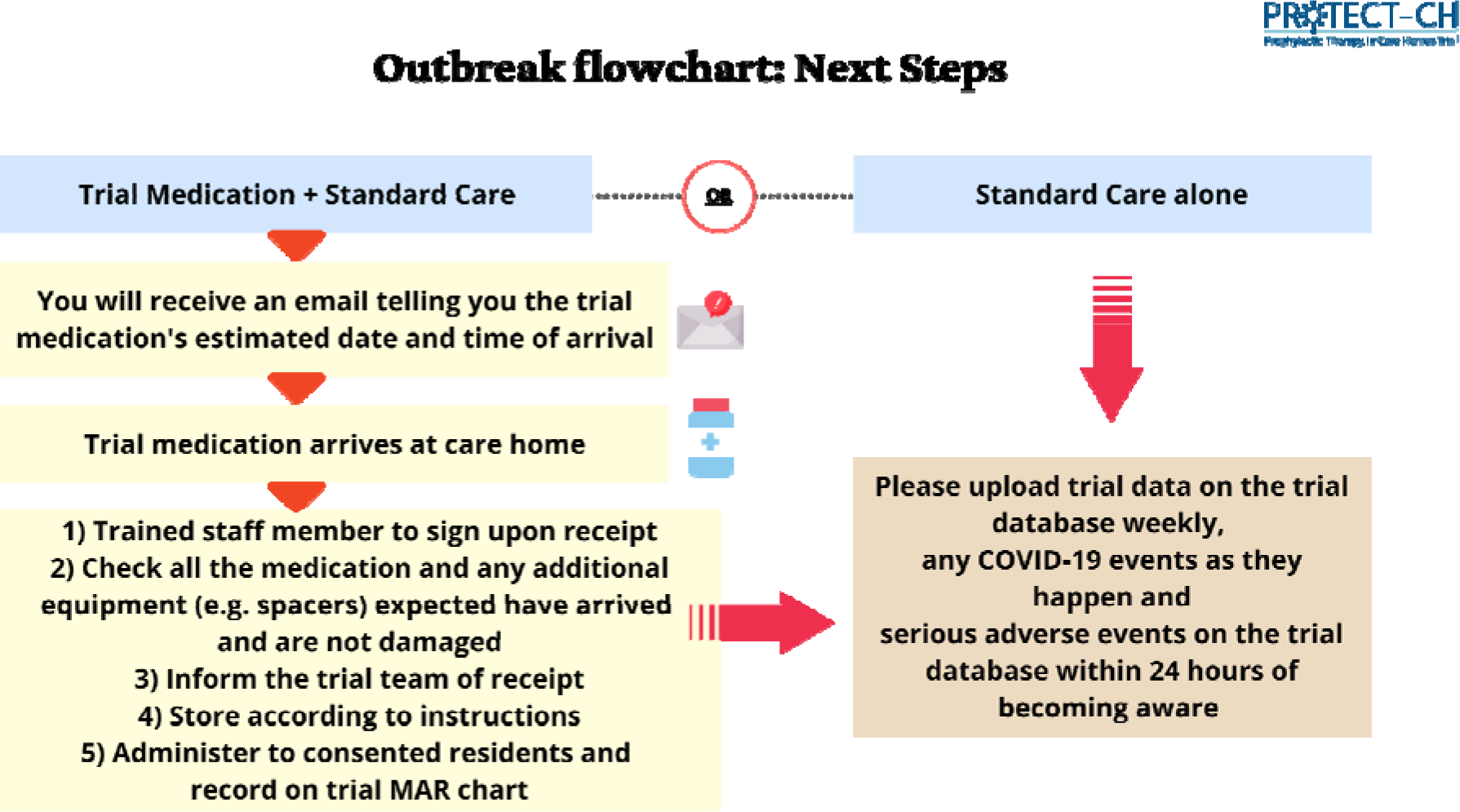
Flowchart: Outbreak - next steps.

**Figure 14.**
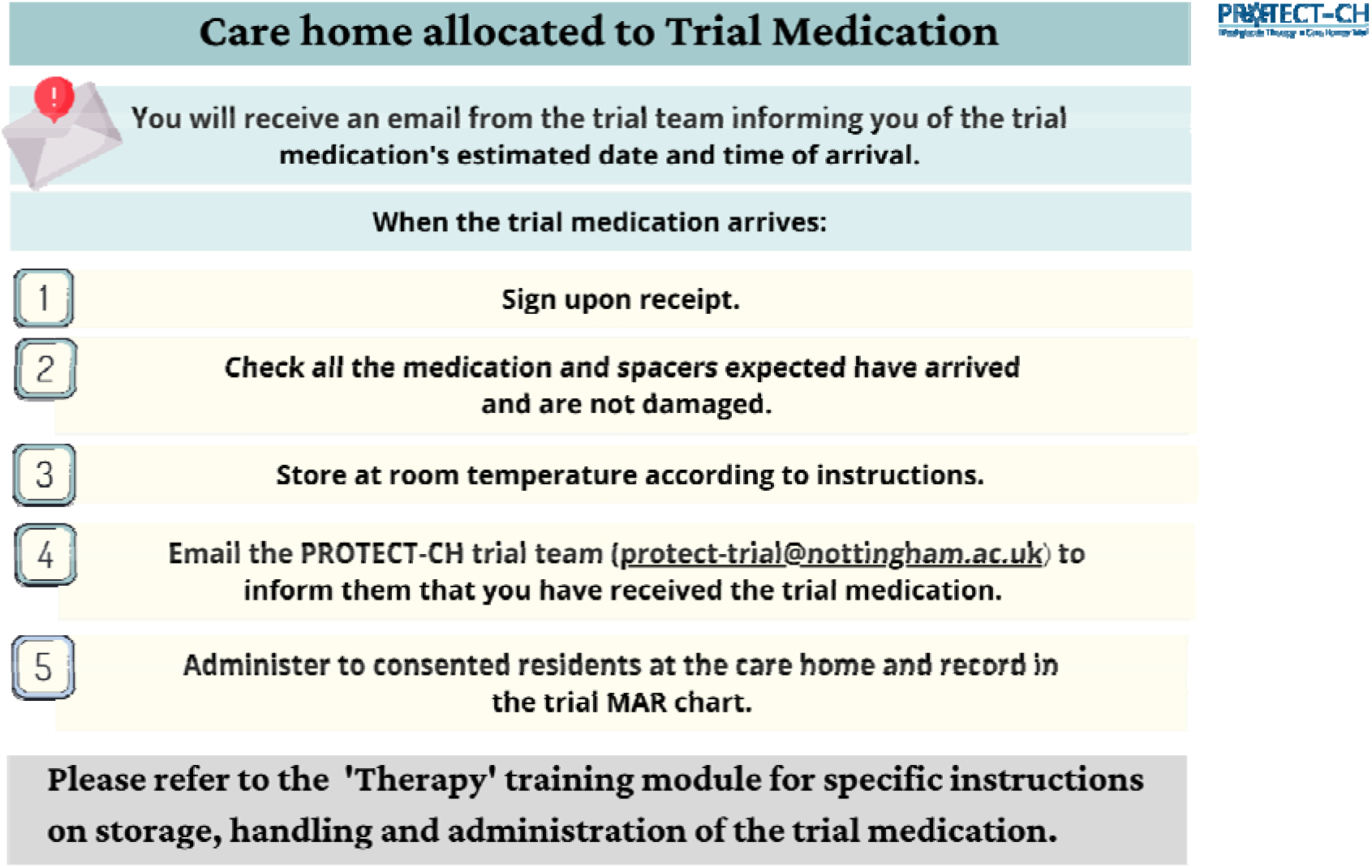
Flowchart: Care home allocated to trial medication.

Once a care home went positive, they were to inform the coordinating centre of which residents were still eligible to take medication (e.g. still alive and not in hospital); care homes would immediately forward electronically a medicine administration record (MAR) chart to the coordinating centre via the Vault database. The duty PI, available by rota 7 days a week from 8am to 8pm, with support from other PIs, were to confirm whether each resident was still eligible based on their Summary Care Record held in the secure data repository and Medication Administration Record, uploaded that day by the care home. Once complete, the PI would randomise the care home to ciclesonide vs niclosamide vs control and submit a signed prescription for active randomisations to the central pharmacy who would then dispense and distribute the relevant medication.

If the initiating SARS-CoV-2 tests was based on an LFT, this would be checked (as is routine) with a PCR test; if this was negative medications would not be dispensed (if possible) or stopped (if administration had already started) and returned to the central pharmacy for destruction (Figure 15).

**Figure 15.**
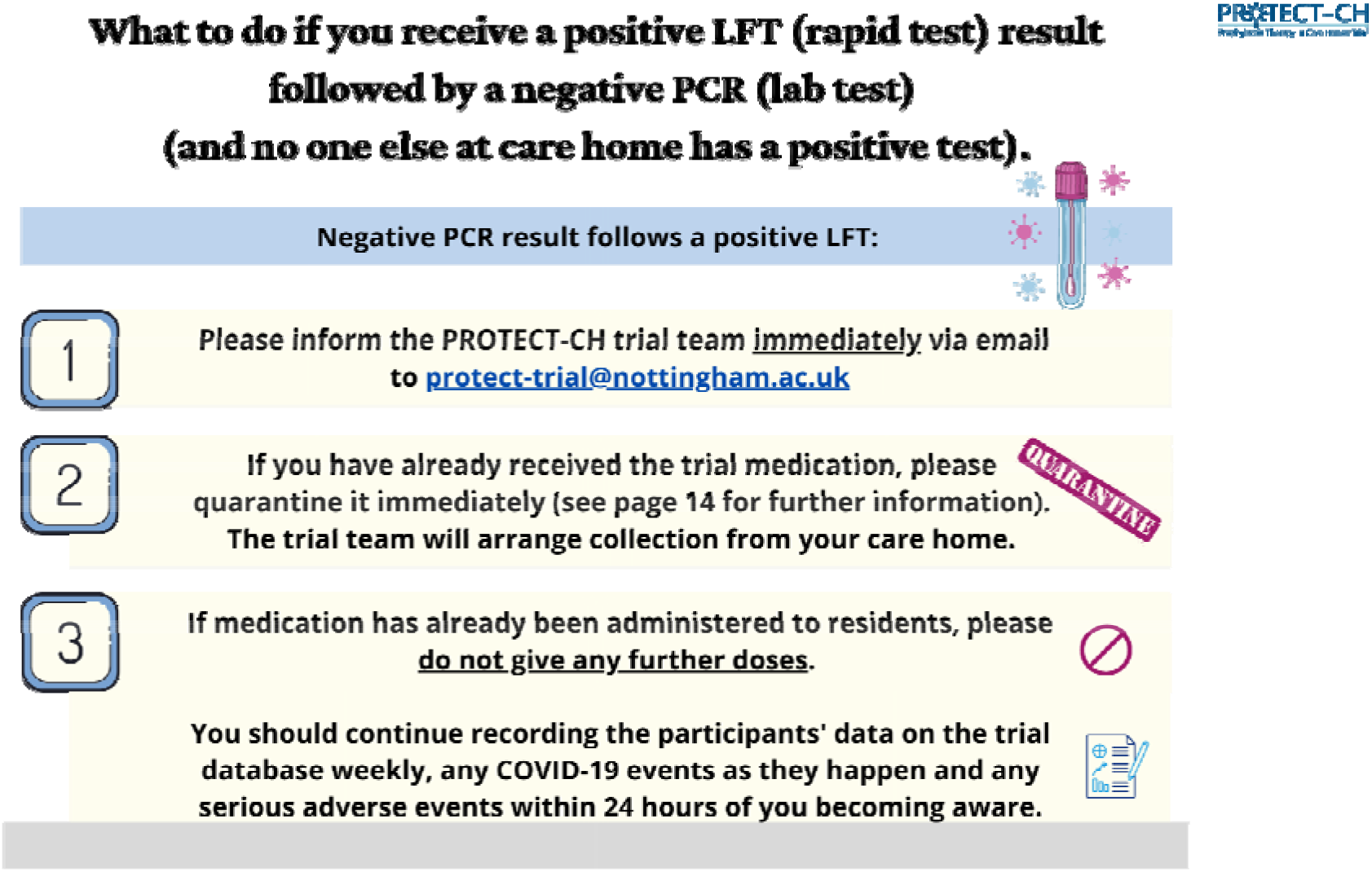
Plan for a positive lateral flow test (LFT) followed by a negative polymerase chain reaction (PCR) test.

#### Training

A suite of training materials were developed targeted at care home staff, PIs, RNs and GPs. Some training was considered mandatory, e.g. related to trial specific good clinical practice and background to the trial (Figure 16). Training materials comprised slide sets, videos, booklets, trial manual, pocket cards and/or posters developed using Microsoft Word and PowerPoint and Canvaa.

**Figure 16.**
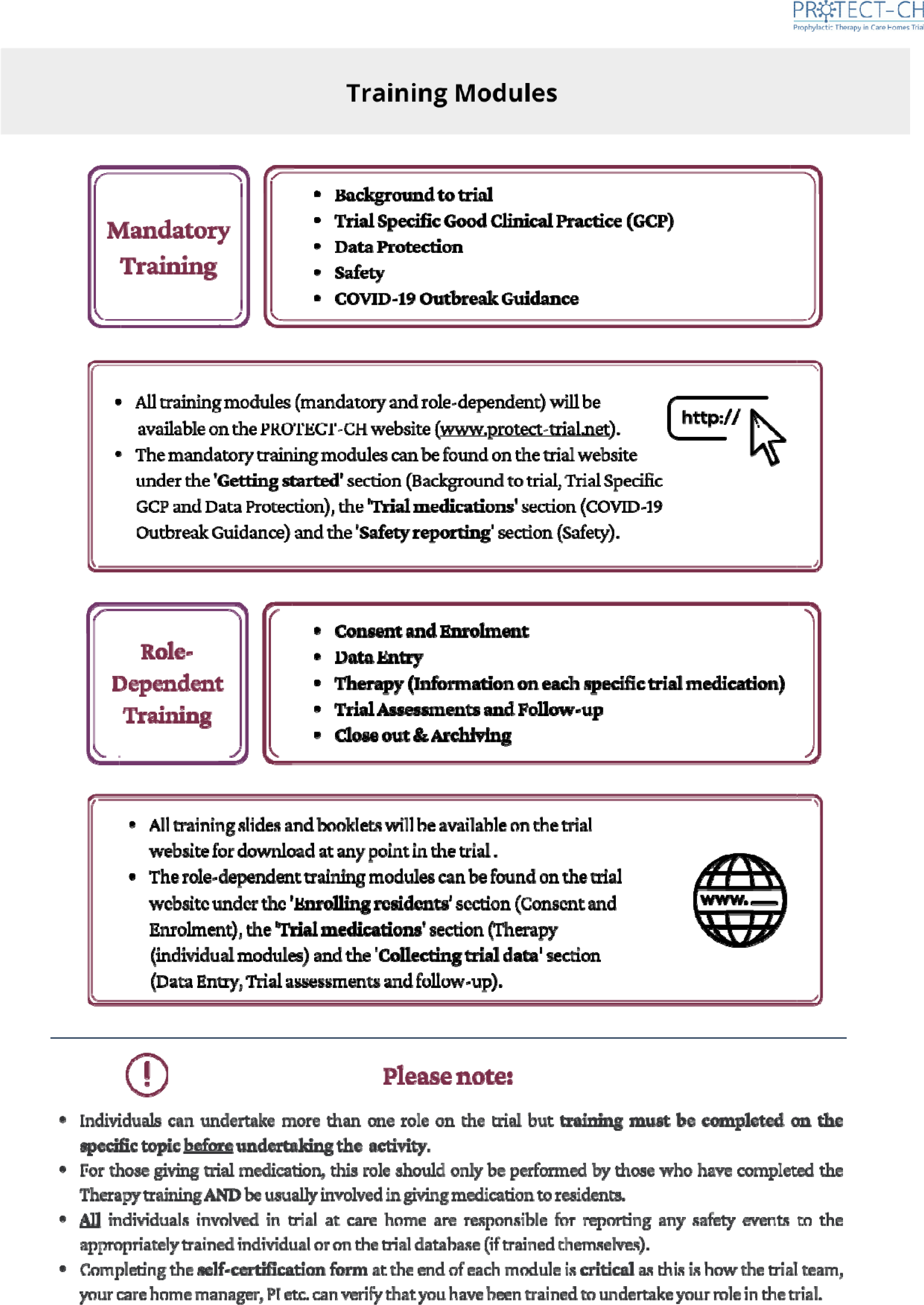
Training summary.

The following modules were developed:

1. Site initiation visit - slide deck and animation video
2. Introduction - video
3. Background to trial platform - slide deck and animation video
4. Trial specific good clinical practice (GCP) - slide deck, video with voiceover
5. Data protection - slide deck, animation video
6. Set-up - slide deck with voiceover, guidance booklet with checklist
7. Consent - slide deck, poster, guidance booklet
8. Therapy, ciclesonide - slide deck
9. Therapy, niclosamide - slide deck
10. COVID-19 outbreak - guidance booklet and checklist
11. Safety - slide deck and pocket card
12. Assessments and follow-up - slide deck and animation video
13. Data entry - user guide, animation video
14. Principal investigator training - slide deck
15. Research nurse training - slide deck, user guide
16. General practitioner training - slide deck
17. Close out and archiving - slide deck

Each product was developed by a trial manager and then quality assessed by a senior trial manager, members of the executive committee and then signed off by the CI. All modules are now present on the platform website. Several of the figures shown here come from these training modules.

### 2.17. Patient and public involvement

The trial was designed and delivered with active support from two PPI members (Maureen Godfrey, Val Leyland) working as part of a wider PPI group, including designing, developing and writing patient and relative information sheets and care home staff training materials., They were members of the Platform Steering Committee. In particular they have shaped the information about the trial, so it is accessible and understandable to care home staff, residents and their families and aimed to view the trial from their perspective at all times to make it as grounded as possible.

Attendance at virtual Trial meetings was initially challenging for PPI members (e.g. use of MS Teams and Zoom) but they rapidly adapted and enjoyed the experience.

The PPI members won the University of Nottingham Community Volunteer of the Year (Group) award in June 2021 (https://www.nottingham.ac.uk/news/local-volunteers-recognised-for-dedication-to-covid-19-care-home-trial, accessed 3 October 2021)

A detailed report on PPI activities, successes and challenges is in preparation.

### 2.18. Contracting

The platform was expected to be very heavy on contracting and Table 4 identifies contracting areas and their status at close down.

**Table 4.** Contracting status.

| Contract | Contractors | No. of contracts |
| --- | --- | --- |
| ***Signed*** |  |  |
| Main programme and funding contract | NIHR-University of Nottingham | 1 |
|  | UoN - Applicants' Universities | ~15 |
| Host website | FRY-IT | 1 |
| ***Not signed*** |  |  |
| Care homes | Care home/chains | ~750 |
| General practitioners | General practices | ~3000 |
| Principal Investigators | Universities/hospitals | ~30 |
| Research Nurses | Hospitals/CRN | ~30 |
| Service delivery agreements | Pharmaceutical companies: Ayrtons, Union | 2 |
|  | Therapeutics |  |
|  | Central pharmacy | 1 |
|  | Site management system (University of Aberdeen) | 1 |
|  | NHS Digital | 1 |
| Trusted Research Environment | University of Dundee | 1 |

### 2.19. Trial management

Multiple systems were designed and developed to support day-to-day trial management (Table 5). Systems used REDCap websites and Adobe Sign to support these activities.

**Table 5.** Trial management processes and systems.

| System | Facility |
| --- | --- |
| Delegation logs | PIs to authorise other PIs to deputise for them, i.e. all PIs can supervise other CHs as necessary |
|  | PIs to authorise CH managers to act as CH lead |
|  | CH managers/CH lead to add/remove staff |
|  | PIs authorise RNs |
| Consent | Support for both eConsent (using REDCap) and paper-based consent for both residents with capacity and personal legal representatives (PLR, if resident lacks capacity) |
| Eligibility assessment | GPs assess outline eligibility of interested residents and upload summary care record (SCR) to secure Vault |
|  | CH staff upload medicine administration record (MAR) to secure Vault once CH has a positive test |
|  | PIs assess final eligibility based on SCR and MAR chart prior to prescribing randomised intervention |
| Randomisation | Configurable stratification/minimisation module to support addition of new drugs to platform and /or removal of ineffective drugs from platform |
|  | Configurable random factor to prevent prediction during stratification/minimisation |
| Trial Master File | Document management system using SharePoint: configurable for read/edit/approval |
|  | permissions by multiple staff, check-out/in control, all actions logged, automatic version control, meta-tagging to facilitate searches |
| Data export | From REDCap to trusted research environment (TRE) |
| Data communication | Bespoke REDCap module for interfacing with external database application programming interface (API), e.g. to Aberdeen Centre for Healthcare Randomised Trials CHaRT site management system |

## Chapter 3. Progress of platform

### 3.1. Progress

#### Timelines

As a COVID-19 pandemic platform, the timelines from NIHR call to project start were extraordinarily short with platform call in late October 2020 and start of grant at the beginning of January 2021 (Table 6). The grant started on 1 January 2021 with the start-up phase lasting to end April 2021; with two drugs to test, we needed one year to test it, as illustrated in the Gantt chart (Figure 17).

**Figure 17.**
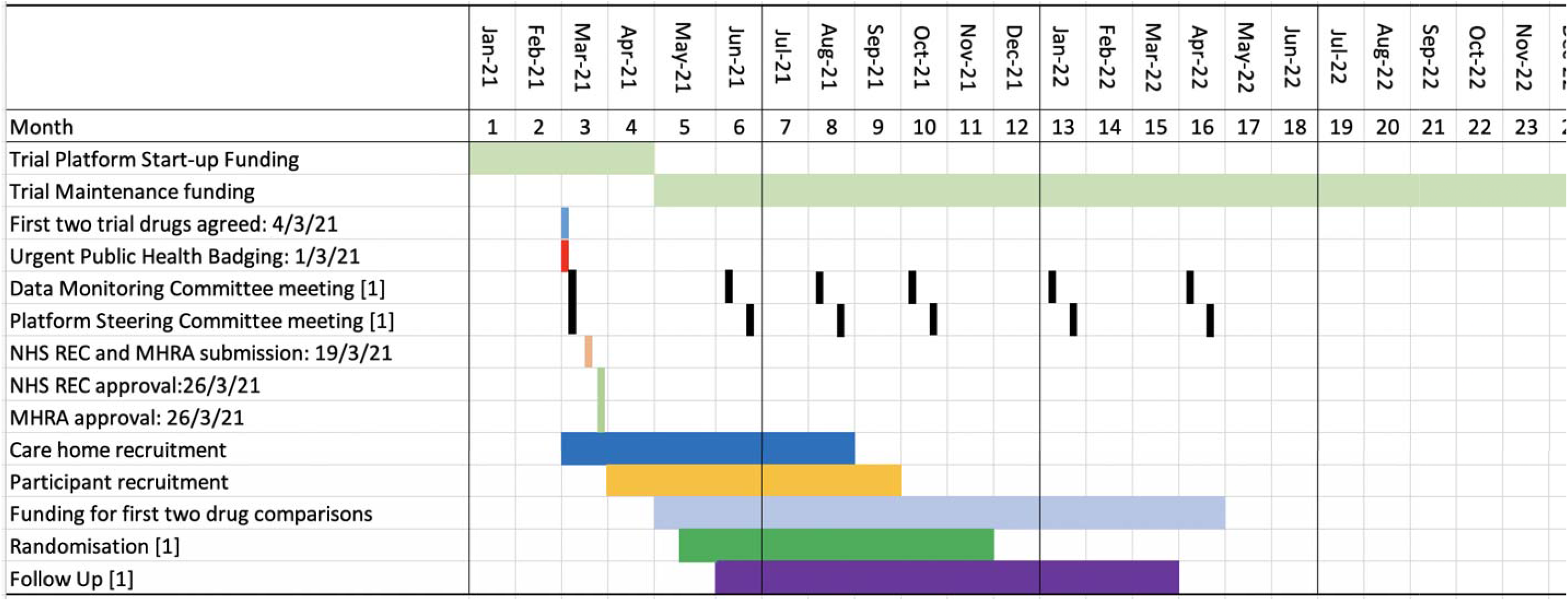
Gannt chart of trial platform flow for testing first two drugs and through to end of two-year contract.

**Table 6.**
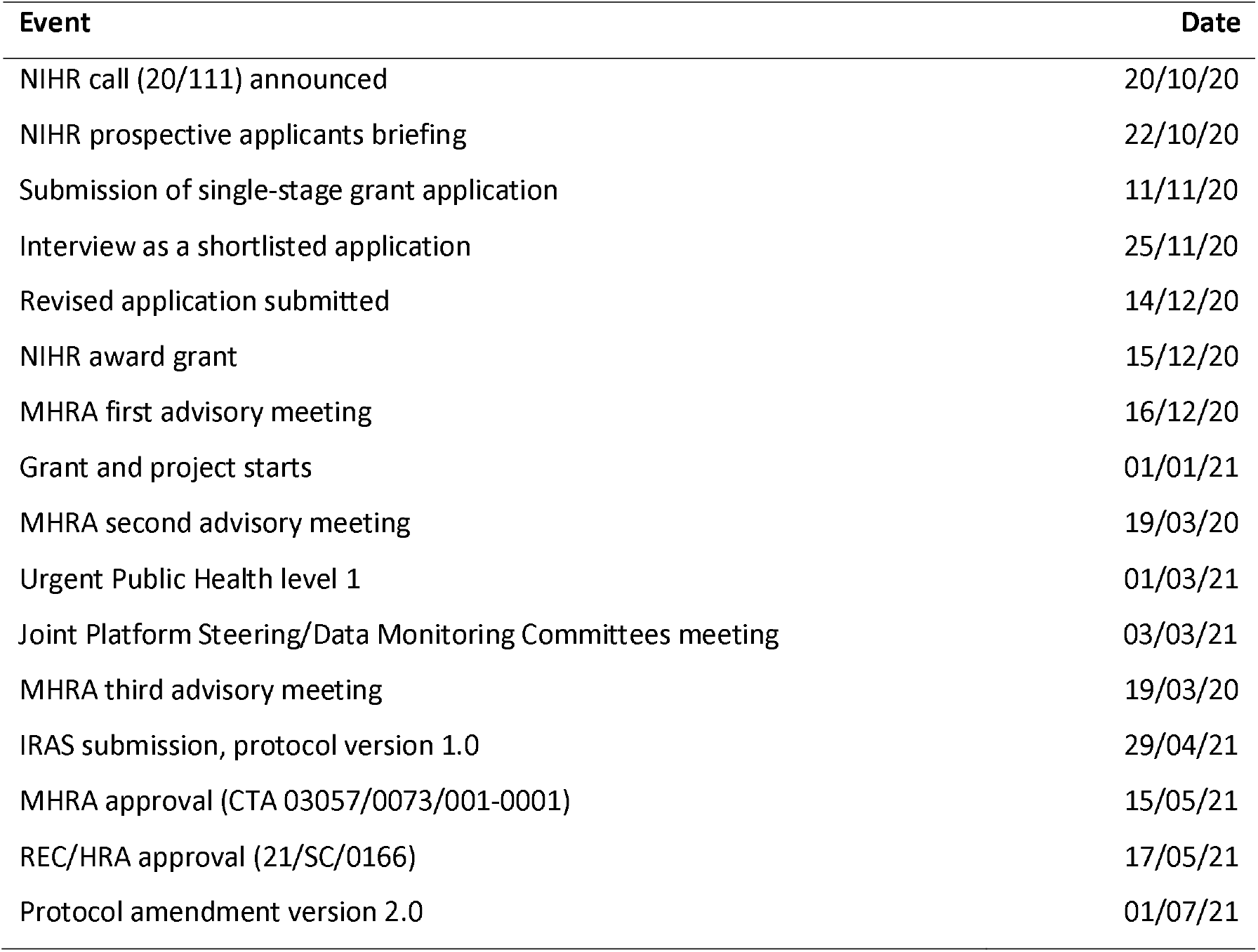
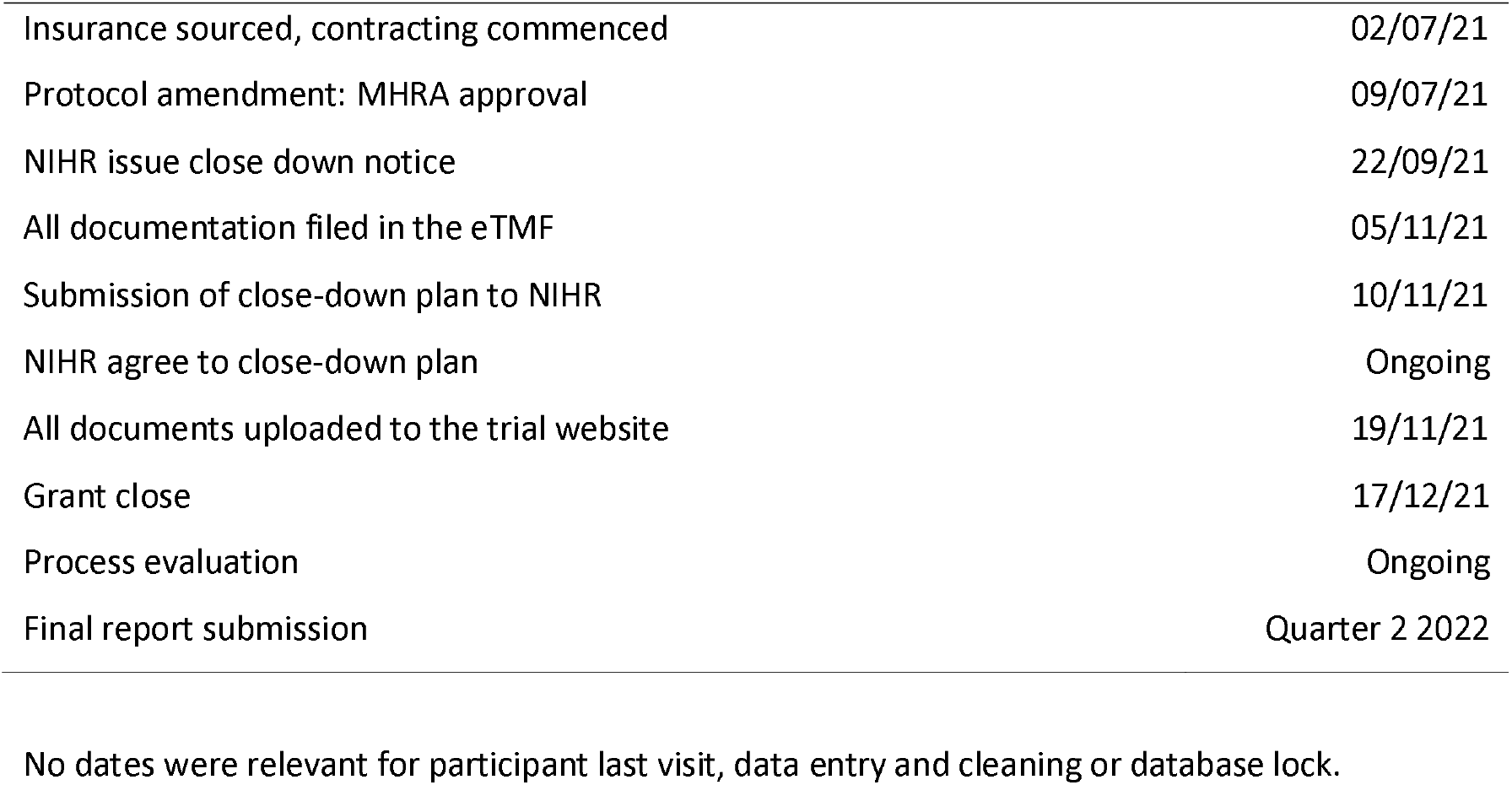
Record of key dates.

| Event | Date |
| --- | --- |
| NIHR call (20/111) announced | 20/10/20 |
| NIHR prospective applicants briefing | 22/10/20 |
| Submission of single-stage grant application | 11/11/20 |
| Interview as a shortlisted application | 25/11/20 |
| Revised application submitted | 14/12/20 |
| NIHR award grant | 15/12/20 |
| MHRA first advisory meeting | 16/12/20 |
| Grant and project starts | 01/01/21 |
| MHRA second advisory meeting | 19/03/20 |
| Urgent Public Health level 1 | 01/03/21 |
| Joint Platform Steering/Data Monitoring Committees meeting | 03/03/21 |
| MHRA third advisory meeting | 19/03/20 |
| IRAS submission, protocol version 1.0 | 29/04/21 |
| MHRA approval (CTA 03057/0073/001-0001) | 15/05/21 |
| REC/HRA approval (21/SC/0166) | 17/05/21 |
| Protocol amendment version 2.0 | 01/07/21 |
| Insurance sourced, contracting commenced | 02/07/21 |
| Protocol amendment: MHRA approval | 09/07/21 |
| NIHR issue close down notice | 22/09/21 |
| All documentation filed in the eTMF | 05/11/21 |
| Submission of close-down plan to NIHR | 10/11/21 |
| NIHR agree to close-down plan | Ongoing |
| All documents uploaded to the trial website | 19/11/21 |
| Grant close | 17/12/21 |
| Process evaluation | Ongoing |
| Final report submission | Quarter 2 2022 |

No dates were relevant for participant last visit, data entry and cleaning or database lock.

However, it became clear in August 2021 that delays in drug contracting and the profound efficacy of vaccines and so reduced infections in care homes meant that the platform could no longer meaningfully test the scientific question. Following discussions between the applicants, University of Nottingham, NIHR and DHSC, NIHR issued a close down notice on 22 September 2021 and the grant ceased on 17 December 2021 (Table 6).

This chapter explores these events, issues and associated learnings. First we discuss what was delivered and then the impediments that led to the project closing early.

#### Protocol

The trial was conceived and designed by the grant applicants who wrote the protocol. Protocol version 1 was fully approved:

https://www.protect-trial.net/files/documents/protect-ch-protocol-final-v1-0-13-may-2021-1.pdf

One protocol amendment (version 1 to 2) was made during the trial; this was submitted on 1 July 2021 and approved by MHRA and HRA on 9 July 2021:

https://www.protect-trial.net/files/resources/protocol-final-v2-0-01-jul-2021_for-website-corrected-1.pdf

#### Urgent Public Health badging

Following confirmation of the two potential drugs to test, the study was given NIHR Urgent Public Health level 1 badging (https://www.nihr.ac.uk/covid-studies/, downloaded 30 December 2021) on 1 March 2021.

#### Ethics approval

The IRAS submission was made on 29 April 2021 with the CI and Co-CI attending the South Central – Oxford A Research Ethics Committee on 7 May 2021. Following minor changes to the PIS and ICF, UK Research Ethics Committee and Health Research Authority gave approval (21/SC/0166) on 17 May 2021.

#### MHRA approval

Prior to the IRAS submission we sought advice from Medicines and Healthcare products Regulatory Agency on two occasions (16 December 2020 and 19 March 2021). Guidance from these meetings were that a trial testing unlicensed drugs in a vulnerable population would require active involvement of each participating resident’s general practitioner, which we had not planned or costed for. Following IRAS submission, we received MHRA approval (CTA 03057/0073/001-0001) on 15 May 2021.

#### Registrations

The trial/platform was registered with the European Union Drug Regulating Authorities Clinical Trials Database (EudraCT 2021-000185-15). Initially we planned not to register with other databases but with BREXIT, it made sense to also submit to ISRCTN and this was in progress when the decision to close the programme was made.

### 3.2. Media and publications

#### Media

Written, radio and television media and press are presented in Appendix 5.

#### Publications

***To date***

None

***Planned***

- This report
- Process evaluation
- PPI experience
- Plain English summaries

### 3.3. Delays

Having started the grant on 1 January 2021, our original intention was to design and develop the trial for four months and then start contracting care homes in early May 2021 and recruiting residents in late May 2021. Two logistical challenges (Table 4), significantly delayed the start:

1. Protracted negotiations between NIHR and the donor pharmaceutical companies. Key differences between NIHR and the companies were:

a. NIHR access to background intellectual property;
b. Company access to anonymised individual patient data after the trial completed and use of this for licensing and marketing purposes.
2. Identifying a source of insurance for care homes since their own insurance did not cover research. At one stage, government might have had to cover the trial under the Coronavirus Act 2020. Eventually a company was identified who was willing to insure care homes and their staff as well as principal investigators and research nurses.

While the above issues were ongoing it became apparent that the phenomenal success of vaccination meant that transmission and illness rates were much lower than when the trial was planned and so rather than needing 750 care homes and 24,000 residents, the trial would need to have recruited many thousands of care homes and more than 100,000 residents. These factors were compounded by multiple other delaying issues as summarised in Table 7.

**Table 7.** Summary of main challenges and issues that complicated and delayed the trial and learning points.

| **Challenge** | **Summary of main issues** | **Learning points** |
| --- | --- | --- |
| IMP identification | There was lack of clarity as to which committee was to identify the drugs to choose, i.e. UK COVID-19 Therapeutics Advisory Panel (UK-CTAP) or NIHR Prophylaxis Oversight Group (POG); recommended drugs differed between these two committees. | A single committee should identify the preferred drugs. This would accelerate trial start-up and ensure that DHSC and NIHR were fully behind the trial. |
| IMP identification | NIHR instructed us not to publicise what IMP were to be tested until contracts with the manufacturers had been signed.<br>This delayed recruitment and contracting with care homes and GPs.<br>This reduced our ability to advertise the trial to media and their interest in the trial. | Whether announcement of IMP should be made without contracts is a moot point. |
| IMP contracting | Delay in obtaining drug due to a lack of agreement between NIHR and the companies over contracts.<br>If this delay had been resolved it is probable, all other things being equal, that the platform trial could have started contracting in May 2021. Once started, the trial would likely have continued to recruit, treat and follow-up residents and run through the winter of 2021. | The key driver of patient benefit must underline all processes.<br>NIHR should consider whether companies must agree to handing over background IP.<br>NIHR should agree on use of trial data for licensing and marketing purposes before the trial starts rather after the results are available. This would reassure companies that they would have access to data and would accelerate licensing, marketing and patient benefit in the event of a positive result. |
| Research nurses | Research nurses are essential in a trial testing IMP in care homes. | Research delivery organisations such as CRN need to identify |
|  | There may be insufficient numbers of RNs to support a large trial-platform testing IMP, especially in a PEP design where several RNs may be needed at very short notice when a CH needs to start immediate treatment.<br>How to contract with hospital-based RNs was unclear, as acute research trusts often did not have governance arrangements in place to allow their nurses to visit care homes. | how they can support care home based trials in respect of numbers of RNs needed, their skill set for recruiting, consenting and treating in a CH population, and how to make them available at very short notice in a PEP design. |
| General practitioners | MHRA requirement that all residents must have their GP participate in the trial through providing information on medical history and potential reasons for exclusion.<br>Care homes may have between one and ten GPs associated with them resulting in a very large number to contract with and train. GPs are often, in addition, responsible for more than one care home.<br>Without knowing which residents were going to sign up to the trial platform or being able to discuss which drugs were to be tested, it was not possible to formally start contracting with GPs. | GP contracting and training will be a massive load on any large trial in care homes if unlicensed drugs are being tested. Testing a repurposed licensed drug (rather than an unlicensed drug) would reduce the need for detailed safety reporting and perhaps involvement of GPs. Testing of a non-IMP<sup>58</sup> would mean MHRA and GPs would not need to be involved and would simplify safety recording and reporting. |
| CH identification | Although all care homes are listed on CH registers (Table 8) across the UK, it is challenging to work out from this who the primary contact for each care home should be. The registered manager may not have “sign off” responsibility for involvement in research, particularly in chain providers.<br>Changes in personnel and their contact details (e.g. emails) change frequently so that registers were frequently out of date. All potentially interested care homes and chains were unwilling to progress contracting without knowledge about the interventions. | A pan-UK register should host an accessible database of care homes and contact details to facilitate research. |
| CH ownership | Unlike UK NHS hospitals, CHs are variously owned by commercial chains and charities with a few being council or NHS. Hence, NHS research and governance processes do not translate automatically to social care |  |
| CH staffing | Often insufficient staff numbers to support research <sup>58</sup> due to enhanced care during a pandemic (e.g. due to increased infection control), illness, Brexit and, most recently, mandatory vaccination that has resulted in a significant minority of staff moving out of the sector. | Specific resources for supporting CHs should be costed into future platforms; relying on support from CRN/EnRICH and Service Support Costs is insufficient. |
| CH communication | Challenge to find suitable times to discuss trial with coordinating centre. | Regular forums addressed some of this difficulty but finding a time convenient for all managers was difficult.<br>Specific resources for running CH forums should be costed into future platforms. Social media solutions, such as a WhatsApp group, should be used to support such forums and be staffed by research teams to enable CHs to interact regularly, in a responsive way, with researchers. |
| CH research experience | Challenges of introducing a trial to a research-naïve group of staff who may not know about good clinical practice, consent, IMP administration and management, and data integrity and management. CH funding and care delivery models also impeded their sign-up. | Detailed and repeated training addresses these problems.<br>Involvement of research nurses to assist with drug administration and data collection. |
| CH as an institution versus being a resident's home | Governance and insurance were complicated by interpretation of the role of a CH in a resident's life. For example, GPs may enrol people living at home into a trial but not in an institution. And yet, many CH resident would argue that the CH is their home. | - |
| Insurance | Identification of insurance to cover care homes delayed the trial by several | Companies insuring care homes should consider including |
|  | months. | research in their policies. Clinical trials have a very low risk of legal action and incurring costs for insurance companies whether drugs are licensed or not. NIHR should prioritise solving the governance issues surrounding CH research, including insurance, a strategic priority. |
| Contracting | Contracting would involve many hundreds of care homes, thousands of general practitioners and tens of PIs and RNs. | Standardised template contracts, drafted with and reviewed by care homes umbrella organisations, should be established.<br>Pilot initiatives, such as the EnRICH Cymru model (https://healthandcareresearchwales.org/enrich-cymru). Accessed 3/1/22), where EnRICH staff scrutinise and “vouch for” research contracts to care homes, should be replicated nationally.<br>Similar initiatives to centralise contracting with GPs, for example at the PCN level, should be considered. |
| Four-nation working | CHs and their chains, structures such as CRN and EnRICH, GP data, data sources (Table 8) and regulations each differ across the four UK nations thereby complicating the design and delivery of a CH trial. | Accepting that devolution makes such differences inevitable, PROTECT-CH identifies some of the solutions that will ease future CH trials working across the UK. |
| SoECAT | The NHS SoECAT model restricted funding that we could offer to care homes so potentially making the trial less attractive to Scottish care homes. | Review SoECAT model for funding of care homes in trials.<br>This should be done in conjunction with care home provider organisations. |
| Research environment | At commencement of the pandemic research was prioritised at all levels, e.g. the RECOVERY trial was able to start within 10 days of conception. | Prioritisation of resources for pandemic research needs to continue for the duration of the pandemic. All research in a |
|  | When PROTECT-CH started in early 2021, many aspects of research were attempting to return to pre-COVID-19 practice and activity, e.g. CRN wanted research nurses to return to supporting non-COVID-19 trials. | pandemic context is time-critical and will be disproportionately affected by research delays by comparison with more routine research. |
| COVID-19 rates | The trial was designed in the midst of the second UK wave when care home resident infection, hospitalisation and death rates were high. The sample size calculation required recruitment of 750 care homes and 24,000 residents. The decision to close was made in late summer when care home resident infection, hospitalisation and death rates were low. Adjustment of the sample size calculation required recruitment of thousands of care homes and ~100,000 residents, which was unrealistic. | N/A |
| Coordinating centre team working | Challenge of remote working by large team at coordinating centre when developing a new trial.<br>Led to protracted discussions. | Establish processes for regular updates, constructive challenge, and resolution of disagreements, including written as well as online/telephone meetings. |

**Table 8.** Organisational challenges in working in all four UK nations. Differing sources of central routine data are shown in Table 2.

| Issue | Challenge | England | Northern Ireland | Scotland | Wales |
| --- | --- | --- | --- | --- | --- |
| CH chains | Most CH medium-large chains not present across all of UK. Most small CH groups are regional. | Barchester<br>Four Seasons<br>HC One | Four Seasons | Barchester<br>Four Seasons<br>HC One | Barchester<br>Four Seasons<br>HC One |
| CH inspectorates and registers | Different organisations in each country so no common list or classification of CHs | Care Quality Commission | Regulation and Quality Improvement Authority | Care Inspectorate Scotland | Care Inspectorate Wales |
| EnRICH | Different organisations in each country | EnRICH (England) | Not applicable | EnRICH Scotland | EnRICH Cymru |

## CHAPTER 4. Learning points and discussion

The phenomenal success of vaccines in reducing transmission and the severity of COVID-19 has resulted in few infections in care home residents since quarter 2 2021. Hence, PROTECT-CH, as designed and funded, became unnecessary.

There were multiple challenges in PROTECT-CH as detailed in Table 7. We will focus here on the key impediments to delivery of the trial.

### 4.1. Contracting with pharma companies

The main logistical reason for a delayed start was the lack of drugs due to a delay in NIHR and the two drug companies agreeing terms. We lost five months over this and could have started recruitment in May 2021 if this had been progressed more efficiently. Particular issues related to:

- Sharing background IP with NIHR in case the company could/would not develop a positive finding, and so waste public money.
- Limiting any agreement such that the company could only access results (especially individual patient data) for the purposes of licensing and marketing once the trial results were known and subject to a commercial agreement.

A key perspective from NIHR is the need to be seen to not be effectively offering state aid through the appearance of giving a company access to data that should be shared more widely. There are two caveats here:

- Both companies were willing to donate the drug, as based on their background IP. For one company, there was no alternative supply of drug. For the other company, the largest supplier was approached but did not respond.
- Both companies were small-to-medium in size. Whilst large companies can negotiate on an “equal footing” with NIHR, we need to consider whether smaller companies can negotiate in an equitable and meaningful way. Of relevance, smaller companies may wish to merge or be taken over by another company and any governance search by a third party would detect a contract that gave the trial drug but gained nothing definite in return.

These positions were complicated since both drugs were unlicensed and so needed more detailed safety reporting. Further a positive trial result would first need a licensing application, this further delaying any benefit to patients. We urge NIHR to take a proportionate decision depending on factors such as company size and licensing status. The alternative is that key scientific questions are never answered, and potential patient benefit is lost.

The issues related to contracting with drug companies is not specific to care home research. Outside of time-pressured pandemic research it is less likely that such protracted negotiations could critically injure a research project in the way they have done here – but such delays replicated across the range of studies co-ordinated by NIHR will, cumulatively, lead to substantial inefficiencies. Reducing the time-delay associated with these processes should be a priority both for future pandemic research and the routine running of the NIHR if efficient use of public resources is to be prioritised.

### 4.2. Insurance

As highlighted earlier, care home insurance does not usually cover research. Our initial searches for insurers failed because many were leaving the care home market in view of perceived risks associated with COVID-19 in care homes. A backup plan was to ask Government to underwrite the trial in care homes through the COVID-19 2019 Act. Eventually, one company was identified who would insure the trial covering care homes, PIs and RNs as well as the University of Nottingham and the members of the oversight committees. The platform contracted with the company and the trial could have started subject to resolution of other issues. This problem delayed start by 2 months.

This is a care home specific issue and is a consequence of the fact that care homes are predominantly private or third sector organisations, not covered by Crown or NHS indemnity. The market for routine insurance in care homes has been harmed substantially by the increased costs borne by insurers during the pandemic. NIHR either need to replicate the insurance arrangements established for PROTECT-CH for all future drug trials in the sector and ensure that these costs are included in the trial overheads, or they need to look at indemnifying the sector against the risks associated with drug trials in a strategic way. This should be done in conjunction with care home providers to ensure that the solutions developed meet their needs and, even more importantly, are recognised and accepted by the sector and the companies that regularly insure them.

### 4.3. Contracting

The sheer volume of potential legal contracting with many hundreds of care homes, several thousand general practitioners and many tens of PIs and RNs challenged the available staff numbers both within NCTU and UoN Research and Innovation. With care homes and general practices being autonomous organisations, it is difficult to see how this workload could be reduced although we note that Hospital Trusts are increasingly moving to a model where decisions made by the lead Trust are then followed by other hospitals without detailed assessment.

Although 299 care homes submitted an expression of interest via the REDCap database page, just 82 confirmed capacity and capability (Figure 7). A common reason for lack of progression was that care homes wanted to know what treatments were going to be administered and we were unable to tell them since we had not contracted with the pharmaceutical companies. As a result, no care homes could be contracted.

Finding a way to contract with such a wide array of organisations in a more feasible way requires careful consideration. It is unlikely that care home organisations will be prepared to negotiate on each other’s behalf given the nature of the sector in the UK as a semi-regulated market. Within this each provider is accountable both to its own shareholders or trustees, and the regulator, for its own governance. Standard template contracts, designed in conjunction with care home representative organisations and scrutinised by lawyers, could potentially streamline these processes. EnRICH Wales have a process whereby administrative staff, known to and trusted by local care homes, can scrutinise research contracts and “vouch” for these. Such arrangements could be replicated elsewhere in the country.

Contracting with GPs raised similar issues and few GPs had agreed to support the trial at the time the decision was made to close it (Figure 6).

Although the opportunities for more standardised contracting approaches are greater because GPs do not operate in a competitive market in the way that care homes do. NIHR should explore opportunities for syndicated contracting with GPs, for example within each UK nation and through the clinical research networks, including primary care network, in order to minimise inefficiencies.

### 4.4. Support for pandemic trials

During the pandemic there has been a steady fall in the success of interventional platform trials; the RECOVERY trial has been most successful followed by REMAP-CAP and then PRINCIPLE. PROTECT-CH was funded much later at the start of 2021 and by May 2021, when we might have started enrolling care home residents, the UK second and third waves of infection ^65^ had declined and healthcare and care home staff who had worked on COVID-19 were moving back to their main roles. As such, we struggled to get traction on the imperative to set-up the platform and move it onto the delivery phase. This was despite a rapid funding decision and fast ethics and MHRA approvals following IRAS submission. In the future, central and peripheral support for pandemic studies needs to be maintained whilst it is still considered important to run pandemic-related research. The issues of contracting and regulatory hurdles has similarly been raised by the RECOVERY and REMAP-CSAP investigators.^66^

### 4.5. Declining SARS-CoV-2 infection and COVID-19 disease rates in care homes

Although logistical issues as highlighted above impeded the platform’s progress, declining SARS-CoV-2 rates and disease severity in care homes meant that the trial lost its scientific purpose and would have required contracting and recruitment of unrealistic numbers of care homes and residents if it were to continue.

### 4.6. Closedown decision

The decision to close the platform prematurely was challenging to make but necessary because of the success of vaccination and significant reduction in care home SARS-CoV-2 related events. The decision was promoted by the Executive Committee and discussed and debated with:

- Trial, internally: Platform Steering Committee and Data Monitoring Committee (who both sent endorsing letters), Platform Steering Group (grant applicants).
- UoN, internally: Sponsor, School of Medicine, Faculty of Medicine and Health Sciences, Pro-Vice Chancellor for Research.
- Externally: Chief Medical Advisor, NIHR (funder), Department of Health and Social Care (DHSC).

With their support, a formal decision to close was made with NIHR. This decision was then shared with HRA/REC, MHRA, CRN/EnRICH, care homes, GPs, PIs, Insurer, Drug Companies, Central Pharmacy, and the EudraCT and ISRCTN registries. Since no care homes had been contracted or residents enrolled, no data were collected.

### 4.7. Design alternatives

#### Cluster versus individual randomisation

Having a cluster randomised rather than individual randomised trial has both advantages and disadvantages. Cluster designs are especially relevant to care home trials since they reduce the risk of bias due to contamination,^67^ allow ‘real world’ investigation of transmission, facilitate recruitment, enable simpler drug management and delivery, and ease identification/attribution of serious adverse events. Further, they most reflect the manner that prophylactic interventions will be used in care homes, i.e. for all residents following an outbreak. In view of these clear practical advantages especially in a research naïve environment in a pressurised situation, we elected to use a cluster trial design. Nevertheless, a cluster trial design leads to a much larger trial than using individual randomisation, in the case of PROTECT-CH by a factor of more than five (see sample size calculation in section 2.11.1 above). In retrospect and taking account of the problems of contracting with multitudinous care homes and GPs and recruiting many thousands of residents, some might argue that we should have used individual randomisation and accepted this lesser design; this would have required recruitment of only 1,000-2,000 residents from fewer than 100 care homes and is a case of “best [perfect] is the enemy of good” (Voltaire 1770). However, others would counter that such a design would not address the commissioned brief/research question and would therefore be unethical and tantamount to research waste.

#### Feasibility trial

The PROTECT-CH platform was designed to provide definitive evidence for preventing COVID-19 in care homes; as such it was large and complex. In preparing the trial, we solved numerous problems and demonstrated feasibility for multiple challenges, many highlighted above. However, by stopping the trial when we did, we were unable to test the feasibility of contracting with care homes, GPs, PIs and RNs; screening, consenting and treating residents; and obtaining central and care home data.

### 4.8. Strengths

The key strength is that the start-up phase supported the feasibility of obtaining regulatory approvals (MHRA, REC, HRA), developing the website, database and training materials, obtaining insurance and identifiers suppliers of routine data, central pharmacy support and engagement and collaboration between the four UK nations. Further, hundreds of care homes and staff were primed as to the importance and practice of clinical research.

Additionally, PROTECT-CH has added value to other projects and programmes:

#### Aberdeen Centre for Healthcare Randomised Trials

We asked them to adapt their site-tracking database for site approvals, green light documents, processes and trial staff training. We were using this system at close-down.

#### Air Filters to Prevent Respiratory Infections including COVID-19 in Care Homes (AFRI-c) trial

We were approached by the AFRI-c team to share information on care homes that had expressed interest in PROTECT-CH. We asked our ∼300 care homes that had expressed interest that if they might be interested in joining AFRI then they should contact them.

#### EnRICH Scotland (https://www.nhsresearchscotland.org.uk/research-in-scotland/facilities/enrich)

PROTECT-CH provided a focus around which the newly inaugurated EnRICH network in Scotland could become established, with funding from the Chief Scientist Office. This was not in place pre-pandemic. Weekly meetings were established with NCTU and the Scottish lead (Dr Shenkin) and other Scottish Co-Is (Prof Guthrie) and PIs and key Scottish stakeholders (NHS Research Scotland Central Management team, representative from NRS boards, NRS Primary Care Network, NRS Ageing Network to develop processes to deliver the trial. PROTECT-CH provided an opportunity for practical and focussed conversations with care homes, leading to their recruitment into the network. A bid, capitalising on this progress, and using SOPs from PROTECT-CH, is being collated by senior Scottish academics to start a trial of anticholinergic burden reduction, and has been submitted to the Scottish Chief Scientist Office (CSO).

### 4.9. Limitations

The main limitation is that the trial never recruited, treated or followed any care home residents so that the primary aim of testing post exposure prophylaxis with ciclesonide and niclosamide was never achieved.

### 4.10. Conclusions

PROTECT-CH was commissioned in quarter 4 2020 during the UK’s second wave of the COVID-19 pandemic in response to very high care home infection and death rates in early 2020, and prior to detailed knowledge about vaccine efficacy in older people. By the time it would have been ready to start contracting with care homes in quarter 2 2021, most residents and staff had been vaccinated and SARS-CoV-2 infection rates were very low. By the third quarter of 2021, the underlying research question of how to reduce infection rates and disease severity had become irrelevant. Assuming a decision by CMO, DHSC and NIHR to commission the platform in mid 2020, we do not believe that this commissioning decision nor the trial start-up were wasted; it only became clear in early to mid 2021 that COVID-19 rates in care homes were much lower and that vaccine take-up and efficacy would be high in older people. The key take home message is that any decision to set up a platform in care homes would need to be taken much earlier in any future pandemic.

PROTECT-CH would have been the world’s largest care home trial and was, as such, a trailblazer and achieved many successes in preparing to recruit care homes and then residents across all UK nations; however, the above impediments prevented further progress. We have shared, where possible, these successes though the trial website, GitHub and this report, and will expand on aspects of these in future publications. We believe that our solution for insurance and process for managing national routine data is relevant to following care home intervention trials.

#### Implications for healthcare

We did not recruit, treat or follow-up any randomised and treated residents and so there are no implications for the prevention of transmission of SARS-CoV-2 and reduction of COVID-19 severity in care homes.

#### Recommendations for research

Care home research is in its infancy and with an ageing population and vast numbers of research questions, it is vital that future trials become more feasible. Multiple recommendations arise, including optimising the manner of contacting between NIHR and commercial companies; streamlining the mechanism for contracting with large numbers of care homes, GPs, PIs and RNs; and improved support for trials that start later in any future pandemic so they may benefit from efficient processes and government, healthcare and social care support. A trial assessing the feasibility of contracting; and recruiting, treating and following up residents including taking account of differences in legislation, governance and health and social care infrastructure between UK nations, is still needed. Indeed, because of the considerable inertia required in setting up a care home platform trial, there is an argument that one should be maintained in hibernation so that the main initial activity in any future pandemic would be to start recruitment of residents rather than all the initial activities performed for PROTECT-CH in January-May 2021; this is analogous to the HTA-funded Nottingham-based adjuvant steroids in adults admitted to hospital with pandemic influenza (ASAP) trial.^68^

Much of the apparatus developed during PROTECT-CH could be repurposed rapidly, in the event of a future pandemic, to conduct rapid evaluations of prophylactic, or even treatment, agents in the care home setting. Early identification of this as a research priority would enable a research team to establish a platform more rapidly than we were enabled to do during PROTECT-CH. It would be reasonable to include materials and protocols from PROTECT-CH when considering pandemic preparedness of the research infrastructure as a whole, and to iterate these as pandemic preparedness is developed.

Between pandemics, the work undertaken during PROTECT-CH emphasises what has long been recognised in the care home research community, which is that establishing trial infrastructure in this setting is labour intensive and complex. This results in considerable inefficiencies when trial infrastructure is built, dismantled, and then rebuilt again, for each study that NIHR wants to fund in the sector. This is in contrast to hospitals, and NHS community organisations, where expertise and capacity for research is retained between studies through ongoing infrastructure investment. Existing endeavours to render the care home setting “research-ready” (EnRICH) have largely focussed on raising awareness of research within the care home sector and building links with care home organisations and staff. These approaches haven’t, however, built organisational capacity to do research on a regular basis or at short notice. The SOPs developed during PROTECT-CH could provide the foundation for a more sustainable and strategic research infrastructure in the sector. Against this would be longstanding issues with staff turnover, and hence loss of expertise over time, and the challenges of engaging with a sector facing historically unprecedented challenges around funding and capacity. There is though, potential, to test the feasibility of such infrastructure based upon the work that we have started here.

## Supporting information

Appendix 2. Statistical Analysis Plan

Appendix 3. Health Economic Analysis Plan

## Data Availability

The trial never recruited patients so no data are available.

https://www.protect-trial.net

## Declared competing interests

1. Philip M Bath: Grants from British Heart Foundation and NIHR Health Technology Appraisal programme during the conduct of the study; personal fees from DiaMedica and Phagenesis outside the submitted work.
2. Jonathan Ball: Reports no competing interests.
3. Matthew Boyd: Personal fees from Delphi Healthcare outside the submitted work.
4. Heather Gage: Reports no competing interests.
5. Matthew Glover: Reports no competing interests.
6. Maureen Godfrey: Reports no competing interests.
7. Bruce Guthrie: Grants from NIHR, Medical Research Council, Legal and General PLC, and Chief Scientist Office outside the submitted work; and is a member of the SAGE Social Care Workgroup.
8. Jonathan Hewitt: Funding from Health and Social Care Research Wales (HSCRW) during the grant.
9. Robert Howard: Grants from NIHR Health Technology Appraisal, Medical Research Council and NIHR UCLH Biomedical Research Centre.
10. Thomas Jaki: Grants from NIHR and MRC during conduct of the study.
11. Edmund Juszczak: Grants from NIHR during conduct of the study.
12. Dan Lasserson: Grants from NIHR HTA Clinical Evaluation and Trials, NIHR Policy Research Programme, NIHR RfPB, NIHR ARC and MIC funding schemes, institutional funding for quality improvement studies from Vifor Pharma outside of the current work.
13. Paul Leighton: Grants from NIHR during conduct of the study.
14. Val Leyland: Reports no competing interests.
15. Wei Shen Lim: Grants from NIHR and my institution has received unrestricted investigator-initiated research funding from Pfizer for a study in which WSL is CI, during the conduct of the study.
16. Pip Logan: Grants from NIHR Health Technology Appraisal, NIHR RfPB.
17. Garry Meakin: Reports no competing interests.
18. Alan A Montgomery: Member NIHR HTA Clinical Evaluation and Trials Funding Committee; grants from NIHR during conduct of the study.
19. Reuben Ogollah: Grants from NIHR during conduct of the study.
20. Peter Passmore: Reports no competing interests.
21. Phil Quinlan: Reports no competing interests.
22. Caroline Rick: Grants from NIHR during conduct of the study.
23. Simon Royal: Reports no competing interests.
24. Susan D Shenkin: Grants from British Geriatrics Society, NIHR, Edinburgh Lothian Health Foundation, Marie Curie and Chief Scientist Office during the conduct of the study.
25. Clare Upton: Reports no competing interests.
26. Adam L Gordon: Grants from NIHR, Asthma UK, the British Lung Foundation, the Wellcome Trust and British Geriatrics Society during the conduct of this research; and is a member of the SAGE Social Care Workgroup.

## LIST OF ABREVIATIONS

acOR: adjusted common odds ratio
aHR: adjusted hazards ratio
aMD: adjusted mean difference
BLR: binary logistic regression
BNF: British National Formulary
CI: chief investigator
CMO: chief medical officer
CPHR: Cox proportional hazards regression
CRN: clinical research network
cOR: common odds ratio
DHSC: Department of Health and Social Care
DMC: data monitoring committee
EnRICH: Enabling Research in Care Homes
EQ-5D-5L: EuroQol-5 dimensions-5 levels
EQ-VAS: EuroQol-Visual Analogue Scale
eTMF: electronic Trial Master File
HR: hazard ratio
HRA: Health Research Authority
IMP: investigational medicinal product
IQR: interquartile range
LFT: lateral flow test
MAR: medicine administration record
MD: mean difference
MHRA: Medicines and Healthcare products Regulatory Agency
MLR: multiple linear regression
NC: not calculated
NICE: National Institute for Health and Care Excellence
NIHR: National Institute for Health Research
NS: not significant
OLR: ordinal logistic regression
OR: odds ratio
OTR: onset to randomisation
P: Probability
PCN: primary care network
PCR: polymerase chain reaction
PEP: post-exposure prophylaxis
PI: principal investigator
PLR: personal legal representatives
PPI: patient-public involvement
PrEP: pre-exposure prophylaxis
PSC: platform steering committee
QP: qualified person
REC: research ethics committee
RRR: relative risk reduction
SAE: serious adverse event
SCR: summary care record
SD: standard deviation
SoECAT: schedule of events cost attribution template
SOP: standard operating procedure
TA: technology appraisal (by NICE)
TRE: trusted research environment
UoN: University of Nottingham
XP: cross programme (NIHR-hosted)

## Plain English Summary

The COVID-19 pandemic has devastated care homes through causing illness and death and by limiting freedom of movement and family visits. In the first wave of the pandemic in the UK, 40% of COVID-19 related deaths happened in care home residents. Internationally by October 2020, almost 50% of COVID-19 deaths happened in care home residents or similar facilities in other countries. Older people are at high risk of complications from COVID-19 illness due to the presence of other illnesses and an ageing immune system. At the time of starting this work, it was not clear how effective vaccination programmes for care home residents would be, and there were no other effective treatments that could reduce the risk of catching or becoming unwell with COVID-19.

We set up a large “platform” trial to test treatments that might reduce the spread of SARS-CoV-2, the virus that causes COVID-19, within care homes and reduce the severity of COVID-19 with fewer people being admitted to hospital or dying. A trial platform allows multiple treatments to be tested at the same time, or one after the other. The results are analysed regularly, and as soon as a treatment is shown to be effective or ineffective, it is taken out of the trial. This makes space for new treatments to be added to the trial, and for treatments that work to be recommended for all suitable people. This process of testing treatments and then replacing them with new treatments can go on for many months or years. Government advisors chose two drugs for us to test: inhaled ciclesonide (a drug used to prevent asthma by reducing inflammation) and intranasal niclosamide (a drug used to treat hookworm). We aimed to recruit more than 750 care homes for older people from across the UK and, from them, approximately 24,000 residents. Following an outbreak of COVID-19 (classed as a single positive test by any member of staff or resident) care homes would be randomised (like a toss of a coin) to the drugs or control. Treatment was to be taken for six weeks and residents followed for a total of four months. We would also study whether the treatments were cost-effective and the best ways of setting-up and delivering the trial.

We developed a trial protocol – a description of how the trial would run - which included information sheets for residents or their personal legal representatives, training materials (including videos and audio descriptions) for care home staff, protocols to recruit and randomise residents with and without mental capacity, and ways of storing data safely, securely and ethically. A national network of doctors and trial specialists was planned to support the project including consent of residents living with dementia, together with a website, ways to collect routinely available clinical data to measure outcomes, and contracts for a central pharmacy to arrange drug management were in development. Much of this work was supported by an embedded Patient Public and Carer team. However, problems with contracting for the drugs and obtaining insurance meant the trial was delayed by more than five months and the exceptional benefit of vaccination meant that care home infections and the number of residents affected by COVID-19 fell. Following widespread consultation, a decision was made to close the platform in September 2021 with the grant ceasing in December 2021.

The trial was run from the University of Nottingham with support by the Universities of Cambridge, Edinburgh, Surrey and Warwick, and University College London and involved an experienced team of doctors, nurses, allied health professionals, statisticians, trial methodologists, health economists and public partners, all with considerable experience in care home research and running trials and platforms.

This report describes the trial platform, problems in delivery and learnings from the numerous challenges. We have made many of the deliverables available via the trial website for others to review, develop and improve.

## Scientific Summary

The COVID-19 pandemic devastated care homes through causing illness and death and by limiting family visits. In the first UK wave of the pandemic, 40% of COVID-19 related deaths occurred in care home residents; internationally by October 2020, almost 50% of COVID-19 deaths happened in care homes (or long-term care facilities). Older people are at high risk of complications from COVID-19 illness due to the presence of other illnesses and an ageing immune system. Beyond public health measures to prevent infection (hygiene, masks, personal protective equipment, maintaining distance), treatments are urgently needed to minimise these direct and indirect impacts on residents. At the time the trial was designed, which was before mass vaccination programmes, only two treatments, steroids and antibodies, had been shown to reduce death due to COVID-19 but these only work in people with severe disease such as those who need oxygen.

We set up a large platform trial to test treatments that might reduce the spread of SARS-CoV-2 within care homes and reduce the severity of COVID-19 with reduced hospitalisation and death. Government advisors chose two drugs for us to test: inhaled ciclesonide and intranasal niclosamide. We aimed to recruit more than 750 care homes for older people from across the UK and, within them, approximately 24,000 residents. Following an outbreak of COVID-19 (classed as a single positive test by any member of staff or resident), care homes would be randomised to ciclesonide, niclosamide or a control group. Treatment would be taken for six weeks, and residents followed up for a total of four months. The primary ordinal outcome comprised no infection, SARS-CoV-2 infection without admission to hospital, all-cause hospitalisation and all-cause death. Secondary outcomes included time to events, causes for hospitalisation and death, frailty, and health economic and process evaluations.

We developed a trial protocol and information sheets for residents, training materials including videos and audio descriptions for care home staff, a regular training and information sharing forum for care home staff, a clinical database and secure data repository, a website, a thirty strong team of principal investigators across all four nations and arranged to use routinely collected (national) clinical data and for a central pharmacy to arrange drug management. Much of this work was supported by an embedded Patient Public and Carer team. However, problems with contracting for the drugs and obtaining insurance meant the trial was delayed by more than five months and the exceptional benefit of vaccination meant that care home infections and the number of residents affected by COVID-19 fell dramatically. The resulting effect on the sample size required to answer the research question (increasing from 750 to several thousand care homes and from 24,000 to more than 100,000 participants) meant that the study was no longer feasible. Following widespread consultation, a decision was made to close the platform in September 2021 with the grant ceasing in December 2021. This was before the emergence of the Omicron variant, the impact of which on care homes is uncertain at the time of writing.

The trial was run from the University of Nottingham Clinical trials Unit with support by the Universities of Cambridge, Edinburgh, Surrey and Warwick, and University College London and involved an experienced team of doctors, statisticians, trial methodologists, health economists and public partners, all with considerable experience in care home research and running clinical trials and platforms.

This report describes the trial platform, problems in delivery and learnings from the numerous challenges. We have made many of the deliverables available via the trial website (<u>protect-trial.net)</u> for others to review, develop and enhance.

## Trial registration and identifiers

- EudraCT: 2021-000185-15
- IRAS project ID: 294832
- NCTU reference number: 2033
- NIHR grant reference: NIHR133443
- Sponsor reference/protocol number: 21001
- Twitter: @ProtectTrial
- Web url: https://www.protect-trial.net
- WHO universal trial no: UTN U1111-1265-4068

## Chapter 5: Acknowledgements

We thank the participants, investigators and research staff at the participating sites and members of the Platform Steering Committee and Independent Data Monitoring Committee, for their involvement in and support for, the study. PROTECT-CH thanks the National Institute for Health Research Clinical Research Network and NHS Research Scotland for their support for the platform.

### 5.1. Contribution of authors

- Philip Bath (Stroke Association Professor of Stroke Medicine, Director Stroke Trials Unit, Emeritus NIHR Senior Investigator): Chief Investigator, Grant applicant and member of Platform Management and Executive Committee. PMB is guarantor for the study and wrote the first draft of the manuscript.
- Jonathan Ball (Professor of Virology, University of Nottingham): Grant applicant.
- Matthew Boyd (Associate Professor of Patient Safety and Pharmacy Practice, University of Nottingham). Trial Pharmacist
- Heather Gage (Professor of Health Economics, University of Surrey): Grant applicant and lead health economics.
- Matthew Glover (Senior Health Economist, University of Surrey): Grant applicant and member health economics group.
- Maureen Godfrey (Patient and Public Representative): Grant applicant and member of PPI group.
- Bruce Guthrie: Contributed to study design, study implementation in Scotland, and the drafting of the final report.
- Jonathan Hewitt (Clinical Senior Lecturer, Cardiff University): Co-Applicant and Lead for PROTECT-CH in Wales.
- Robert Howard (Professor of Old Age Psychiatry, UCL). Grant applicant and member of the PI group.
- Thomas Jaki (Professor of Statistics, University of Cambridge). Grant applicant and member of the Platform Management committee and the Analysis Group.
- Edmund Juszczak (Professor of Clinical Trials and Statistics in Medicine, Nottingham Clinical Trials Unit). Grant applicant and member of Platform Management and Executive Committee, eCRF, Analysis (statistics and health economics) and Website Groups.
- Dan Lasserson (Professor of Acute Ambulatory Care, university of Warwick): Grant applicant and member of the PI group, Lead for Acute Hospital at Home related drug delivery.
- Paul Leighton (Associate Professor of Applied Health Research, University of Nottingham). Grant co-applicant and member of the Platform Management Group. Lead for Process Evaluation Group.
- Val Leyland (Patient and Public Representative): Member of PPI group.
- Wei Shen Lim (Honorary Professor of Medicine, Consultant Respiratory Physician, Nottingham University Hospitals NHS Trust) Grant co-applicant.
- Pip Logan, Professor of Rehabilitation Research and Occupational Therapist, Grant Holder and member of the Platform Management Group and PPI group.
- Garry Meakin: (Trial Manager, University of Nottingham): Trial Manager.
- Alan A Montgomery (Professor of Medical Statistics and Clinical Trials, Director Clinical Trials Unit). Grant applicant and member of Platform Management and Executive Committee, and Website Groups.
- Reuben Ogollah (Associate Professor of Medical Statistics, Nottingham Clinical Trials Unit): Member Statistical Analysis Group.
- Peter Passmore (Professor of Ageing and Geriatric Medicine, Queen’s University Belfast): Grant applicant, N Ireland Lead, member of PI group.
- Phil Quinlan (Honorary Professor and Director of Health Informatics, University of Nottingham): Grant applicant, Lead Routine data.
- Caroline Rick (Associate Professor of Clinical Trials) Grant co-applicant and member of the platform management and executive committee and the website, eCRF, trials management, patient and public involvement and Process evaluation groups.
- Simon Royal (General practitioner/Research Lead University of Nottingham Health Service). Grant applicant, member PI group.
- Susan D Shenkin (Reader in Ageing and Health; co-chair of EnRICH Scotland) Grant applicant, Scottish lead, member of PI group.
- Clare Upton (Trial Manager, University of Nottingham): Trial Manager.
- Adam L Gordon (Professor of Care of Older People, University of Nottingham). Co-Chief Investigator, Grant applicant, member of Platform Management and Executive Committee, Lead for PI and Care Home forums.

### 5.2. Funding

PROTECT-CH was funded by the NIHR Cross Programme (grant NIHR133443, 1 January 2021 to 17th December 2021). This paper presents independent research funded by the NIHR. The views expressed are those of the author(s) and not necessarily those of the NHS, the NIHR or the Department of Health and Social Care. Although intended, there was no commercial support for the trial. The trial was sponsored by the University of Nottingham.

### 5.3. Data sharing

No research data were generated. Access to components of the REDCap database are available via GitHub at:

https://github.com/Nottingham-CTU/PROTECT-CH-TRIAL

Access to training materials and other trial documents are available via the trial website at: https://www.protect-trial.net

All queries and data requests should be submitted to the corresponding author for consideration in the first instance.

## APPENDICES

## APPENDIX 1. STUDY INVESTIGATORS

### Writing Committee

Philip M Bath, Jonathan Ball, Matthew Boyd, Heather Gage, Matthew Glover, Maureen Godfrey, Bruce Guthrie, Jonathan Hewitt, Robert Howard, Thomas Jaki, Edmund Juszczak, Daniel Lasserson, Paul Leighton, Val Leyland, Wei Shen Lim, Pip Logan, Garry Meakin, Alan Montgomery, Reuben Ogollah, Peter Passmore, Philip Quinlan, Caroline Rick, Simon Royal, Susan D Shenkin, Clare Upton, Adam L Gordon; on behalf of the PROTECT-CH Trialists.

### Platform Steering Committee (independent)

*Independent members:* Alistair Burns (Manchester, Chair), Shaun Treweek (Aberdeen), James Wason (Newcastle), Anne Forster (Leeds), Azhar Farooqi (Leicester), Martin Vernon (Manchester), Peter Pratt (NHS England), Vic Rayner (National Care Forum).

*Grant holder:* Philip Bath (Nottingham, Chief Investigator)

*Sponsor’s representative:* Angela Shone (University of Nottingham)

### Data Monitoring Committee (independent)

Kennedy Lees (Glasgow, Chair), Tim Peters (Bristol), Ben Carter (London), Finbarr Martin (London), Eileen Burns (Leeds)

### NIHR Grant Applicants

Philip M Bath (Nottingham), Adam Gordon (Nottingham), Thomas Jaki (Cambridge), Cally Rick (Nottingham), Alan Montgomery (Nottingham), Edmund Juszczak (Nottingham), Pip Logan (Nottingham), Wei Shen Lim (Nottingham), Simon Royal (Nottingham), Heather Gage (Guildford), Bruce Guthrie (Edinburgh), Dan Lasserson (Warwick), Rob Howard (London), Matthew Glover (Guildford), Maureen Godfrey (Nottingham)

### Executive Committee

Philip M Bath, Adam Gordon, Alan Montgomery, Edmund Juszczak, Cally Rick.

### Nottingham Clinical Trials Unit

*Senior Trial Managers:* Senior Trial Managers: Kathryn Starr, Clare Brittain

*Trial Manager:* Garry Meakin, Clare Upton, Sarah Craig, Jenny Riga, Lindsay Armstrong-Buisseret, Katherine Belfield, Eleanor Harrison, Sarah Norton

*Trial Coordinators:* Damini Mistry, Leila Thuma, Wendy Daunt, Joanne Brooks

*Statisticians:* Reuben Ogollah, Chris Partlett, Lucy Bradshaw

*Programming/database management:* Nicholas Hilken, Nina Carlisle, Richard Dooley, Chien May

*Data managers*: Stella Tarr

*Quality Assurance:* Melanie Boulter, Lynsey Burns

*Finance:* Charlotte Curnow

*Administrator*: Matea Svigac

### Nottingham Stroke Trials Unit

*Senior Trial Manager:* Diane Havard

*Programmer:* Lee Hayward

### University of Nottingham

*Media:* Charlotte Anscombe

*National data:* Phil Quinlan, Linda Fiaschi

### Public, Patient Involvement

*Public:* Maureen Godfrey, Val Leyland

*Professional support:* Kirsty Sprange, Katie Robinson, Katie Belfield, Lindsay Armstrong-Buisseret, Wendy Daunt, Cally Rick

### Principal Investigators

Asangaedem Akpan, Liverpool University Hospitals NHS Foundation Trust

Jay Amin, Southern Health NHS Foundation Trust

Susan Angus, NHS Tayside

Philip Bath, Nottingham University Hospitals NHS Trust

Andrew Clegg, Bradford Teaching Hospitals NHS Foundation Trust

Graham Devereux, Aintree University Hospital

Serge Engamba, UEA Medical Centre

Adam Gordon, University Hospitals of Derby and Burton NHS Foundation Trust

Amy Heskett, Kent Community Health NHS Foundation Trust

Jonathon Hewitt, Aneurin Bevan University Health Board

Tania Kalsi, Guy’s and St Thomas’ NHS Foundation Trust

Daniel Lasserson, Oxford University Hospitals NHS Foundation Trust

Farhana Lockat, Manor Court Surgery, Nuneaton

Sarah Mitchell, Sheffield Teaching Hospitals NHS Foundation Trust

Elizabeta Mukaetova-Ladinska, University Hospitals of Leicester NHS Trust

David Mummery, North End Medical Centre,

Aditya Nautiyal, Nene Valley Hodgson Practice

Shelagh O’Riordan, Kent Community Health NHS Foundation Trust

Peter Passmore, Belfast Health and Social Care Trust

Terence Quinn, NHS Greater Glasgow and Clyde Board

Susan Shenkin, NHS Lothian

David Smithard, Lewisham and Greenwich NHS Trust

Divya Tiwari, University Hospitals Dorset NHS Foundation Trust

Emma Vardy, Salford Royal NHS Foundation Trust

Tomas Welsh, Royal United Hospitals Bath NHS Foundation Trust

Miles Witham, The Newcastle upon Tyne Hospitals NHS Foundation Trust

## APPENDIX 2. STATISTICAL ANALYSIS PLAN (SAP)

## Appendix 3. HEALTH ECONOMIC ANALYSIS PLAN

## APPENDIX 4. EVALUATION OF PROTECT-CH DEVELOPMENT

### Introduction

Despite PROTECT-CH being stopped prior to contracting of care homes there is still potential to gain valuable insight about its development which can inform future research endeavour.

Here we describe a brief on-going process evaluation which seeks to establish further lessons that can be gained from the development of the PROTECT-CH trial protocol and research infrastructure.

This work is distinct from, and separate to, the process evaluation sub-study described previously which was intended to run nested within the platform.

### Background

Despite an establishedlllliterature on the RCT method,^69, 70^ the publication of RCT protocols and reportinglllguidelines for RCTs ^71^ parts of the RCT process remain a black box.lllMaking explicit what happens in the set-up and operation of clinical trialslllmay support more efficient trial delivery. Given recent concerns for efficiencies in clinical research ^72^ it is timely to investigate this.lll

The emergence of platform trials as a more efficient trial design adds to the timeliness of this evaluation. As does the recognition that platform trials exhibit a number of inherent challenges and difficulties (https://www.ctu.mrc.ac.uk/news/news-stories/2019/may/the-challenges-of-running-platform-trials/).

More specifically, regarding care home research, COVID-19 has emphasised the central role that care homes play in health and social care of an aging population.lllHowever, research in care homes is inherently challenging and under-reported.^73^ The authors have previously recognised these challenges – ranging from a naivety about research processes to practical challenges associated with collecting data in a ‘domestic’ (rather than clinical) setting.^67, 74^

Beyond this the PROTECT-CH trial faced challenges in its size (one of the largest care home trials ever proposed), its scope (CTIMP trials are uncommon in care homes), and in the required speed of set-up (conceivably protecting a vulnerable population during a global pandemic).

That PROTECT-CH has not entered the trial delivery phase allows us the resource to reflect upon how it was conceived, operated and what it has achieved. This is a unique opportunity to better understand care home trials and/or research during a global pandemic. It is opportunity to identify lessons that others might put into operation when working in challenging circumstances, or when trying to instigate a step-change in social care research.

### Aims and Objectives

The aim of this evaluation is to provide insight about the set-up of a platform trial in care homes. It will identify those factors (contextual and ways of working) which support this process, and those which inhibit trial set-up.

### Objectives include

1. To map the organisational landscape of PROTECT-CH - who was involved? In what way? How were aspects of PROTECT-CH connected?
2. To establish key substantive issues related to rapid, platform trial set-up and researching in care homes – what worked well? What was achieved? What barriers were experienced?
3. To generate detailed and specific insight about operational, strategic, and organisational matters – to consider how things might have been done differently. To identify those internal and external factors which were key enablers or barriers.

### Evaluation Plan

This evaluation is conceived in three parts.

#### 1. PROTECT-CH INFRASTRUCTURE

To establish the organisational landscape of PROTECT-CH the membership and minutes of those working groups and strategic groups identified in the initial PROTECT-CH organogram will be reviewed.

**Appendix 4, Figure 1.**
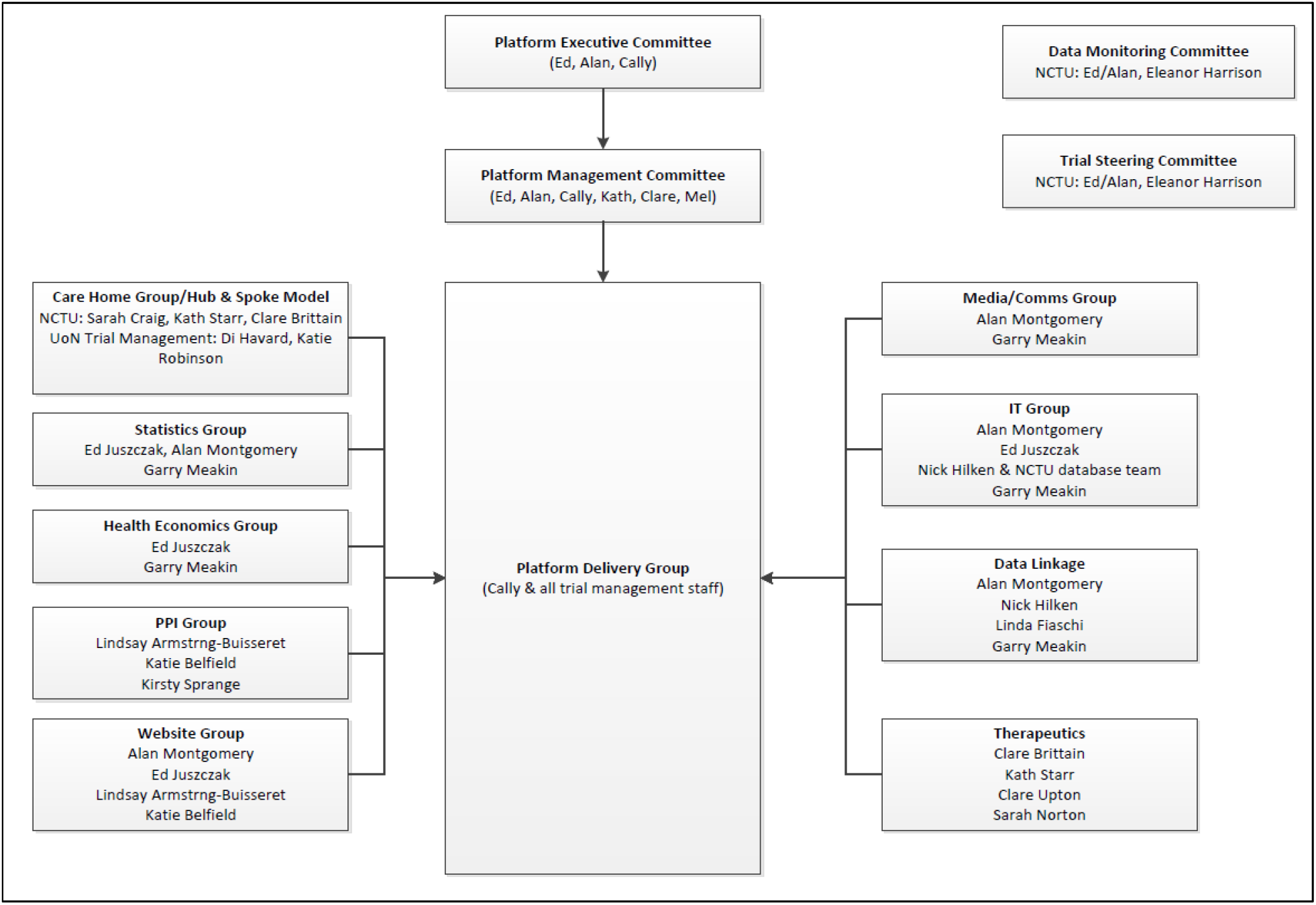
Initial PROTECT-CH Organogram.

Data review will demonstrate the number of individuals involved in the PROTECT-CH set-up, the range of professions contributing to the working groups and will demonstrate how this network of groups are effectively organised and connected. It will demonstrate the contribution of local (Nottingham University and NCTU) and national collaborators and will consider the multi-disciplinary nature of the PROTECT-CH team.

Data review will identify key stakeholders who are involved in multiple PROTECT-CH working and strategic groups.

#### 2. WORKING GROUP SURVEY

The administrative chair of each PROTECT-CH working group will be asked to complete a short, online survey which captures the challenges and successes of each element of the PROTECT-CH set-up. The survey will consist primarily of open-response questions which consider: the purpose of the working group, challenges facing the working group, successes, and key learning.

Responses will be summarised to populate a simple, descriptive analytic table.

**Appendix 4, Table 1.**
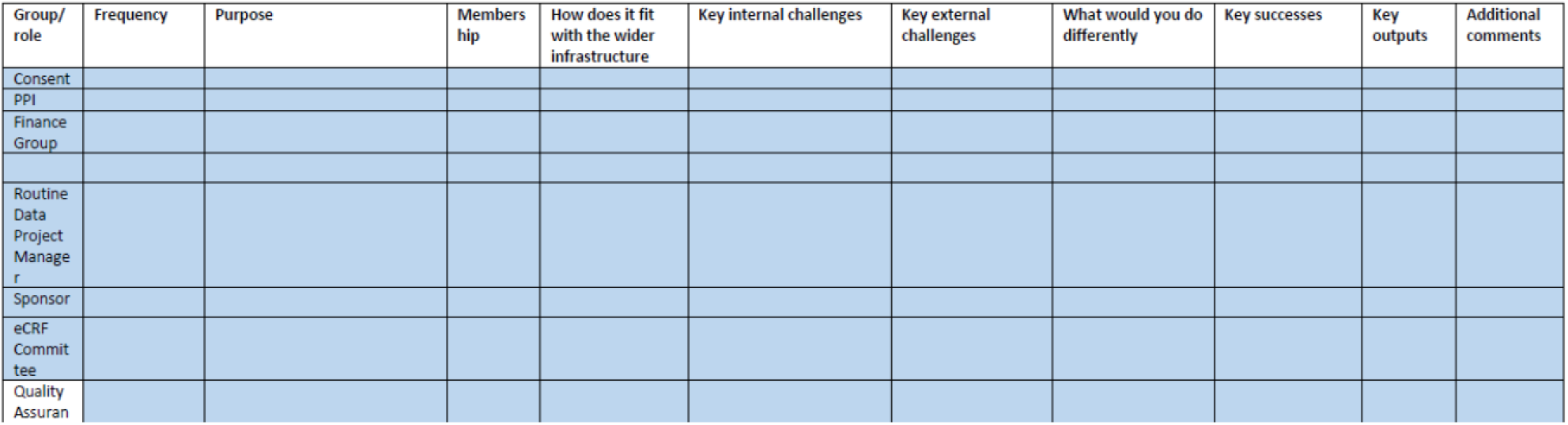
Summary of evaluation survey responses.

Data in the table may be summarised horizontally (to reflect the experience of an individual working groups) or vertically (to capture recurrent concerns or patterns across multiple working groups).

#### 3. INTERVIEWS WITH KEY STAKEHOLDERS

Further detail on the successes and challenges of developing PROTECT-CH will be sought in one-to-one interviews with key stakeholders. These will be selected from the membership of the PROTECT-CH working and executive groups and will include all original co-applicants.

Interviews will consider similar ground to the online-survey, but will allow more detailed exploration of pertinent issues, they will allow key stakeholders to share their novel perspective on PROTECT-CH. Interviews will explicitly consider how PROTECT-CH has related to external organisations and navigated the challenge of setting up research in multiple care home settings. The interviews will generate key learning about: (i) rapid, set-up of platform trials; and (ii) the challenges of delivering prophylactic intervention in care homes.

Data will be analysed following the conventions of framework analysis ^75^ with initial analytical tables informed by the MRC guidelines on process evaluation in the development and evaluation of complex interventions.^76^ Analysis will consider the context of PROTECT-CH development, the implementation of development processes, mechanisms triggered by PROTECT-CH actions, and the outcome of PROTECT-CH development stage.

**Appendix 4, Table 2.** Example of a provisional analytic framework.

| CONTEXT |  |  |  |  |
| --- | --- | --- | --- | --- |
|  | Care home research | Platform design | COVID circumstances | Other .... |

### Progress to Date

Ethical approval for this work has been received from the University of Nottingham, Faculty of Medicine and Health Sciences Research Ethics Committee – reference FMHS 333-0921 (29^th^ September 2021).

Work has commenced on all aspects of the evaluation.

## APPENDIX 5. PRESS, MEDIA AND SOCIAL MEDIA OUTPUTS

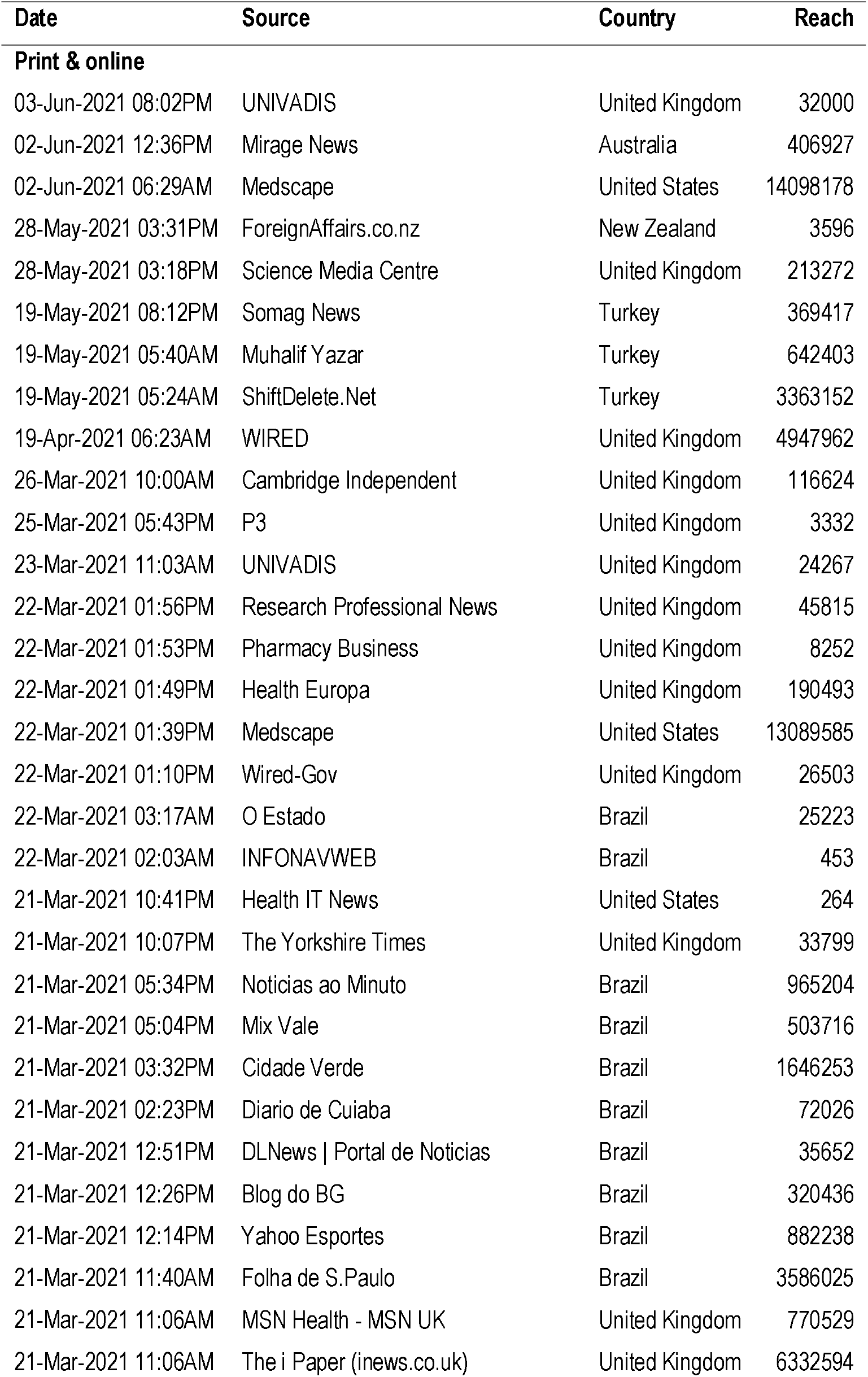
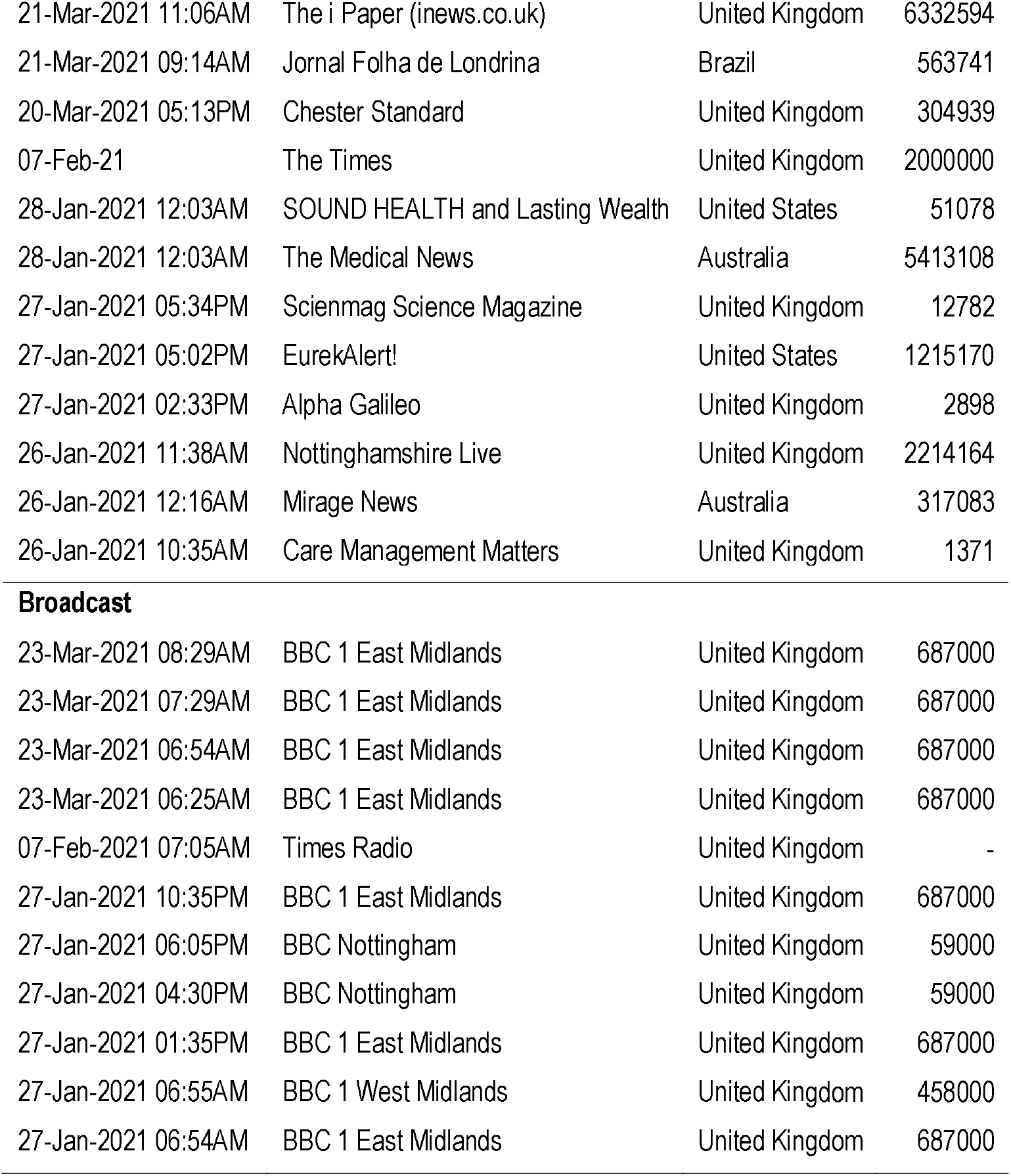

## APPENDIX 6. CARE HOME CONTINUING PROFESSIONAL DEVELOPMENT WEBINARS

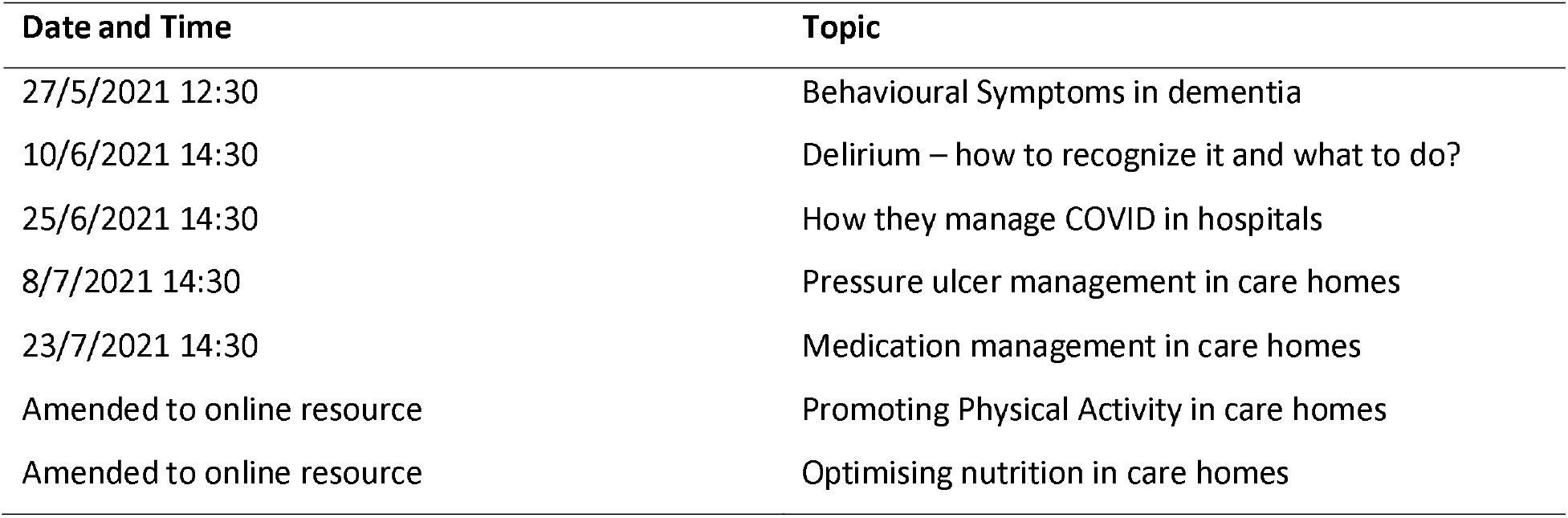

| **Date and Time** | **Topic** |
| --- | --- |
| 27/5/2021 12:30 | Behavioural Symptoms in dementia |
| 10/6/2021 14:30 | Delirium – how to recognize it and what to do? |
| 25/6/2021 14:30 | How they manage COVID in hospitals |
| 8/7/2021 14:30 | Pressure ulcer management in care homes |
| 23/7/2021 14:30 | Medication management in care homes |
| Amended to online resource | Promoting Physical Activity in care homes |
| Amended to online resource | Optimising nutrition in care homes |

