## Appendix 2. Statistical Analysis Plan for "Prophylactic Treatment of COVID-19 in Care Homes Trial (PROTECT-CH)"

### Prophylactic Therapy in Care Homes Trial

### Statistical Analysis Plan

Final version 1.0
(08 Oct 2021)

Based on Protocol version 2.0 (dated 01 July 2021)
Trial registration: EudraCT number 2021-000185-15
UTN: U1111-1265-4068

**Approval**

| **Name** | **Job title** | **Trial Role** | Signature | Date |
|---|---|---|---|---|
| Philip Bath | Professor of Stroke Medicine | Chief Investigator | [signature] | 20/10/2021 |
| Ed Juszczak | Professor of Clinical Trials and Statistics in Medicine | Senior Trial Statistician | [signature] | 11/10/2021 |
| Alistair Burns | Professor of Old Age Psychiatry | Chair of the Platform Steering Committee | [signature] | 21.10.2021 |

#### List of authors and reviewers

##### Author

Reuben Ogollah – Associate Professor of Medical Statistics and Clinical Trials, NCTU

##### Reviewers

Lucy Bradshaw – Medical Statistician, NCTU

Christopher Partlett – Assistant Professor of Medical Statistics and Clinical Trials, NCTU

Martin Law – Research Associate, MRC Biostatistics Unit, University of Cambridge

Thomas Jaki – Professor of Statistics, Lancaster University & MRC Biostatistics Unit, University of Cambridge

Ed Juszczak – Professor of Clinical Trials and Statistics in Medicine, NCTU

Alan Montgomery – Professor of Medical Statistics and Clinical Trials, NCTU

Tim Peters – Professor of Primary Care Health Services Research, University of Bristol, Data Monitoring Committee (DMC) independent statistician

Ben Carter, Senior Lecturer in Biostatistics, King's College London, DMC independent statistician

James Wason – Professor of Biostatistics, Newcastle University, PSC independent statistician

**Table of Contents**

1. INTRODUCTION & PURPOSE ... 6
2. SYNOPSIS OF STUDY DESIGN AND PROCEDURES ... 7
 2.1. Sample size and justification ... 11
 2.2. Blinding and breaking of blind ... 11
 2.3. Trial committees ... 11
 2.4. Outcome measures ... 11
3. INTERIM ANALYSIS ... 16
4. GENERAL ANALYSIS CONSIDERATIONS ... 16
 4.1. Framework ... 16
 4.2. Analysis sets ... 16
 4.3. Timing of primary analysis ... 17
 4.4. Statistical software ... 17
 4.5. Procedures for missing data ... 17
5. DESCRIPTION OF PARTICIPANT CHARACTERISTICS ... 17
 5.1. Participant flow ... 17
 5.2. Baseline characteristics ... 18
6. ASSESSMENT OF STUDY QUALITY ... 19
 6.1. Randomisation ... 19
 6.2. Adherence ... 19
 6.3. Follow-up and discontinuations ... 19
 6.4. Protocol deviations ... 20
7. ANALYSIS OF EFFECTIVENESS ... 20
 7.1. Primary analysis ... 20
 7.2. Sensitivity analysis of primary outcome ... 20
 7.3. Subgroup analysis of primary outcome ... 21
 7.4. Secondary outcomes ... 21
 7.5. Supplementary analysis of primary outcome and key secondary outcomes ... 22
8. ANALYSIS OF SAFETY ... 22
 8.1. Serious adverse events (SAEs) ... 22
 8.2. Adverse events relevant to the intervention ... 22
9. FINAL REPORT TABLES AND FIGURES ... 22
10. REFERENCES ... 24
11. Appendix ... 25
 11.1. SARS-CoV-2 infection scenarios after a test in PROTECT-CH ... 25

#### Abbreviations

| Abbreviation | Description |
|---|---|
| **AE** | Adverse Event |
| **AR** | Adverse Reaction |
| **CH** | Care home |
| **COVID-19** | Coronavirus-induced disease-19 |
| **DMC** | Data monitoring committee |
| **HRQoL (EQ-5D)** | Health-related quality of life instrument |
| **LFT** | Lateral flow test |
| **PCR** | Polymerase Chain Reaction |
| **PEP** | Post-exposure prophylaxis |
| **POG** | Prophylaxis Oversight Group |
| **PrEP** | Pre-exposure prophylaxis |
| **PSC** | Platform Steering Group |
| **PROTECT-CH** | PROphylactic ThErapy in Care homes Trial |
| **RIS** | Relative Information Sheet |
| **SAE** | Serious Adverse Event |
| **SAP** | Statistical Analysis Plan |
| **SAR** | Serious Adverse Reaction |
| **SARS-CoV-2** | Severe acute respiratory syndrome coronavirus 2. |
| **SUSAR** | Suspected Unexpected Serious Adverse Reaction |
| **TMG** | Trial Management Group |

**Changes from protocol**

The table below details changes to the planned analyses in the SAP compared to the protocol which after discussion with the TMG are not considered to require a protocol amendment.

| Protocol version and section | Protocol text | SAP version and section | SAP text | Justification |
|---|---|---|---|---|

**Amendments to versions**

| Version | Date | Change/comment | Statistician |
|---|---|---|---|

**Additional contributors to the SAP (non-signatory)**

| Name | Trial role | Job Title | Affiliation |
|---|---|---|---|

### 1. INTRODUCTION & PURPOSE

This document details the rules proposed and the presentation that will be followed, as closely as possible, when analysing and reporting the main results from the PROTECT-CH trial. The content of this document adheres to the guidelines for the content of statistical analysis plans in clinical trials [1].

The purpose of the plan is to:

1. Ensure that the analysis is appropriate for the aims of the trial, reflects good statistical practice, and that interpretation of a priori and post hoc analyses respectively is appropriate.

2. Explain in detail how the data will be handled and analysed to enable others to perform or replicate these analyses.

Additional exploratory or auxiliary analyses of data not specified in the protocol may be included in this analysis plan.

This analysis plan will be published on the study website, hence will be available publicly. It will be uploaded with the main papers when they are submitted for publication. Additional analyses suggested by reviewers or editors will be performed if considered appropriate. This should be documented in a file note.

Any amendments to the statistical analysis plan after unblinding of the statisticians producing DMC reports will be made by the blinded statisticians and will be described and justified.

Health economic and qualitative analysis plans are beyond the scope of this document.

#### 2. SYNOPSIS OF STUDY DESIGN AND PROCEDURES

| | |
|---|---|
| **Title** | **PROphylactic ThErapy in Care homes Trial** |
| **Acronym** | PROTECT-CH |
| **Short title** | PROTECT-CH |
| **Chief Investigator** | Professor Philip Bath |
| **Research Question** | In residents in a UK care home setting (P), which drug or antibody interventions (I) when compared to standard care (C) are effective, safe and cost-effective (O) as prophylaxis for COVID-19? |
| **Objectives** | **Primary objective**<br><br>To provide reliable estimates of the effect of trial treatments for each pairwise comparison with the standard care arm on SARS-CoV-2 infection, morbidity and mortality 60 days after randomisation (or appropriate point dependent on the intervention – where this differs to 60 days further detail will be provided in the specific investigational medicinal product (IMP) Appendix).<br><br>**Secondary objectives**<br>To assess the effects of trial treatments on mortality (all-cause and cause-specific), admission to hospital (all-cause and cause-specific), healthcare referrals for COVID-19, infection (asymptomatic, symptomatic), time to symptomatic infection and safety through serious adverse reactions.<br>To assess the effects of trial treatments on transmission of SARS-CoV-2 infection.<br><br>**Tertiary objectives**<br>To assess the cost-effectiveness of trial treatments and explain the contextual factors which influence trial processes including adherence to intervention and outcome measurement regimens, and which might impact on subsequent implementations of pre- or post-exposure prophylaxis for COVID-19 in care homes. |
| **Trial Configuration** | Overarching platform trial designed to provide reliable evidence on the efficacy of candidate therapies for preventing SARS-CoV-2 infection and transmission in care homes. |
| **Setting** | UK Care homes (residential, nursing, mixed). |
| **Sample size estimate per comparison of an active treatment with standard care** | In order to detect an odds ratio of 0.67, a sample size of 200 care homes per principal comparison (around 6,400 residents) is required assuming 1:1 randomisation, two-sided significance level 5%, power 90%, 4-level ordinal primary outcome (no infection proportion 60%, infection remain in CH 15%, admission to hospital all-causes 10%, all-cause mortality 15%), intra-cluster correlation 0.11, coefficient of variation for care home size 0.49, average |

| | |
|---|---|
| | number of residents per care home in trial 32, design effect/inflation factor 5.25. |
| **Number of participants** | Each comparison of an active treatment with standard care requires 200 care homes i.e. in the region of a total of 6,400 residents. |
| **Eligibility criteria** | **Care Home criteria**<br>***Inclusions***<br>• Location: UK care homes for older people, with and without nursing.<br>• Size: ≥20 beds in the care home in total.<br>***Exclusions***<br>• Care Quality Commission quality: Inadequate, or equivalent in devolved administrations.<br><br>**Care Home criteria at treatment phase**<br>***Exclusions:***<br>• Positive PCR or lateral flow test (or equivalent) for SARS-CoV-2 in any resident and/or staff within previous 4 weeks<br><br>**Resident criteria at trial entry**<br>***Inclusions***<br>• Resident in a Care Home.<br>• Age ≥65 years<br>• Able to give informed consent for participation or a personal legal representative has been identified who can give consent if resident lacks capacity.<br><br>***Exclusions***:<br>• Identified by care home staff to have entered end-stage palliative care.<br>• Resident in care home for short-term respite care.<br>• Resident's general practitioner is unable to support their involvement in the trial.<br><br>**Resident criteria at treatment phase**<br>***Exclusions***:<br>• Currently taking all of the trial interventions.<br>• Contraindication to all trial interventions - see IMP appendix.<br>• In treatment phase of another COVID-19 prevention or treatment trial |
| **Description of interventions** | **Intervention:** To be identified by NIHR Prophylaxis Oversight Group (POG) plus standard care. Each intervention will be described within a separate IMP Appendix, each of which will be updated upon confirmation of use within the trial.<br><br>**Comparator:** Standard care, i.e., no additional intervention. |

| | |
|---|---|
| **Randomisation and blinding** | The unit of randomisation is the care home, not the (individual) resident. Allocation concealment will be ensured by enrolling care homes and residents prior to revealing the allocation. There will be no blinding apart from the outcomes sourced from routinely collected health data. Where possible IMP interventions will be open-label.<br><br>Dynamic randomisation will be employed using a probabilistic minimisation algorithm to balance across important baseline characteristics including:<br>• Care home type (residential vs nursing vs nursing and residential dual registration)<br>• Prior COVID-19 in care home at any time (yes vs no)<br>• Size of Care Home - Total number of residents in care home (small (≤30 residents), medium (>30, ≤50 residents), large (>50 residents))<br>• Care home has capacity to give oxygen (yes vs no)<br><br>Trials of pre-exposure prophylaxis (PrEP, i.e. before the care home has a case) and/or post-exposure prophylaxis (PEP, once the care home has a new case to control an outbreak) will be conducted on the platform.<br><br>• For trials of PrEP, the care home will be randomised once an IMP has been identified by the NIHR POG and the necessary processes have been implemented. The care home must have had no evidence of SARS-CoV-2 infection for at least 4 weeks.<br>• For trials of PEP, care homes will only be randomised once they have an indication of a developing infection, e.g. recent positive PCR or lateral flow test (or equivalent) in a resident or member of staff (index case). The care home must have had no evidence of SARS-CoV-2 infection for at least 4 weeks prior to the index case. |
| **Outcome measures** | **Primary endpoint:**<br><br>Four-level ordered categorical scale. Participants will be classified according to the highest level, that is, the most serious event they experience during the 60-day period following randomisation:<br>1. No SARS-CoV-2 infection.<br>2. SARS-CoV-2 infection but resident remains in care home.<br>3. Admission to hospital, all-cause.<br>4. Death, all-cause.<br><br>SARS-CoV-2 status (positive or negative) will be diagnosed using PCR or lateral flow testing (or equivalent). A flow chart describing different scenarios of test results depending on which test(s) is (are) performed is presented in Figure A1 (Appendix).<br><br>**Secondary endpoints during the 60 days post-randomisation (unless otherwise stated):**<br><br>Clinical |

| | |
|---|---|
| | • Time to healthcare referral for COVID-19, e.g. discussion outside of care home with GP (excluding routine visit), 111, 999 paramedic or Emergency Department assessment, remote hospital consultation.<br>• Time to use of dexamethasone in the care home for COVID-19<br>• Time to use of oxygen in the care home for COVID-19<br>• Time to SARS-CoV-2 infection - positive PCR or lateral flow test (or equivalent) (i) with symptoms of COVID-19, (ii) without symptoms of COVID-19, (iii) total i.e. either with or without symptoms of COVID-19.<br>• Time to first admission to hospital<br>• Cause-specific hospital admission<br>• Time to death<br>• Days alive and not in hospital<br>• Cause-specific mortality, including COVID-19, stroke, pulmonary embolism, myocardial infarction<br>• Electronic frailty index at 60 days<br>• Ordinal outcome for the most serious event experienced during the 120 days post-randomisation with the following levels: 1. No SARS-CoV-2 infection, 2. SARS-CoV-2 infection but resident remains in care home, 3. Admission to hospital, all-cause, 4. Death, all-cause.<br><br>Safety<br>• Serious Adverse Reactions (SAR, excluding primary and secondary outcomes) and Suspected Unexpected SARs (SUSARs).<br>• Adverse events relevant to the intervention (see relevant IMP-specific Appendix)<br><br>Clinical – care home level<br>• Number of SARS-CoV-2 infections in residents in the care home (aggregate data including residents not participating in PROTECT-CH).<br><br>Economic evaluation<br>• EQ-5D-5L utilities and EQ-VAS at 60 days<br>• Quality Adjusted Life Years (QALY)<br>• Healthcare resource use and costs<br>• Incremental cost-per QALY and Net Monetary Benefit |
| **Statistical methods** | All comparative analyses will be based on contemporaneously enrolled care homes. The primary approach to between-group comparative analyses will be by intention-to-treat (i.e., according to randomised allocation regardless of adherence to trial allocation). Cluster-level and resident-level descriptive statistics will be used to illustrate balance between the groups at baseline. Primary comparative analyses will employ a multi-level ordinal logistic regression model with a random effect to account for clustering within care homes. The model will adjust for characteristics used in balancing random allocation between treatment arms and individual-level characteristics of age, sex, and vaccination status. |

#### 2.1. Sample size and justification

A total of 530 residents per group are required to detect an odds ratio of 0.67 for a 4-level ordinal primary outcome (no infection proportion 60%, infection remain in CH 15%, all-cause hospitalisation 10%, all-cause mortality 15%), assuming a two-sided significance level of 5% and 90% statistical power, with no adjustment for clustering [2].

Care homes of varying size will be included, and the number of residents recruited per care home will likely be in the range 20–60. Assuming an intra-cluster correlation of 0.11, a coefficient of variation for care home size of 0.49 (Lothian population analysis in 189 care homes[3]) and an average of 32 residents per care home in the study (assuming that the average number of beds per care home is 40[4] and that not all residents will take part in the study), this gives rise to a design effect or inflation factor of 5.25[5].

Therefore, to compare a single active treatment versus usual care, we will need in the region of 174 care homes, i.e., in excess of 5,500 residents. Allowing for the uncertainty surrounding the parameters listed above (e.g. levels of mortality and transmission rates are expected to be different in the second wave due to improved preparedness, better treatments and the potential impact of the expected vaccination programme, therefore it is possible that the observed proportions in the control group may differ), we propose a sample size of 200 care homes and a total number of residents in the region of 6,400 per comparison, with sample size re-estimation during the trial (once 60-day outcome data are available for at least 75% of residents randomised to standard care).

Therefore, comparing three active (unrelated) treatments concurrently versus standard care (in a 1:1:1:1 allocation ratio) would require 400 care homes in total, corresponding to around 13,000 residents.

#### 2.2. Blinding and breaking of blind

Care home residents and staff will not be blinded to treatment allocation. Dynamic randomisation of care homes using a minimisation algorithm and release of allocation only following enrolment, resident consent, and baseline data collection, will ensure allocation concealment. The trial statisticians, CI, and members of the TSC will remain blinded prior to treatment codes being revealed (for the final analysis of a comparison). Interim closed reports for the Data Monitoring Committee (DMC) will be prepared by a separate team of statisticians (unblinded to trial intervention) to those that will conduct the final analysis.

#### 2.3. Trial committees

A trial management group (TMG), platform steering committee (PSC) and data monitoring committee (DMC) will be assembled to oversee the trial. The general purpose, responsibilities and structure of the committees are described in the protocol. Further details of the roles and responsibilities of the PSC and DMC can be found in their respective charters agreed prior to the start of recruitment to the trial.

#### 2.4. Outcome measures

Primary and secondary outcomes will be assessed during the 60 days post-randomisation unless stated otherwise. The primary outcome is a four-level ordered categorical outcome. Participants will be classified according to the highest level, that is, the worst, event they experience during the 60-day period following randomisation:

1. No SARS-CoV-2 infection.
2. SARS-CoV-2 infection but resident remains in care home.
3. Admission to hospital, all-cause.
4. Death, all-cause.

The outcome measures and their sources are summarised in **Error! Reference source not found.**.

**Table 1:** Summary of the outcome measures

| Outcome measures | Description | Sources | | Type of data | Analysis method |
|---|---|---|---|---|---|
| | | **Care Home ascertainment through eCRF** | **Routine data sources** | | |
| **Primary outcome** | | | | | |
| A four-level ordinal outcome to capture the ability of the drug candidate(s) to prevent/reduce morbidity and mortality from COVID-19 in care home residents, and to reduce transmission in care home settings | Participants will be classified according to the highest level, that is, the most serious event they experience during the 60-day period (or relevant time point) following randomisation: | | | Ordered categorical | Mixed-effect ordinal logistic regression |
| | 1. No SARS-CoV-2 infection | ✓ | - | | |
| | 2. SARS-CoV-2 infection (infection diagnosed with PCR or lateral flow testing (or equivalent)) but resident remains in care home i.e., resident not admitted to hospital. The PCR test results will take primacy if both tests are performed as presented in the flowchart in Section 11.1 . | ✓ | ✓ | | |
| | 3. Admission to hospital, all-cause | ✓ | ✓ | | |
| | 4. Death, all-cause | ✓ | ✓ | | |
| **Secondary outcomes** | **(during the 60 days post-randomisation unless stated otherwise)** | | | | |
| **Clinical: individual resident-level** | | | | | |
| Healthcare referral for COVID-19 | Healthcare referral for COVID-19 e.g. discussion outside of care home with GP (excluding routine visit), 111 or, 999 paramedic or Emergency Department assessment , remote hospital consultation. | ✓ | ✓ | Time-to-event (first event) | Competing-risks survival regression with cluster–robust standard errors |

| Outcome measures | Description | Sources | | Type of data | Analysis method |
|---|---|---|---|---|---|
| Participant receives dexamethasone in the care home for COVID-19 | Use of dexamethasone in care home for COVID-19 | ✓ | - | Time-to-event | Competing-risks survival regression with cluster–robust standard errors |
| Participant receives oxygen in the care home for COVID-19 | Use of oxygen in care home for COVID-19 (for care homes with capacity to give oxygen) | ✓ | - | Time-to-event | Competing-risks survival regression with cluster–robust standard errors |
| Time to SARS-CoV-2 infection* - positive PCR or lateral flow test (or equivalent)<br>(i) with symptoms of COVID-19<br>(ii) without symptoms of COVID-19,<br>(iii) Total, i.e., either with or without symptoms of COVID-19. | Time from randomisation to SARS-CoV-2 infection - positive PCR or lateral flow test (or equivalent). The date of the LFT test will be used as the date of event if LFT is positive and confirmed by the PCR test.<br>(i) with symptoms of COVID-19<br>(ii) without symptoms of COVID-19<br>(iii) with/without symptoms of COVID-19 | ✓ (presence/ absence of symptoms) | ✓ (PCR test results) | Time-to-event | Competing-risks survival regression with cluster–robust standard errors |
| Time to first admission to hospital† | Time from randomisation to first hospitalisation for any reason | ✓ | ✓ | Time-to-event | Competing-risks survival regression with cluster–robust standard errors |
| Cause specific hospital admission | | ✓ | ✓ | | Descriptive |
| Days alive and not in hospital† | | - | ✓ | Continuous | Linear mixed-effect model |
| Time to death | Time from randomisation to death (all-cause) in days | ✓ | ✓ | Time-to-event | Shared frailty model |

| Outcome measures | Description | Sources | | Type of data | Analysis method |
|---|---|---|---|---|---|
| Cause-specific mortality, including COVID-19, stroke, pulmonary embolism, myocardial infarction | COVID-19 | ✓ | ✓ | Binary | Descriptive |
| | stroke | | | Binary | Descriptive |
| | pulmonary embolism | | | Binary | Descriptive |
| | myocardial infarction | | | Binary | Descriptive |
| Electronic frailty index at 60 days | Electronic frailty index (eFI)<br>The eFI is presented as a score (e.g. if 9 deficits are present out of a possible total of 36 the FI score = 0.25) - higher scores indicate increasing frailty. The eFI score is used to define frailty categories:<br>- Fit (0 - 0.12)<br>- Mild frailty (> 0.12 – 0.24)<br>- Moderate frailty (> 0.24 – 0.36)<br>- Severe frailty (> 0.36)<br>A composite endpoint strategy will be used to handle the eFI outcome truncated due to death by adding an extra category for death to ensure that death equates to the worst outcome. | - | ✓ | Ordered categorical | Mixed-effect ordinal logistic regression |
| Ordinal outcome for the most serious event experienced during the 120 days post-randomisation | Four-level ordinal outcome: No SARS-CoV-2 infection, SARS-CoV-2 infection but resident remains in care home, Admission to hospital, all-cause and Death, all-cause. | ✓ | ✓ | Ordered categorical | Mixed-effect ordinal logistic regression |
| **Clinical: care home level** | | | | | |

Written using WPD 16.5 version 3.0, 13-Sep-2018. Effective date: 13-Oct-2018. Template Author: Lucy Bradshaw

| Outcome measures | Description | Sources | | Type of data | Analysis method |
|---|---|---|---|---|---|
| Number of SARS-CoV-2 infections in residents in the care home | Aggregate infection rate in the care home. The numerator will be all confirmed SARS-CoV-2 infections including residents not participating in PROTECT-CH trial and the denominator will be the number of residents in the care home at randomisation. | ✓ | ✓ | Rate (Proportion) | Linear regression (rate difference) and Poisson regression (rate ratio) |
| **Additional Safety:** | | | | | |
| Serious Adverse Reactions (SAR, excluding primary and secondary outcomes) and Suspected Unexpected SAR (SUSAR) | | ✓ | | | |
| Adverse events relevant to the intervention as specified in relevant IMP specific appendix to protocol | | ✓ | | | |

*For PEP- only applicable for residents not positive at randomisation; †excludes residents with directive for no hospitalisation

### 3. INTERIM ANALYSIS

There are no formal interim analyses of clinical outcomes planned during the trial, hence no pre-planned design changes or provisions to avoid an inflation of the Type I error.

As part of continuous monitoring, the DMC will be provided with confidential report by trial arm, containing information on recruitment, protocol compliance, safety data, interim assessments of outcomes (between-group estimates of differences in efficacy and/or safety outcomes), and conditional power for futility assessment if necessary. The DMC will inform the TSC if, in its view, there is evidence or reason why the comparison within the platform should be modified or terminated prematurely, for example, if the conditional power at a given point is low. Under such circumstances where unblinded information on efficacy is necessary and randomisation of care homes is still ongoing, the impact on the Type I error will be properly taken into consideration. If the TSC and sponsor decide that the recommended changes be implemented, then this will be done via study protocol amendments.

### 4. GENERAL ANALYSIS CONSIDERATIONS

#### 4.1. Framework

The overall objective of the platform trial is to test the superiority of each intervention to standard care.

#### 4.2. Analysis sets

| Outcome | Analysis set |
|---|---|
| *Primary, secondary and safety outcomes – individual-level* | Contemporaneously randomised care homes and their eligible residents analysed according to the group their care home was randomly allocated to regardless of adherence to trial allocation (Intention-to-treat [ITT] analysis population).<br><br>Since not all residents will be eligible for all trial interventions, residents in homes allocated to standard care will only be included in a given comparison if they were eligible to be given the intervention of interest had their home been randomised to it (i.e. not taking the intervention of interest, or another drug in the same class, and did not have any contraindications to it).<br><br>In the case where an outbreak based on a positive result from a lateral flow test is not confirmed with a PCR test, then the site remains in the trial in the ITT analysis population.<br><br>Primary analysis will be for participants with complete outcome data collected (i.e., without imputation for missing data). |
| Care home level infection rate | All residents in contemporaneously randomised care homes including residents not consenting to PROTECT-CH included in the comparison. |

| Outcome | Analysis set |
|---|---|
| Supplementary analyses (for primary outcome and the secondary outcome related to transmission (time to SARS-CoV2 infection)) | All residents with consent regardless of whether eligible/took part in treatment phase of trial, analysed according to the group their care home was randomly allocated. |

#### 4.3. Timing of primary analysis

The primary analysis for each intervention will be performed once all participants in the active and usual care groups have reached 60 days of follow-up (or appropriate time point as detailed in the IMP appendix) and have outcome data available from both the care home (via the trial database) and the central routine data repository.

The secondary analysis of the ordinal outcome at 4 months will be performed once all participants in the active and usual care groups have reached this time point after randomisation and have outcome data available (as described above).

#### 4.4. Statistical software

Analyses will be performed using Stata version 16 or above or R version 3.6.2 or above as appropriate.

#### 4.5. Procedures for missing data

**Missing baseline data**

Missing data for the baseline covariates in the primary analysis (cluster-level minimisation factors and selected individual-level characteristics) is expected to be very rare. However, any missing baseline data will be imputed using the mean score at each care home in order to be able to include these participants in the analysis. These simple imputation methods are superior to more complicated imputation methods when baseline variables are included in an adjusted analysis to improve the precision of the treatment effect [6].

**Missing outcome data**

Missing values for the components of the composite primary outcome is expected to be very minimal as data collection for primary outcome will be combined with linkage to routine clinical data. In addition, ascertainment of the primary outcome measure will be performed centrally using UK death, hospitalisation and COVID-19 registers. As a result, primary analysis will be based on complete cases with the assumption that missingness is independent of the outcome, given the covariates. No imputation is planned.

### 5. DESCRIPTION OF PARTICIPANT CHARACTERISTICS

#### 5.1. Participant flow

A CONSORT diagram will be used to summarise the flow of care homes into the trial and the flow of residents within care homes over the course of the trial. The flow diagram will include the number of care homes assessed for eligibility and those randomised. For each treatment group, the numbers of care homes that were randomly assigned, as well as the number of care homes and residents within care homes that received allocated treatment and were analysed for primary outcome will be presented. Post-randomisation discontinuations will be presented by treatment group and overall, plus reasons.

#### 5.2. Baseline characteristics

Baseline characteristics for cluster and individual participant levels will be summarised both overall and by treatment groups using appropriate descriptive statistics for continuous (mean, standard deviation, median, lower & upper quartiles, minimum, maximum and number of observations) and categorical (frequency counts and percentages) characteristics to illustrate balance between the groups at baseline. Baseline summaries will be based on the principal comparisons (between each treatment group and the concurrent standard care group respectively).

Cluster-level characteristics will include:

- Minimisation variables
    - care home type (residential vs nursing vs nursing and residential dual registration)
    - prior COVID-19 in care home at any time (yes vs no)
    - size of care home - Total number of residents in care home (small ($\leq$30 residents), medium (>30, $\leq$50 residents), large (>50 residents))
    - care home has capacity to give oxygen (yes vs no)
- Any residents in care home received vaccination against COVID-19
- Any staff in care home received vaccination against COVID-19
- Country
- Care home ownership
- Registered for clients with learning disabilities
- CQC rating (outstanding, good, requires improvement) or equivalent in devolved administrations.
- Time between positive test for COVID-19 in resident/member of staff and care home randomisation (For PEP only)

Individual resident level baseline demographic and clinical characteristics will include:

- Age
- Sex
- Ethnicity
- BMI (from height and weight)
- Advance directive for no hospitalisation
- Do Not Attempt Resuscitation order
- Smoking status
- Co-morbidities (diabetes, heart disease, chronic lung disease, severe liver disease, severe kidney impairment, dementia, stroke)
- Previous positive test for COVID-19
- Tested positive for COVID-19 at randomisation (For PEP only)
- Received vaccination for COVID-19
- Time in care home
- Electronic frailty index
- Type of consent

### 6. ASSESSMENT OF STUDY QUALITY

#### 6.1. Randomisation

The unit of randomisation is the care home, and thus eligible consenting individual residents will follow the treatment pathway to which their care home is randomised. Dynamic randomisation will be employed using a minimisation algorithm to balance across important baseline characteristics including:

- Care home type (residential vs mixed nursing/residential vs. nursing)
- Prior COVID-19 in care home at any time (yes vs no)
- Size of Care Home - Total number of residents in care home (small (<30 residents), medium (>30, <50 residents), large (>50 residents))
- Care home has capacity to give Oxygen (yes vs no)

A team of central PIs will use the remote, internet-based randomisation system to obtain the treatment allocation for each care home after confirmation of care home participation and eligibility checks for the residents.

For trials of PrEP the care home will be randomised once an IMP has been identified by the NIHR POG and the necessary processes have been implemented.

For trials of PEP the care home will be randomised once an IMP has been identified by the NIHR POG and the necessary processes have been implemented and a resident or staff member in the care home has tested positive for SARS-CoV-2.

The number of care homes randomised to each treatment group will be tabulated. The minimisation variables will be tabulated (for each separate pairwise comparison of active treatment with the standard care treatment arm), as part of the baseline characteristics. The unlikely event of a care home withdrawing from the study will be reported with the timing and reasons for withdrawal, if available.

#### 6.2. Adherence

The number and proportion of residents who did not receive the treatment they were allocated to as intended will be reported. Details on the number of days (or doses) of treatment received will be reported for all trial treatments received where available. Cluster-level non-adherence, if any, will also be reported alongside the participant non-adherence.

Information will also be reported on receipt of relevant concomitant medications received as part of the resident's usual care during the treatment period (e.g. steroid inhalers and oral steroids).

#### 6.3. Follow-up and discontinuations

All reasonable efforts will be taken to minimise loss to follow-up, which is expected to be minimal as data collection for primary and secondary outcomes using trial-specific eCRFs is combined with linkage to routine clinical data on study outcomes. In addition, ascertainment of the primary outcome measure will also be performed centrally using UK death, hospitalisation and COVID-19 registers.

The number and percentage of homes and participants with follow-up information at day 60 days after randomisation will be reported by treatment groups.

#### 6.4. Protocol deviations

Non-compliance with allocated treatment will be reported as described in Section 6.2 (Adherence).

Protocol violations are defined in the protocol. Non-compliance with the protocol will be reported on a deviation form and assessed by the NCTU to determine if constitutes a violation. The number of participants with protocol violations will be summarised by treatment group along with the type of deviation. Protocol violations will also be listed.

### 7. ANALYSIS OF EFFECTIVENESS

The analysis and reporting of the trial will be in accordance with CONSORT guidelines for cluster[7] and adaptive[8] designs. Analyses will be based on pairwise comparisons between each treatment arm and the concurrent standard care arm (reference group). Primary comparative analyses will be based on intention-to-treat principle, with all care homes and residents (eligible for the intervention prior to randomisation) within these care homes analysed according to treatment to which the care home was randomised. All tests will be two-tailed with point estimates and 95% confidence intervals for the treatment effect presented. No formal adjustment for multiple significance testing will be applied [9]. Secondary outcomes will be considered supportive to the primary outcome.

#### 7.1. Primary analysis

Primary between-group comparison will be based on individual-level data rather than cluster-level summaries. Primary comparative analyses will employ a multi-level mixed-effects ordinal logistic regression model to reflect the ordinal nature of the primary outcome and allow for the between-cluster variation (clustering within care homes). A random intercept model will be fitted. The model will adjust for all the cluster-level minimisation factors and individual-level factors including age, sex, and vaccination status (ordinal: no vaccination, partial dose, full dose) as fixed effects. These individual-level factors have been shown to contribute to disparities in fatality rates in COVID-19 patients. The treatment comparison will be presented as an adjusted common proportional odds ratio together with 95% confidence interval, and p-value.

The ordinal logistic regression model is based on the proportionality of odds (parallel lines) assumption, that is, the treatment effect is consistent across the whole composite ordinal primary outcome spectrum. We will test for non-proportionality using a Likelihood Ratio test comparing the model where the coefficients are constrained to be equal across the three log-odds contrasts and the model where the proportional odds assumption is relaxed. If the data sets show significant departures from the proportionality assumption, then an extension of the multilevel ordinal logistic regression model to allow for non-proportional odds will be employed.

#### 7.2. Sensitivity analysis of primary outcome

The following sensitivity analyses will be performed on the primary outcome:

- Inclusion of symptomatic residents without a positive test in the "SARS-CoV-2 infection but resident remains in care home" category of ordinal outcome.
- Further adjustment for any other covariate not pre-specified in the primary analysis that is predictive of the primary outcome and with marked imbalance between treatment arms at baseline.

#### 7.3. Subgroup analysis of primary outcome

Subgroup analyses will be conducted for the ordinal primary outcome and for all-cause mortality for the following factors for each separate principal comparison:

- Care home type (residential vs mixed nursing/residential vs. nursing)
- Prior COVID-19 in care home at any time (yes vs no)
- Size of Care Home - Total number of residents in care home ((small (<30 residents) vs medium (≥30 to <50 residents) vs large (≥50 residents))
- Care home has capacity to give Oxygen (yes vs no)
- age group (<80 vs 80-89 vs ≥ 90 years old)
- sex (female vs male)
- vaccination status (none, partial, full)

The analysis will be conducted by including an interaction term with the allocated treatment in the analysis model (mixed-effects ordinal logistic regression for primary outcome and shared frailty model for all-cause mortality) to assess whether there is evidence that the effects of subgroups differ substantially from the overall effect seen in all patients combined. The models will adjust for the same variables as the primary analysis. Between-group treatment effects will be provided for each subgroup, but interpretation of any subgroup effects will be based on the treatment-subgroup interaction and 95% confidence interval. Results will be presented on forest plots containing the estimated effects with confidence intervals. The descriptive statistics and interaction coefficients and CIs will be presented regardless of the interaction p-values to provide information for any future more targeted trials. The subgroup analyses will be regarded as exploratory as the trial is not powered to detect interactions.

#### 7.4. Secondary outcomes

Table 1 lists all the secondary outcomes and the type of variables.

Individual-level secondary outcomes will be analysed using mixed-effects regression models appropriate for the type of outcome variable, adjusting for minimisation variables and individual-level factors including age, sex, and vaccination status as fixed effects with a random effect for care home.

Ordered categorical outcomes will be analysed similarly as the primary outcome.

Continuous outcomes will be analysed using linear mixed-effect model, with further adjustment for baseline outcome measure if available. The estimated between group effect will be presented using the adjusted difference between means, along with a 95% confidence interval.

Binary variables will be analysed using mixed effects logistic regression model. The between group effect will be reported using an adjusted risk difference and adjusted risk ratio along with corresponding 95% confidence intervals, obtained using Stata's Margins command with standard errors computed using the delta method .

All-cause mortality (time to death) will be analysed using mixed-effects Cox regression model (shared frailty model). A Kaplan-Meier curve will be presented by trial arm. Other time-to-event outcomes will be analysed using competing-risks survival regression with cluster–robust standard errors. The between group effect will be reported using an adjusted hazard ratio along with a corresponding 95% confidence interval. Estimated cumulative incidence function graphs will be presented for each of the outcomes by trial arm.

The care home level infection rate outcome will be analysed using multiple linear regression of the cluster proportions, adjusted for the cluster-level minimisation variables, to obtain the adjusted difference in mean rates between the intervention and control arms along with the corresponding 95% CI. Poisson regression on cluster-level rates, adjusted for the cluster-level minimisation variables, will be used to obtain estimated rate ratio with corresponding 95% CI. The regression models will be weighted by the inverse of the variance of each cluster summary to take account of variability in cluster size. For this outcome, there will be no adjustment for individual-level covariates since not all residents will have individual level infection data to allow for individual-level analysis.

##### 7.5. Supplementary analysis of primary outcome and key secondary outcomes

For the primary outcome and the secondary outcome related to transmission (time to SARS-CoV2 infection) additional analysis will also include all residents with consent regardless of whether eligible/took part in treatment phase of trial.

#### 8. ANALYSIS OF SAFETY

##### 8.1. Serious adverse events (SAEs)

Safety reporting in the trial will focus only on those events that could be related to the trial medication, are serious in nature (according to the SAE definition in the protocol) and are unexpected (either the event itself or in severity) according to the reference safety information (RSI) for the IMP. Further information regarding the RSI can be found in the appendix in the protocol for the relevant IMP.

Descriptive summaries for each treatment group will be presented to show:

- The number and percentage of residents with at least one SAE and the total number of SAEs.
- The number and percentage of residents with at least one SAR (serious adverse reaction, related to IMP not unexpected), the total number of SARs and number with each event (MedDRA preferred term)
- The number and percentage of residents with at least one SUSAR (Suspected Unexpected Serious Adverse Reaction), the total number of SUSARs and the number with each event

Serious adverse reactions (SARs) and suspected unexpected serious adverse reactions (SUSARs) will be listed by treatment group.

##### 8.2. Adverse events relevant to the intervention

Adverse events relevant to the intervention as specified in the relevant IMP-specific protocol appendix will be reported on the eCRF by asking care home staff if the resident has experienced the event. These adverse events will be presented descriptively by treatment group for each pairwise comparison.

#### 9. FINAL REPORT TABLES AND FIGURES

The trial will be reported according to the principles of the CONSORT statements[7, 8, 10, 11]. The exact composition of the trial publication(s) depends on the availability of drugs, and the findings from the various pairwise comparative analyses in the main trial.

Dummy tables version 1.0  is appended to this SAP.

#### 11. Appendix

##### 11.1. SARS-CoV-2 infection scenarios after a test in PROTECT-CH

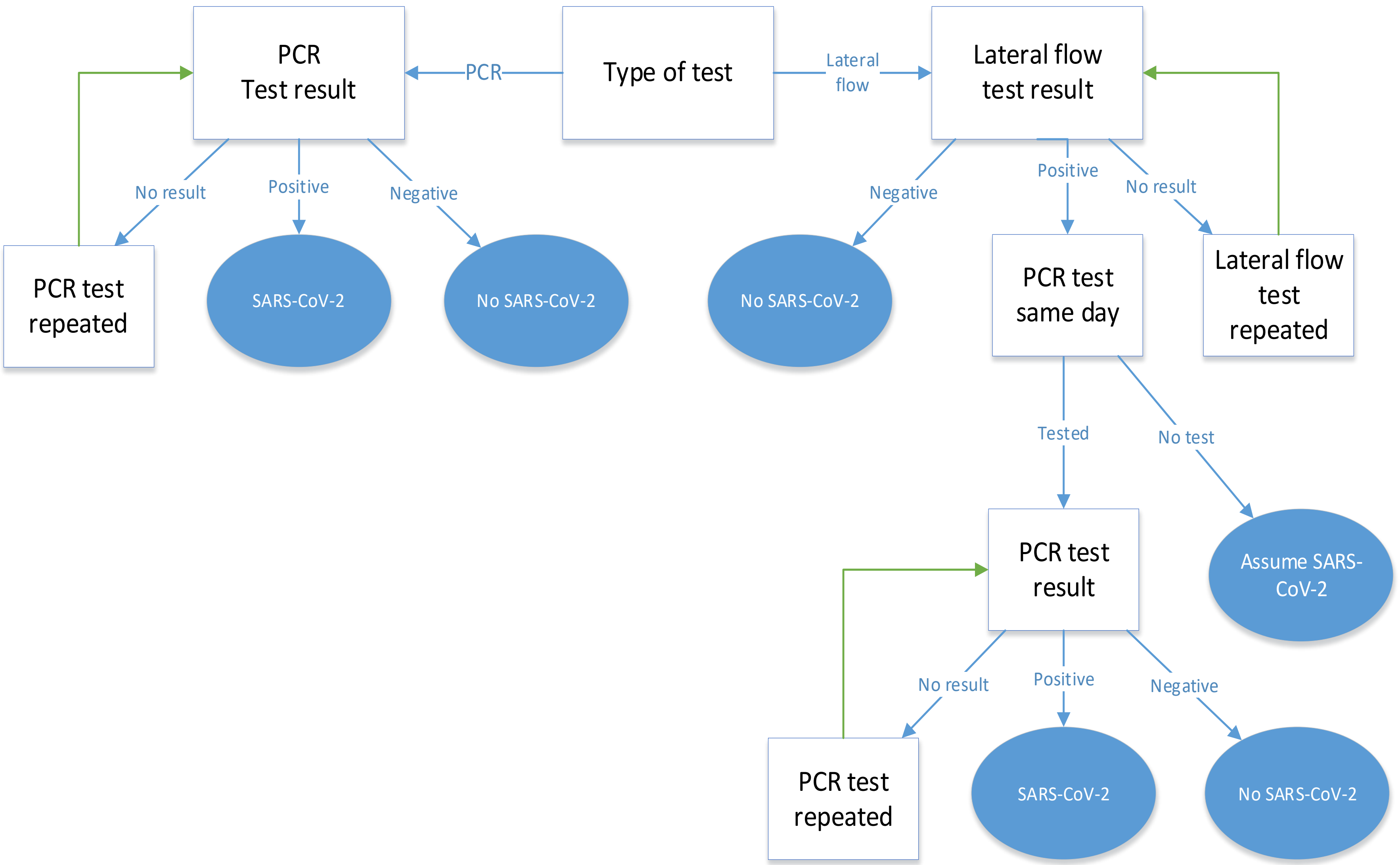
