## Appendix 3. Health Economic Analysis Plan for "Prophylactic Treatment of COVID-19 in Care Homes Trial (PROTECT-CH)"

UNIVERSITY OF SURREY

### Prophylactic Therapy in Care Homes Trial-CH (PROTECT-CH)
### Health Economic Analysis Plan (HEAP)

**Authors:** Matt Glover (University of Surrey), Heather Gage (University of Surrey)

**Approval**

| **Name** | **Job title** | **Trial Role** | Signature | Date |
|---|---|---|---|---|
| Philip Bath | Professor of Stroke Medicine | Chief Investigator | [signature] | 20/10/2021 |
| Alistair Burns | Professor of Old Age Psychiatry, | Chair of the Platform Steering Committee | [signature] | 21.10.2021 |

#### Version 1.0

#### 15th October 2021

| HEAP Version number | Detail of revisions with reason | Initials |
|---|---|---|

#### Associated Documents

| Document | Version | Date |
|---|---|---|
| PROTECT-CH Economic Evaluation Dummy Tables | V1.0 | 15th October 2021 |
| PROTECT-CH Protocol | V1.0 | 27th April 2021 |
| PROTECT-CH Statistical Analysis Plan | V1.0 | 8th October 2021 |
| PROTECT-CH Dummy Tables | V1.0 | 8th October 2021 |

### 2. List of tables and figures

Table 1: Healthcare resource use data sources ........ 6

#### 3. Abbreviations

| Abbreviation | Description |
|---|---|
| **A&E** | Accident and emergency |
| **COVID-19** | Coronavirus-induced disease-19 |
| **eCRF** | e-case report form |
| **EQ-5D-5L** | EQ-5D-5L health related quality of life instrument |
| **EQ-VAS** | Health related quality of life visual analogue scale |
| **HCRU** | Healthcare resource use |
| **HES** | Hospital Episodes Statistics |
| **HRQoL** | Health related quality of life |
| **ICER** | Incremental cost-effectiveness ratio |
| **INMB** | Incremental net monetary benefit |
| **IQR** | Interquartile range |
| **MAR** | Missing at random |
| **MCAR** | Missing completely at random |
| **NMB** | Net Monetary Benefit |
| **NHS** | National Health Service |
| **NIHR** | National Institute for Health Research |
| **PEP** | Post-exposure prophylaxis |
| **POG** | Prophylaxis Oversight Group |
| **PrEP** | Pre-exposure prophylaxis |
| **PROTECT-CH** | Prophylactic Therapy in Care Homes Trial-CH |
| **PSSRU** | Personal Social Services Research Unit |
| **SAP** | Statistical Analysis Plan |
| **SARS-CoV-2** | Severe acute respiratory syndrome coronavirus |
| **QALYs** | Quality Adjusted Life Years |

#### 4. Introduction

This document details the proposed handling of data, analysis and presentation of results for economic outcomes of treatments compared in the PROTECT-CH randomised clinical trial. Associated documents, including the study protocol, are listed on the title page and this analysis plan should be interpreted alongside these.

Where possible, analyses and data handling are consistent with the approach to other clinical outcomes (as described in the SAP), however, any deviations are detailed herein.

In brief, PROTECT-CH is a clinical trial with a pragmatic adaptive cluster-randomised parallel group platform design, to compare a suite of interventions to prevent COVID-19 infection and reduce severity/transmission and death in care homes. Interventions may either be administered as pre-exposure prophylaxis (PrEP), where care homes will be randomised once residents have consented to the trial or as post-exposure prophylaxis (PEP), where care homes will only be randomised once they have an indication of a developing infection.

The trial will recruit residents in UK care homes (England, Wales, Scotland, Northern Ireland), with and without on-site nursing staff, aged 65 years and over.

The primary endpoint is a four-level ordered categorical scale at 60 days post randomisation (1. No SARS-CoV-2 infection. 2. SARS-CoV-2 infection but resident remains in care home. 3. Admission to hospital, all-cause. 4. Death, all-cause). Secondary endpoints include healthcare referrals, use of COVID-19 specific treatments in care homes and time to infection, hospitalisation and death as well as the ordinal outcome used for primary endpoint, including components, measured at 120 days. Additional safety outcomes will also be assessed.

Interventions included in the trial will be those specifically identified by NIHR Prophylaxis Oversight Group (POG) plus usual care and the comparator will be usual care.

#### 5. Aim

The aim of the economic evaluation is to determine the cost-effectiveness of prophylactic interventions to prevent COVID-19 infection and reduce severity/transmission and deaths in care home residents (UK, ≥65 Years old) compared to usual care, from an NHS perspective.

#### 6. Design

The primary economic evaluation will be a within trial cost-utility analysis, based on 60-day follow-up. Incremental costs (including any potential savings) associated with prophylaxis for care-home residents will be estimated. Health related quality of life measured using the EQ-5D-5L and EQ-VAS (proxy report) at 60 days will be used to compute Quality Adjusted Life Years (QALYs) and estimate incremental QALYs. Costs and QALYs will be combined to estimate the incremental cost-effectiveness ratio (ICER) or cost per QALY of interventions compared to usual care, and present incremental net monetary benefit (INMB) at various willingness to pay thresholds.

#### 7. Data collection and preparation

PROTECT-CH will use routine healthcare data sources to inform data collection, alongside an eCRF to collect data directly in care homes. Where possible, patient level data will be collected from routine sources to inform economic outcomes. Once data are available for analysis, all cleaning and analysis will be conducted using Stata V15] (StataCorp LP).

#### 1.1. Healthcare resource use

Healthcare resource use (HCRU) data collection was designed to be parsimonious and feasible, given the scope and scale of the platform study. Items to be measured reflect attempts to capture the predominant primary and secondary care contacts and admissions experienced in the care home population (and associated with frailer populations), as well as encompassing possible HCRU related to COVID-19 symptoms and complications that may be impacted by prophylaxis[1–4].

Healthcare resource use data will be collected by electronic case report form (eCRF) and from external routine sources, including Hospital Episode Statistics (England) and equivalent databases for other UK nations (Table 1). Data collection in the eCRF will be completed at 60 days post care home randomisation, based on care records and staff completion and attendances and admissions for the 60-day period extracted from external routine sources. For healthcare contacts that may occur face-to-face or remotely, this distinction will be made in eCRF data collection and unit costs appropriately differentiated.

***Table 1: Healthcare resource use data sources***

| **Resource use item** | **Routine data sources** | **eCRF** |
|---|---|---|
| GP visit | | X |
| Nurse visit (Practice Nurse, Nurse Specialist) | | X |
| Allied Health Professional visit<br>(Physiotherapist, Occupational Therapist, Respiratory Therapist, Speech and Language Therapist) | | X |
| Outpatient visit | | X |
| 999 call | | X |
| 111 call | | X |
| Ambulance attendance | X | |
| A&E attendance | X | |
| Inpatient admission | X | |
| Critical care stay | X | |

#### 7.1. Unit costs

Each resource use item will be assigned a unit cost using nationally validated tariffs such as PSSRU [5], NHS reference costs [6], or wider literature where necessary. Unit costs will be detailed as shown in Dummy Table 5[7]. Unit costs will be collected for an NHS perspective and presented in 2019/20 prices, adjusting for inflation using published indices where necessary[5].

#### 7.2. Intervention costs

Each intervention unit cost will be determined according to individual treatment protocol, accounting for administration and acquisition costs taken from national sources (e.g. BNF [8]) [Dummy Table 7][7].

##### 1.2. Health-related quality of Life (HRQoL)

The outcome of interest for health-related quality of life is the EQ-5D-5L, a generic preference-based instrument. These data will be collected through eCRF at screening after consent and 60-day follow-up, both by self-report, where a resident has capacity to complete the questionnaire and proxy report (completed by carer). For PEP, where care homes are randomised once an outbreak occurs, there may be a delay between collection of EQ-5D-5L at baseline and randomisation.

The UK crosswalk tariff [9] will be used to derive utility scores from responses and combined with survival data. Utility values will be integrated over time to provide accrued QALYs for each participant during the 60-days, using the area under the curve method. Where a care home resident dies during follow-up, a utility score of 0 will be assigned on date of death for purpose of QALY computation.

#### 8. Handling missing cost and HRQoL data

The study has been designed to minimise missing data, but the possibility of missing outcomes data required to perform the economic evaluation cannot be ruled out. For instance, some external routine data may not be available for all four nations of the UK. Reporting missing data in cost-effectiveness analysis, including EQ-5D responses, is common [10–12].

If data is ‘Missing Completely At Random’ (MCAR), that is that the likelihood of missing is not related to the unobserved value of the variable, nor is it correlated with any other observed variable, then it can be ignored since this will not introduce bias. However, if the proportion of cases with incomplete data is large then it may produce results that are imprecise. Most methods for dealing with missing data require data to be at least ‘Missing at Random’ (MAR) - that the likelihood of a data point being missing for variable ‘x’ is not correlated with the unobserved value of x but may be corelated with other observed variables. To test the MAR and MCAR assumptions the pattern of missing data will be examined. The association between missingness and baseline values will be explored through logistic regression.

If data are assumed MAR[1], multiple imputation will be used for the primary economic analysis. A linear regression model of covariates with complete data will be specified, to describe those variables with missing data. The specification of this model will take account of the hierarchical nature of the dataset[13]. Multiple imputation by chained equations will be performed, which adopts an iterative procedure using Markov Chain Monte Carlo (MCMC) equations until coefficient estimates for the missing variables become stable. Once stable, a full imputed data set is created. This process will be set for 5-10 Imputed sets, depending on the percentage of complete cases. If the proportion of missing cases is higher, the number of imputed data sets will be increased.

Depending on the nature of missing HRQoL data, values will be imputed at utility index level[14] and QALYs appropriately constructed. Where missing resource items exist, imputation will take place at cost level, with sensitivity analysis performed for imputation of the sum of costs for each collection source. Analysis will use Stata mi estimate commands to adjust standard errors using Rubin’s combination rules.

[1] If data is believed to be NMAR, further steps will be taken to specify an appropriate model, to minimise the risk that any imputation leads to bias estimates and explore impact of different imputation specification in sensitivity analysis. The exact nature of this model will depend on the missing data and will determined once analysis commences.

### 9. Economic Evaluation

Costs, HRQoL and QALYs at 60-days will be analysed as separate outcomes, before cost-utility analysis using cost and QALYs is conducted. Costs, EQ-5D-5L (proxy and self-report) and QALYs (based on proxy report EQ-5D-5L) will be presented as means at baseline and follow-up, with associated standard deviation, median (and IQR), maximum and minimum values.

#### 1.3. Reporting and analysis of healthcare resource use costs

Descriptive data (mean, standard deviation) on healthcare resource use will be presented at the item level along with associated costs [Dummy Table 6 and 8][7]. Patient level healthcare resource use costs for each item will be summed to compute total costs per patient and mean costs per group.

Total costs accrued over the 60-day follow-up will be analysed in line with other outcomes, using linear mixed effects models to take account of clustering and adjusting for minimisation variables and individual-level factors including age, sex, and vaccination status as fixed effects.

The estimated between group effect will be presented using the adjusted difference between means, along with a 95% confidence interval. A histogram of total costs at 60-days will also be presented [Dummy Figure 3][7].

#### 1.4. Reporting and analysis of HRQoL and QALYs

EQ-5D-5L descriptive system responses will be tabulated at baseline and 60-day follow-up [Dummy Table 1 to 4][7]. Changes from baseline in intervention and comparator groups for each of the levels within dimensions (%) will be presented alongside bar charts [Dummy Figure 1 and 2][7]. No formal analyses will be conducted on descriptive system responses.

The primary analysis of HRQoL will be based on EQ-5D-5L proxy report. It is expected that up to 76% of residents will lack capacity to consent to the trial[15], and will be consented using a legal representative. Capacity to consent will be used as an indicator of whether to approach the resident to complete an EQ-5D-5L self-report. Some residents with capacity may still feel unable to complete by self-report even after guidance. Therefore, the expected proportion of residents for which self-reported EQ-5D-5L could be available is expected to be less than 20% and capacity could change between baseline collection and follow-up.

However, studies have shown there may be systematic differences between proxy and self-report responses, which could affect both absolute estimates of HRQoL and between group estimates of effect[16–19]. EQ-5D-5L self-report and proxy report response patterns will be explored and reporting subgroups (self-report vs proxy) examined in sensitivity analysis.

EQ-5D-5L utility values, EQ-VAS and QALYs at 60-days will be analysed in line with other outcomes, using linear mixed effects models to take account of clustering and adjusting for minimisation variables and individual-level factors including age, sex, and vaccination status as fixed effects, including baseline EQ-5D-5L.

The estimated between group effect will be presented using the adjusted difference between means, along with a 95% confidence interval.

#### 1.5. Cost-utility analysis

Cost-effectiveness will be assessed using the ICER (cost per QALY) and INMB at various willingness to pay thresholds (£15,000, £20,000, £30,000 per QALY).

NMB will be analysed in line with other outcomes, using linear mixed effects models to take account of clustering and adjusting for minimisation variables and individual-level factors including age, sex, and vaccination status as fixed effects, including baseline EQ-5D-5L utility. The estimated between group effect will be presented using the adjusted difference between means, along with a 95% confidence interval at each willingness to pay threshold.

In addition to analysis of NMB in line with other outcomes, to characterise uncertainty in ICERs and facilitate presentation of Cost-Effectiveness-Acceptability-Curves (CEAC) to aid resource allocation decisions, non-parametric bootstrap with replacement will be used to estimate a distribution of incremental costs and effects (or ICERs)[20]. Bootstrap samples will be pulled (after multiple imputation of missing data if necessary) accounting for the clustered nature of data, and a bivariate linear mixed-effect model run within each bootstrap. The number of iterations will be chosen to ensure convergence in estimates of mean ICER (or INMB). A 95% confidence interval can be estimated from these data, but the likelihood of cost-effectiveness will be presented probabilistically. These will be presented on a Cost-Effectiveness-Plane and using the estimated Net-Benefit of treatment to populate the CEAC. The CEAC describes the probability the intervention will be cost-effective compared to usual care at varying willingness to pay thresholds per QALY. For example, NICE indicate a threshold of £20,000–30,000 per QALY be achieved for an intervention to be deemed cost-effective, whilst other empirical estimates of opportunity suggest the threshold may be lower[21,22].

#### 1.6. Sensitivity and subgroup analysis

**Self-report EQ-5D-5L**

Given the issues of capacity highlighted previously, proxy EQ5D responses will used in the primary health economic analyses. However, if investigation of response patterns indicates systematic differences, and sufficient data is collected (% tbc), sensitivity analysis using self-report will be conducted.

**Model specification**

Alternative model specifications will be explored for cost and effects if appropriate and presented alongside primary analysis. For instance, the use of Generalised Linear Models for potentially skewed outcomes will be explored and alternative model specifications included as sensitivity analysis.

**Subgroup analyses**

Exploratory subgroup analyses will be performed in line with subgroup analyses conducted for the ordinal primary outcome as outlined in the SAP, using INMB at a willingness to pay threshold of £20,000 as the cost-effectiveness outcome.

#### 10. Longer-term cost-effectiveness analysis

The primary analysis will be based on the 60-day follow-up, but some data will be available up to 120 days post randomisation which can be utilised in a longer-term cost-effectiveness analysis.

EQ-5D-5L data is not being collected beyond 60 days. QALYs will be constructed using the area under the curve approach using last utility value carried forward, or imputing 0 on the date death has occurred during follow-up and linear interpolation.

Healthcare resource use from routine data sources will be collected up to 120 days post randomisation. Two methods of analysis will be conducted. One using observed data, which will exclude some primary care healthcare resource use not available from routine sources and one with multiple imputation methods and describe in Section 8.

Cost-effectiveness will be assessed using INMB at various willingness to pay thresholds (£15,000, £20,000, £30,000 per QALY). NMB will be analysed in line with other outcomes, using linear mixed effects models to take account of clustering and adjusting for minimisation variables and individual-level factors including age, sex, and vaccination status as fixed effects, including baseline EQ-5D-5L utility. The estimated between group effect will be presented using the adjusted difference between means, along with a 95% confidence interval at each willingness to pay threshold [Dummy Table 10][7].

7. Glover M, Gage H, PROTECT-CH Trial Team. *Prophylactic Therapy in Care Homes Trial-CH (PROTECT-CH): Health Economic Analysis Dummy Tables and Figures V1.0*.

8. BNF British National Formulary - NICE. Accessed March 25, 2021. https://bnf.nice.org.uk/

9. Van Hout B, Janssen MF, Feng YS, et al. Interim scoring for the EQ-5D-5L: Mapping the EQ-5D-5L to EQ-5D-3L value sets. *Value Heal*. 2012;15(5):708-715. doi:10.1016/j.jval.2012.02.008

10. Manca A, Palmer S. Handling missing data in patient-level cost-effectiveness analysis alongside randomised clinical trials. *Appl Health Econ Health Policy*. 2005;4(2):65-75.

doi:10.2165/00148365-200504020-00001

11. Faria R, Gomes M, Epstein D, White IR. A Guide to Handling Missing Data in Cost-Effectiveness Analysis Conducted Within Randomised Controlled Trials. *Pharmacoeconomics*. 2014;32(12):1157-1170. doi:10.1007/s40273-014-0193-3

12. Gabrio A, Mason AJ, Baio G. Handling Missing Data in Within-Trial Cost-Effectiveness Analysis: A Review with Future Recommendations. *PharmacoEconomics - Open*. 2017;1(2):79-97. doi:10.1007/s41669-017-0015-6

13. Gomes M, Díaz-Ordaz K, Grieve R, Kenward MG. Multiple imputation methods for handling missing data in cost-effectiveness analyses that use data from hierarchical studies: An application to cluster randomized trials. *Med Decis Mak*. 2013;33(8):1051-1063. doi:10.1177/0272989X13492203

14. Simons CL, Rivero-Arias O, Yu LM, Simon J. Multiple imputation to deal with missing EQ-5D-3L data: Should we impute individual domains or the actual index? *Qual Life Res*. 2015;24(4):805-815. doi:10.1007/s11136-014-0837-y

15. Logan PA. Evaluation of the Guide to Action care Home Fall Prevention Programme in Care Homes for Older People: A Multi-Centre, Single Blinded, Cluster Randomized Controlled Trial (FiNCH). *NIHR J Ser*. 2020;In Press.
